# ICONIC: An R Package for Integrating Instrumental Variable- and Negative-Control-Informed Causal Discovery and Diagnostics in Multiomic Studies

**DOI:** 10.64898/2026.08.26.26361466

**Authors:** Sean T. Bresnahan, Charis Xiong, Taylor Head, Yung-Han Chang, Arjun Bhattacharya, Jonathan Y. Huang

## Abstract

Unmeasured confounding threatens causal inference and replicability in observational multi-omic studies across variable environments. Genetic instrumental variables (Mendelian randomization) and negative-control calibration each address complementary sources of unmeasured confounding, yet no existing framework unifies them for omics-scale mediation analysis. We introduce ICONIC, an R package that embeds genetic instruments and negative controls within a proximal causal inference framework for total-effect and mediation analysis. ICONIC implements eight estimators spanning five confounding-control strategies, supports continuous, binary, and time-to-event outcomes, and provides extensive diagnostics including sensitivity analyses that map estimator performance across plausible assumptions. Ground-truth benchmarks are calibrated to real-omics covariance structures via a hybrid generative model (GAN + feature-level Gaussian copula) rather than parametric simulation, and a companion planning tool predicts performance gains from collecting additional omic data. We demonstrate ICONIC in two case studies: screening for placental transcriptomic mediators of gestational diabetes on birth weight (n = 164), and tumor-expression mediators of smoking intensity on lung cancer survival (n = 494). Notably, ICONIC’s diagnostics recommended different estimation strategies across the two scenarios, reflecting differences in the likely influence of unmeasured confounding. ICONIC is freely available at https://github.com/sbresnahan/iconic/.

## Introduction

Non-experimental multi-omic studies of human cohorts have generated catalogs of molecular features associated with exposures and health outcomes.^1–4^ These catalogs reveal that genetic effects on molecular traits are themselves context-dependent, varying across social, behavioral, and nutritional co-exposures in cellular and molecular environments (**Figure 1A**).^5–8^ Yet few studies have applied formal causal inference methods to address these complex confounding environments, typically accounting for only a few covariates or technical controls.^9^ However, unmeasured confounders such as socioeconomic status, diet, environmental toxicants, hormonal environment, and/or genetic interactions/epistasis vastly outnumber included controls^10,11^ and systematically bias effect estimates. Importantly, these biases do not shrink with sample size and, therefore, threaten large, high-powered biobank studies.^12,13^ Accordingly, genome-, epigenome-, and transcriptome-wide association (GWAS, EWAS, TWAS) hits suffer from pronounced test-statistic inflation and bias that standard confounder adjustments cannot fully correct.^14,15^

**Figure 1.**
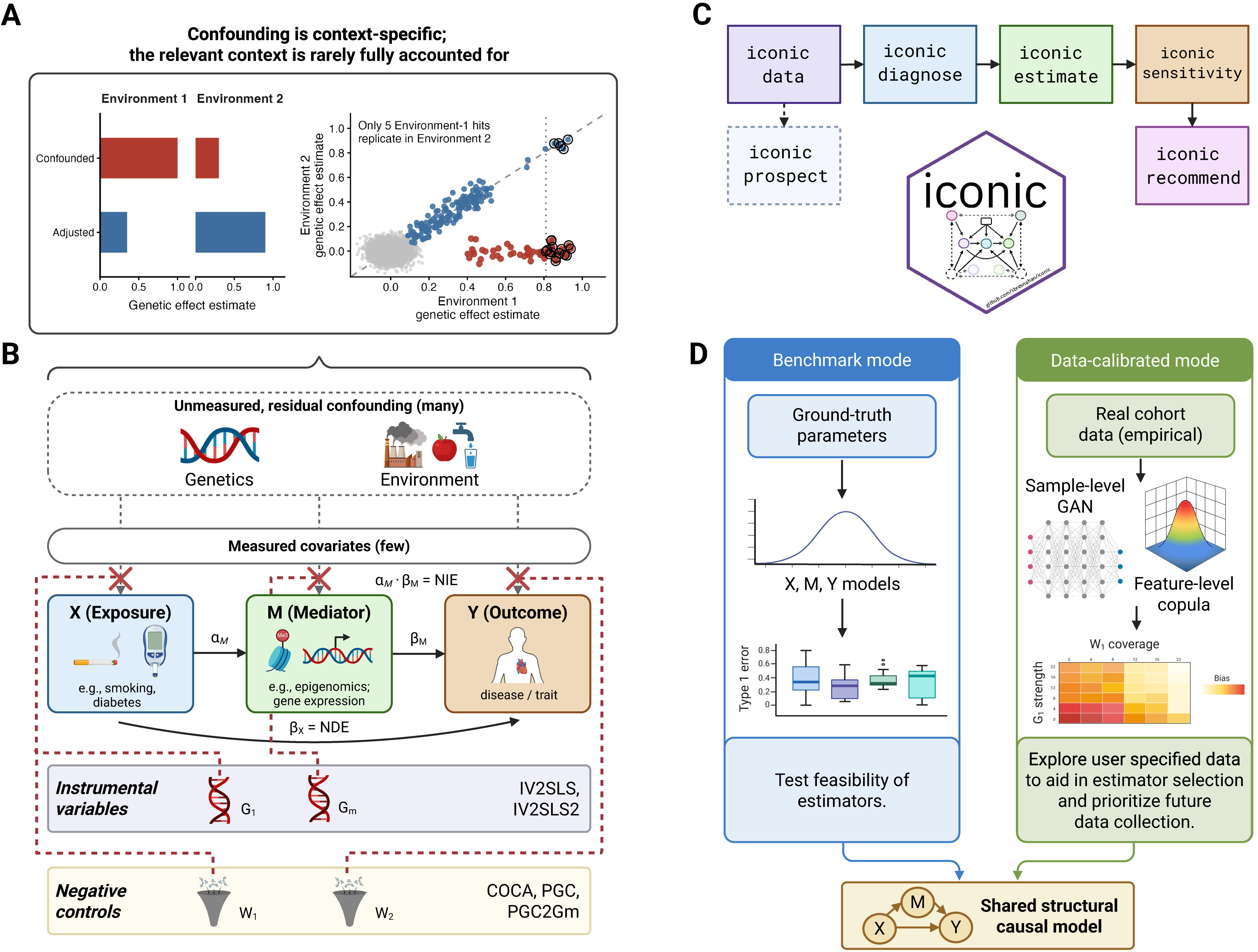
The ICONIC framework. **(A)** Left: for a single genetic variant, the unadjusted effect estimate (red) versus the estimate adjusted for the relevant environmental context (blue), in two environments. Unmeasured environmental contributions inflate the estimate in Environment 1 but mask it in Environment 2. Right: Projected to the omics-wide case (illustrative simulation of 5,000 features): genetic effect estimates in Environment 1 versus Environment 2. Unconfounded effects (blue) lie on the 1:1 diagonal and replicate; associations driven by Environment-1-specific confounding (red) are strong in discovery but absent in replication. Open circles mark hits above a study-wide effect estimate threshold. **(B)** Unmeasured confounders *U* bias the exposure-mediator and mediator-outcome paths, corrupting both the direct and indirect effect. Three routes to identification: genetic instruments (Mendelian randomization, e.g. implemented by estimators IV2SLS and IV2SLS2), negative controls (proximal inference, e.g. implemented by estimators COCA, PGC, and PGC2Gm), and their combination, are used to block the residual confounding. **(C)** The ICONIC workflow. **(D)** Two simulation modes (structural benchmark and data-calibrated generative texture). *Abbreviations: NDE (natural direct effect); NIE (natural indirect effect); GAN (generative adversarial network)*.

These problems are then compounded when attempting to estimate molecular mediators of effects, a common objective in molecular discovery studies: unmeasured confounding will separately threaten exposure-mediator, mediator-outcome, and exposure-outcome estimates. Formal quantitative causal inference approaches are well positioned to close this gap in molecular discovery: Such analyses provide a framework within which prevailing multi-omic data coupled with foundational biological knowledge can be used to explicitly control unmeasured confounding.

Two methodological traditions in formal quantitative causal inference have been proposed to address unmeasured confounding in -omics studies through the use of accessory variables (**Figure 1B**). Mendelian randomization (MR) uses genetic variants as instrumental variables, exploiting conditionally random allele allocation at conception to estimate exposure effects through a channel independent of unmeasured confounding.^16,17^ Its validity rests on untestable assumptions including random allocation, monotonic exposure effects, and no unaddressed pleiotropy. A complementary approach, less commonly used in genomics, is the use of independent, negative control (NC) outcome variables for effect calibration.^18^ NC outcomes are assumed to be causally unaffected by the exposure but share the same set of unmeasured confounders. If these conditions are fulfilled, the NC outcome itself can be used as a proxy variable to directly capture and correct for the biasing effect of the unmeasured confounders.^10,11,13^ In summary, these two methods rest on opposing identifying assumptions and address confounding by complementary strategies: genotype-exposure dependence used to “ignore” unmeasured confounding (MR) vs. exposure-control independence to capture it (NC). Most recently, proximal g-computation (PGC) synthesizes the two approaches, using an instrument to isolate the confounded component of the exposure and negative controls to absorb that component within a single estimator. Importantly, this proximal causal inference framework is potentially more robust than either approach alone by allowing the strengths of each method to make up for residual biases of the other.^12,13,19,20^

Mediation analysis is a natural extension of proximal causal inference for genomics, decomposing exposure effects into indirect effects through a molecular intermediate and the remaining direct effect. However, mediation introduces three sources of unmeasured confounding (exposure-mediator, mediator-outcome, and exposure-outcome). Of these, mediator-outcome confounding is the most damaging as it biases both indirect and direct effect estimates. It is present even in randomized designs, since molecular mediators share post-treatment genetic and environmental drivers with the outcome.^21^ In practice, mediator-outcome confounders are rarely addressed explicitly.^22^ Recent work has extended proximal causal identification to mediation using negative-control proxies at each stage of the mediation path,^23,24^ but these advances have not been integrated with genetic instruments in accessible software or validated against real-world genomic data structures.

Existing software packages implement various causal inference strategies in isolation: MR platforms such as TwoSampleMR and MR-Base^25^ operate on GWAS summary statistics for exposure-outcome effects, summary-data-based methods such as SMR^26^ link GWAS to eQTL summary data for gene prioritization, the PCL^27^ R package implements proximal causal learning for total effects, and CMAverse^28^ provides full causal mediation analysis but assumes no unmeasured confounding. None combine genetic instruments and negative controls, and none develop proximal causal inference estimators for mediation under unmeasured mediator-outcome confounding. **Table 1** compares the capabilities of ICONIC against these existing tools.

**Table 1.**
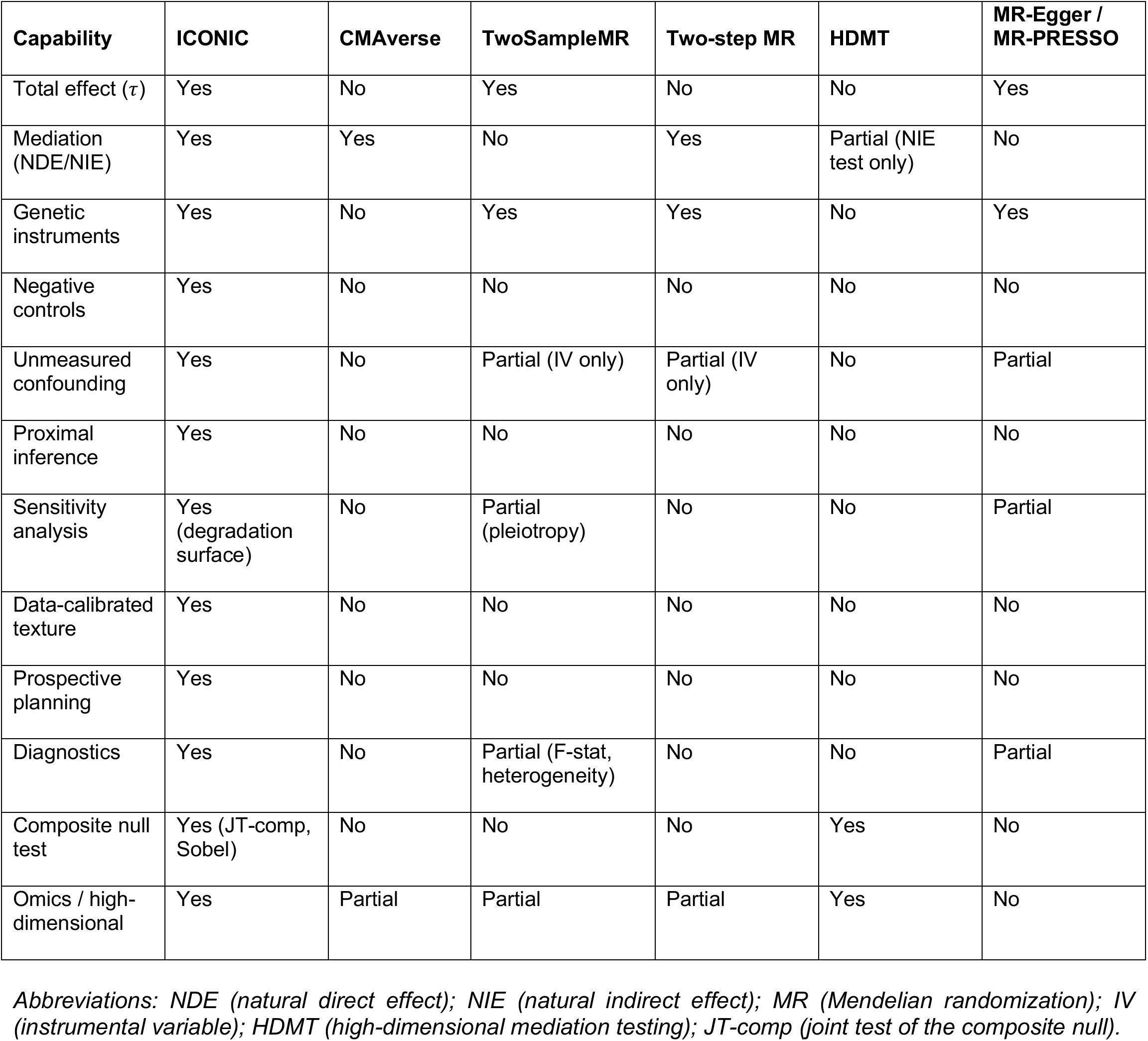
Feature comparison of ICONIC against existing tools.

| Capability | ICONIC | CMAverse | TwoSampleMR | Two-step MR | HDMT | MR-Egger / MR-PRESSO |
| --- | --- | --- | --- | --- | --- | --- |
| Total effect ($\tau$) | Yes | No | Yes | No | No | Yes |
| Mediation (NDE/NIE) | Yes | Yes | No | Yes | Partial (NIE test only) | No |
| Genetic instruments | Yes | No | Yes | Yes | No | Yes |
| Negative controls | Yes | No | No | No | No | No |
| Unmeasured confounding | Yes | No | Partial (IV only) | Partial (IV only) | No | Partial |
| Proximal inference | Yes | No | No | No | No | No |
| Sensitivity analysis | Yes (degradation surface) | No | Partial (pleiotropy) | No | No | Partial |
| Data-calibrated texture | Yes | No | No | No | No | No |
| Prospective planning | Yes | No | No | No | No | No |
| Diagnostics | Yes | No | Partial (F-stat, heterogeneity) | No | No | Partial |
| Composite null test | Yes (JT-comp, Sobel) | No | No | No | Yes | No |
| Omics / high-dimensional | Yes | Partial | Partial | Partial | Yes | No |

*Abbreviations: NDE (natural direct effect); NIE (natural indirect effect); MR (Mendelian randomization); IV (instrumental variable); HDMT (high-dimensional mediation testing); JT-comp (joint test of the composite null).*

We developed ICONIC (Inference with Causal Observational Negative-control Instruments and Controls), an R package that implements five conceptually distinct causal inference approaches, extended to eight estimators when mediation-specific variants are included. ICONIC separates estimator validation from data-calibrated sensitivity analysis. The estimators are first validated on structural synthetic data with known ground truth, and then a generative texture model trained on the user’s own data calibrates the sensitivity analysis to realistic covariate, outcome, and mediator distributions. The texture model learns a nuisance structure (feature marginals, covariate structure, and correlation patterns) and the causal relationships among exposure, mediator, and outcome in the synthetic data are imposed by a structural skeleton with investigator-specified effects, so the ground-truth effect is guaranteed by construction. Any study with an exposure, a molecular mediator panel, an outcome, and at least one of the two identifying resources (genetic instruments or negative controls) can use ICONIC to diagnose estimator validity, to benchmark performance under plausible confounding scenarios calibrated to the user’s own data, and to identify the most robust causal estimate (**Figure 1C-D**).

## Results

### Overview of the ICONIC framework

We begin with the causal concepts before describing the software, since the workflow is organized around them. All estimators are evaluated against a structural causal model (SCM) in which an exposure *X* affects an outcome *Y* both directly and indirectly through a mediator *M* (**Figure 2**). The direct effect (natural direct effect, NDE) is the effect of *X* on *Y* that does not pass through *M*; the indirect effect (natural indirect effect, NIE) is the effect that operates through *M*; and the total effect is their sum. The model includes genetic instruments *G*_1_ (for *X*) and *G*_2_ (for *M*), a negative-control panel *W*, and a *k*-dimensional vector of unmeasured confounders *U* whose path-specific loadings confound the *X* → *M* and *M* → *Y* paths. Two quantities govern how strongly the identifying assumptions can be violated before the estimators cease to be valid. The first is the correlation between each genetic instrument and its corresponding unmeasured confounder (*ρ_G_*_1_ and *ρ_G_*_2_); the second is the negative-control coverage *ω*, the fraction of each path’s confounder composite captured by the control panel. Because the confounders are unobserved, neither quantity can be estimated from data. ICONIC therefore treats them as unknowns to be stress-tested through sensitivity analysis.

**Figure 2.**
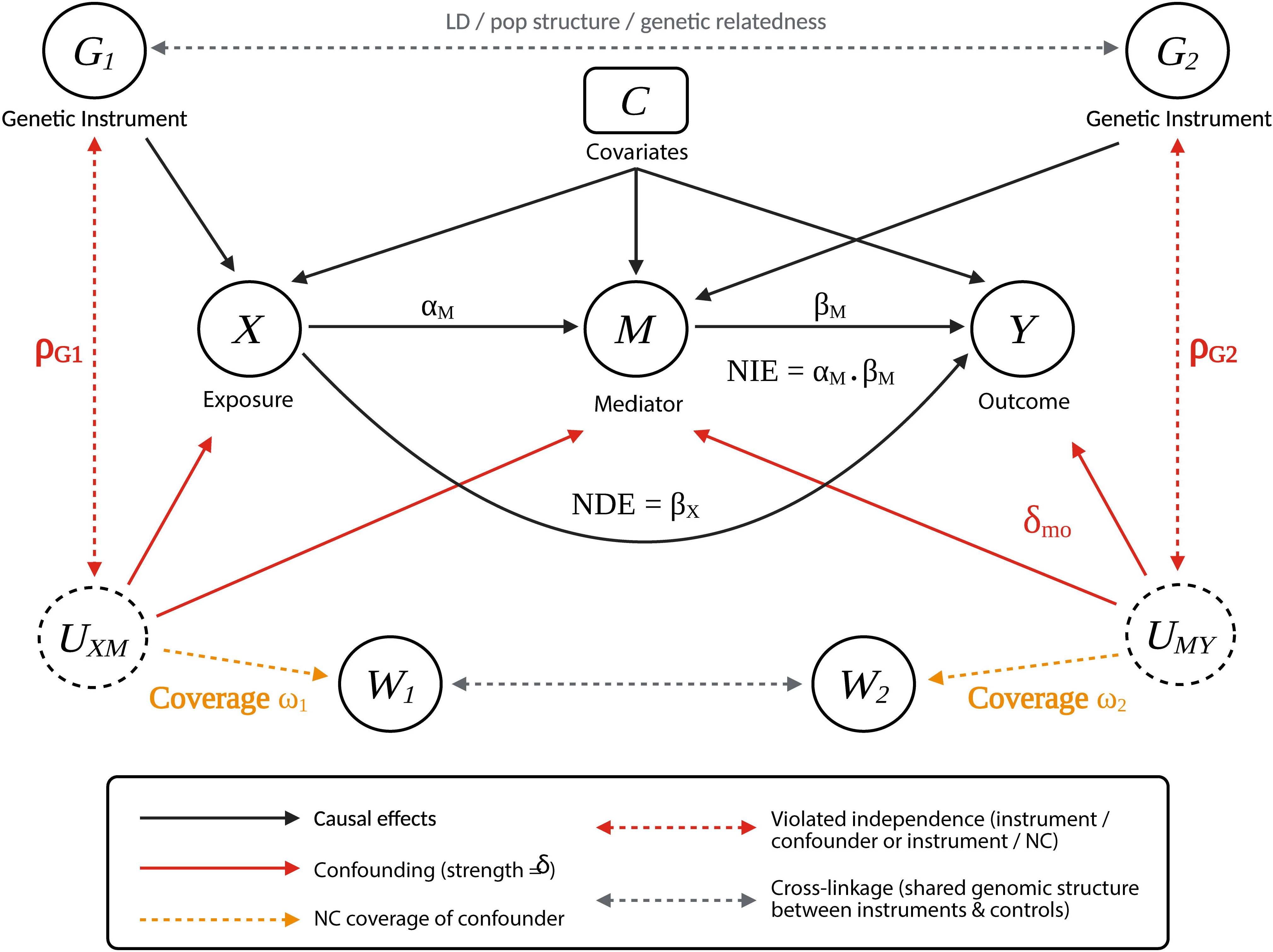
Causal directed acyclic graph for the ICONIC mediation framework. Exposure *X* affects mediator *M* (path *α_M_*) and outcome *Y* both directly (*β_X_* = *NDE*) and indirectly through *M* (path *β_M_*). Genetic instruments *G*_1_ and *G*_2_ instrument *X* and *M* respectively; both are assumed independent of unmeasured confounders and to affect *Y* only through their targets, though linkage or population structure can induce cross- linkage (gray dashed arrow). Unmeasured confounders *U* bias the *X* → *M* and *M* → *Y* paths (red solid arrows). A negative-control panel *W* proxies the confounder composites with coverage *ω*_1_ and *ω*_2_ (orange dotted arrows). Red dashed arrows denote violations: *ρ_G_*_1_ and *ρ_G_*_2_ represent instrument-confounder correlation, and *δ_XM_* / *δ_MY_* denote the confounding paths. Covariates *C* affect *X*, *M*, and *Y* throughout. *Abbreviations: NDE (natural direct effect); NIE (natural indirect effect); LD (linkage disequilibrium)*.

ICONIC provides an integrated workflow for causal inference in observational omics studies that combines genetic instruments and negative controls within a single proximal causal inference framework. The workflow has four layers (**Figure 1C**):

1. **Diagnose:** `iconic_diagnose()` runs instrument-strength checks, negative-control validity screens (A1, A2, A2’), and a two-component path-completeness assessment, returning an eligibility report for all eight estimators.
2. **Estimate:** `iconic_estimate()` fits all eligible estimators on the user’s real data, returning per-feature and per-mediator NDE/NIE estimates with standard errors and confidence intervals.
3. **Stress-test:** `iconic_sensitivity()` varies the untestable quantities introduced above, the instrument-confounder correlations *ρ_G_*_1_ and *ρ_G_*_2_ and the negative-control coverage *ω*, across a plausible range and records how each estimator’s bias changes. We call the resulting map a “degradation surface”, and it is calibrated to the user’s cohort by the generative texture model.
4. **Recommend:** `iconic_recommend()` ranks estimators by per-estimand robustness over that surface, selecting the most robust estimator. A fifth function, `iconic_prospect()`, serves users who do not yet have instruments or negative controls, simulating best-case instruments and controls calibrated to the user’s data and predicting how much the estimate would improve upon collecting them.

ICONIC implements five estimators for the total effect and three additional estimators for mediation (**Table 2**). All estimators except COCA extend to time-to-event (survival) outcomes via two-stage predictor substitution, in which the first-stage regressions remain ordinary least squares (OLS) and only the outcome stage switches to a Cox model (log-hazard-ratio scale) or OLS on restricted mean survival time pseudo-observations, and analogously to binary outcomes, where the outcome stage becomes a logistic regression (log-odds-ratio scale) or a linear probability model (risk-difference scale) **(Supplementary Methods S1 and S13)**.

**Table 2.** ICONIC estimators.

| Estimator | Estimand | Strategy | Identifying assumption | Limitation |
| --- | --- | --- | --- | --- |
| UNADJ | Total effect | Regress $Y$ on $X$ alone | None (reference) | Biased by confounding; bias does not shrink with $n$ |
| DIRECT | Total effect | Regress $Y$ on $X, G, W$, covariates $C$ | All confounders measured | Structural bias from unmeasured confounding; does not shrink with $n$ |
| COCA | Total effect | Regress $W$ on $Y + X$; recover $\hat{\tau} = -\hat{\beta}_X / \hat{\beta}_Y$ | $W$ shares $U$ with $Y$; $W \perp X \parallel C, U$; completeness $\dim(W_{\text{valid}}) \geq k$ | Unstable when denominator $\approx 0$; requires $\geq k$ valid controls |
| IV2SLS | Total effect | Instrument $X$ with $G$ via two-stage least squares | $G \rightarrow X$ only; $G \perp U$; $G \rightarrow Y$ only through $X$ | Requires strong instrument ($F \geq 10$); cannot address $M$ endogeneity in mediation |
| PGC | Total effect | Residualize $X$ on $G$; bridge full $W$ matrix onto residual; include $\hat{W}$ in outcome model | $W$ shares $U$ with $Y$; bridge function correctly specified | $\sim 2 \times$ variance of IV2SLS; requires $\dim(W_{\text{valid}}) \geq k$ (proximal completeness) |
| IV2SLS2 | Mediation | 2-stage MR: instrument $X$ with $G$, instrument $M$ with $G_m$ via sequential 2SLS; optional path-specific NC augmentation ($W_1$ stage 1, $W_2$ stages 2-3) | $G$ valid for $X$; $G_m$ valid for $M$; both strong (partial $F \geq 10$) | Requires a mediator-specific instrument; analysis restricted to mediators with identified instruments; biased if $G_m$ correlated with the $M \rightarrow Y$ confounder composite; when NC-augmented, higher coverage improves (not degrades) the estimate |
| PGC2 | Mediation | Two-stage proximal mediation: bridge $W$ with $X$ (purge the $X \rightarrow M$ composite), bridge $W$ with $M$ (purge the $M \rightarrow Y$ composite), no $G_m$ required | $W$ spans the $X \rightarrow M$ composite (completeness at stage 1); $W$ spans the $M \rightarrow Y$ composite (completeness at stage 2); bridge functions correctly specified at both stages | Completeness must hold at both stages; biased when $W$ coverage of either composite is low |
| PGC2Gm | Mediation | NC-augmented: bridge $W$ for $X$, use $G_m$ to isolate the $M \rightarrow Y$ composite's effect on $M$, bridge $W$ to remove it | $W$ spans the $X \rightarrow M$ composite; $W$ spans the $M \rightarrow Y$ composite; $G_m$ relevant for $M$ | Requires both adequate $W$ coverage and a mediator instrument; residual bias from $G_m$ correlated with the $M \rightarrow Y$ composite is smaller than IV2SLS2 but nonzero |

Abbreviations: MR (Mendelian randomization); 2SLS (two-stage least squares); NC (negative control).

A design principle of ICONIC is that it separates estimator validation from data-calibrated sensitivity analysis. The estimators are first validated on structural synthetic data with known ground truth. A generative texture model (a hybrid of a sample-level GAN and a feature-level Gaussian copula) then trains on the user’s own data so that the sensitivity and prospective analyses are calibrated to realistic covariate, outcome, and mediator distributions rather than to generic assumptions. Throughout, we use “texture” to refer to the nuisance structure of the data (feature marginals and correlation structure), as distinct from the causal parameters under study.

### Implementation validation and expected bias structure

We first confirm that ICONIC’s estimators reproduce the bias structure predicted by published theory. These benchmarks use the structural synthetic-data generator (`generate_toy_data`) with parametric Gaussian noise, so the ground truth is known exactly **(Supplementary Figures S1-S2)**. For total-effect estimation, IV2SLS was unbiased across all confounding strengths under instrument validity, with calibrated Type I error, and its bias increased proportionally to pleiotropy strength **(Supplementary Figure S3)** and rose sharply once the first-stage partial *F*-statistic fell below the conventional weak-instrument threshold of 10 **(Supplementary Figure S4)**. For mediation, estimators without a mediator-specific instrument carried structural, non-diminishing bias for both NDE and NIE, whereas IV2SLS2 reduced bias sharply and PGC2Gm tracked it closely, with the negative-control coverage of each path’s confounder composite further modulating bias **(Supplementary Figure S5)**. NIE Type I error under the null was well-controlled by IV2SLS2 and PGC2Gm.^13,22,23,29^

### Mapping untestable violations via the degradation surface yields a data-texture-dependent estimator ranking

The instrument-exogeneity parameters *ρ_G_*_1_ and *ρ_G_*_2_, the correlations between each genetic instrument and its corresponding unmeasured confounder, cannot be estimated from data because the confounders are unobserved. The same is true of the negative-control coverage *ω*, the fraction of each path’s confounder composite captured by the control panel. The standard approach in MR and proximal causal inference asserts that the instrument correlations are zero and the coverage complete. ICONIC instead maps the full *ρ_G_*_1_ × *ρ_G_*_2_ plane jointly across a sweep of *ω* and reports where each estimator ceases to be valid.

The degradation surface (**Figure 3**) reports each estimator’s NDE and NIE bias across the full *ρ_G_*_1_ × *ρ_G_*_2_ plane at each level of negative-control coverage *ω*, demonstrating how the two estimator families degrade along complementary axes. Instrument-confounder correlation drives bias in the IV estimators while leaving the negative-control estimators largely unaffected, and weakening coverage does the reverse. IV2SLS (total-effect) is coverage-inert, whereas IV2SLS2, when augmented with path-specific negative controls, responds to coverage in the improving direction (higher *ω* reduces its NDE bias), so its coverage gradient is opposite in sign to the degradation the NC estimators show as coverage thins. Sweeping *ω* jointly with *ρ_G_*_1_ and *ρ_G_*_2_ therefore exposes the tradeoff between the two identification strategies rather than treating either as the ground truth. Two summaries annotate the surface. Tipping points record the smallest violation at which an estimator’s absolute bias crosses a user-defined threshold, defined marginally (the first grid value of *ρ_G_*_2_ holding *ρ_G_*_1_ = 0 at which absolute bias crosses the threshold, and symmetrically for *ρ_G_*_1_). Worst-case bias reports the maximum absolute bias over all parameter configurations. Together these allow the analyst to delimit the regime in which each estimator can be trusted and to select the estimator most robust under the violations considered plausible. The surface is calibrated to the user’s data, with sample size, instrument strength, confounding magnitude, and covariate and outcome texture all drawn from the analyst’s dataset and its diagnostic summaries.

**Figure 3.**
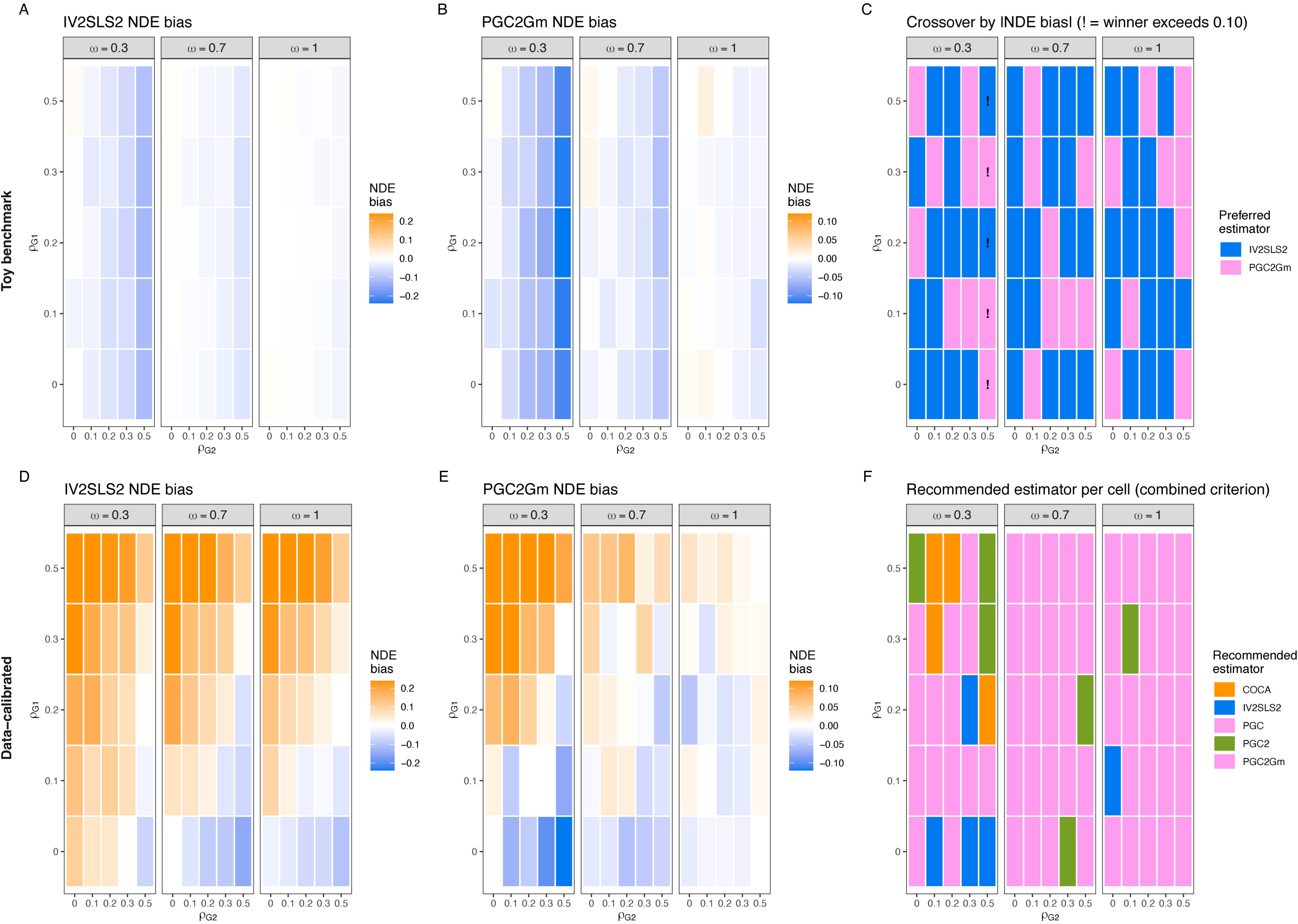
Estimator bias under simultaneous instrument-exogeneity violations, under toy and data- calibrated texture, visualized by the degradation surface. **(A)**-**(C)** Gaussian toy benchmark. **(A)** IV2SLS2 NDE bias and **(B)** PGC2Gm NDE bias across a 5 × 5 grid of *ρ_G_*_1_ × *ρ_G_*_2_, each over {0, 0.1, 0.2, 0.3, 0.5}, faceted by coverage (*ω* ∈ {0.3,0.7,1.0}). Color scale centered at zero (blue = negative, orange = positive). **(C)** Crossover map showing which estimator has lower |NDE bias| at each configuration (blue = IV2SLS2, pink = PGC2Gm); “!” marks configurations where the preferred estimator exceeds |bias| = 0.10. Settings: *n* = 500, 50 replicates per configuration, 10 features, `mo_confounding` = 0.8, *ϕ* = 0.8, *k* = 2, *ω*_1_ = *ω*_2_ = 0.7. **(D)**-**(F)** Data-calibrated surface from `iconic_sensitivity()`, calibrated to a simulated example dataset (200 samples, 1 outcome feature, 1 mediator, 20 replicates per configuration) via the generative texture model (GAN + copula). **(D)**-**(E)** NDE bias for IV2SLS2 and PGC2Gm across the same grid, faceted by coverage. **(F)** The recommended estimator at each configuration, faceted by *ω*. Neither *ρ_G_*_1_ nor *ρ_G_*_2_ nor *ω* can be estimated from data; the surface maps the plausible range calibrated to the user’s cohort. *Abbreviations: NDE (natural direct effect); NIE (natural indirect effect)*.

Under a generic Gaussian benchmark (**Figure 3A-C**), the surface yields no stable ranking. Gridding both instrument-exogeneity violations simultaneously (*ρ_G_*_1_ and *ρ_G_*_2_, each over {0,0.1,0.2,0.3,0.5}) and faceting by negative-control coverage (*ω* ∈ {0.3,0.7,1.0}), both leading estimators were near-unbiased at the origin and degraded only modestly across this grid (max |NDE bias| = 0.121 for IV2SLS2 and 0.123 for PGC2Gm). Both estimators improve as coverage *ω* rises. IV2SLS2’s worst-configuration NDE bias fell from −0.121 at *ω* = 0.3 to −0.054 at *ω* = 0.7 and −0.023 at *ω* = 1.0, with PGC2Gm tracking closely (−0.123, −0.053, and −0.029 respectively). The crossover map (**Figure 3C**) splits the grid between the two estimators with no stable winner. IV2SLS2 is preferred in 48 of 75 parameter configurations (15/25, 17/25, and 16/25 at *ω* = 0.3, 0.7, and 1.0), a near-even split consistent with Monte Carlo noise. A generic benchmark thus leaves the analyst indifferent between the two estimators.

Under data texture learned from real omics cohorts (**Figure 3D-F**), a clear and coverage-dependent ordering emerges. We applied the full model-selection workflow to a simulated example dataset with the generative texture model trained from the example data itself. The degradation surface under learned texture, faceted by coverage (*ω* ∈ {0.3,0.7,1.0}), shows that PGC2Gm has the lower |NDE bias| at the origin and low-violation parameter configurations when coverage is high, while IV2SLS2 degrades more gracefully at the high-violation configurations. The recommended-estimator map (**Figure 3F**) shows the recommended estimator at each configuration shifting toward PGC2Gm as *ω* strengthens, with PGC2Gm being the top estimator in approximately 60% of configurations at *ω* = 0.3, rising to 88% at *ω* = 0.7 and 88% at *ω* = 1.0. Yet the overall recommendation under the combined criterion is IV2SLS2 (final score 0.821 vs 0.658), because PGC2Gm’s strong per-configuration robustness is discounted by its completeness penalty: its identification rests on a bridge-function completeness condition that the diagnostics can only partially verify, whereas IV2SLS2’s instrument-based identification carries no such penalty. Because PGC2Gm leans on the negative-control bridge for identification, it benefits most from rising coverage, whereas IV2SLS2’s identifying resource is primarily its instruments, with coverage only augmenting its stage-2/3 confounder control.

### Diagnostics separate testable from untestable assumptions

The empirically testable projections concern the observable structure of the negative controls: A1 (exposure independence; *W* ⊥ *X* ∣ *C*, *U*), A2 (instrument independence; *W* ⊥ *G*_1_ ∣ *C*, *U*), A2’ (mediator-instrument independence; *W* ⊥ *G_m_* ∣ *C*, *U*), and A3 (completeness, operationalized as a two-component condition combining a dimensional check with a covariance-capture test). These are projections of the underlying identifying assumptions onto observable quantities. Because *U* is unobserved, the conditional independencies cannot be tested directly, but each induces a testable implication in the observed data. We empirically validated these screens across controlled violation scenarios (*n* = 500, 50 replications per sweep; **Supplementary Figure S6**). The A1, A2, and A2’ screens detected injected violations at low strength while keeping clean controls below a low false positive rate. For A3, both data-driven components of the completeness assessment tracked true negative-control coverage across a sweep of *ω*. Covariance-capture *R*^2^ rose monotonically with coverage against a calibrated permutation null, and the support diagnostic *R*^2^(*Ũ* ∣ *W*) progressed from narrow to broad coverage **(Supplementary Figure S6D-E)**. A2 and A2’ violations arising from instrument-confounder correlation (rather than direct instrument-control paths) are not detectable by the empirical screens, and the true coverage of the confounder space is likewise unobservable.

### Generalizability and robustness to data structure violations

We examined whether the benchmark conclusions from the idealized simulations transfer to the structured data encountered in real omics studies, probing two structural violations ubiquitous in molecular data: feature-level correlation structure (co-expression modules) and heterogeneity in confounder strength and negative-control coverage.

#### Feature-level correlation structure

We swept the within-module correlation (`feat_cor`) from 0 (independent) to 0.8 (strong correlation), using a block-diagonal correlation matrix with ⌈√*p*⌉ modules injected into the outcome (*Y*) and negative-control (*W*) noise panels, i.e., the *W*-*Y* residual correlation structure **(Supplementary Figure S7)**. Estimator performance was invariant to this correlation strength. NDE and NIE bias, RMSE, NIE Type I error under the null, the diagnostic screens, the model-selection recommendation, and the worst-case bias on the degradation surface **(Supplementary Figure S8)** were all unchanged across the full sweep.

This invariance is specific to imposed block-diagonal *W*-*Y* structure, however, and does not extend to the full data texture emulated by the generative model trained on the user’s own data. Under the learned structure, the two leading estimators reverse, with PGC2Gm’s worst-case NDE bias rising above IV2SLS2’s **(Supplementary Figure S8).**

#### Heterogeneity in confounder strength and negative-control coverage

We probed heterogeneity via two parameters, `u_strength` (a per-confounder vector scaling each confounder’s contribution) and `w_coverage_profile` (a per-control coverage list assigning each *W* feature its own capture of *U*) **(Supplementary Figure S9)**. Under three *W* coverage profiles (uniform, moderate, sparse), covariance-capture *R*^2^ remained strong (above the 0.3 threshold) in all cases, complementing the coverage-level sweep of the same diagnostic (**Supplementary Figure S6D**). Under three *U* strength profiles (uniform, moderate, extreme), NDE bias for the two leading estimators was largely invariant to the profile when each path’s confounder composite was distinctly loaded (*λ_XM_* = *e*_1_, *λ_MY_* = *e*_2_).

### Prospective analysis predicts the value of collecting instruments

For users who have exposure, mediator, and outcome data but no genetic instruments or negative controls, ICONIC provides a prospective analysis that predicts how much the estimate would change upon collecting an instrument or negative controls, and how strong the instrument would need to be. This tool has no analogue in existing MR or proximal-inference software.

The analysis proceeds in three phases (**Figure 4**). Phase 1 sweeps instrument strength (*γ_G_*) to show how estimates converge to the true effect, contrasting the biased UNADJ estimate against the instrument-based and negative-control estimators. Phase 2 runs a full prospective simulation at a target instrument strength, generating synthetic instruments and negative controls calibrated to the user’s sample size and confounding level, and reports the expected NDE and NIE estimates along with first-stage *F*-statistics. Phase 3 sweeps instrument-exogeneity violations (*ρ_G_*_1_ × *ρ_G_*_2_) jointly with negative-control coverage (*ω*) at the target strength, quantifying how each estimator degrades as instruments become imperfect and controls weaken. The coverage axis of this surface under perfect instrument exogeneity is shown directly in **Figure 4D**. COCA and PGC2 were unstable across this coverage sweep (COCA converged in only 12-17 of 20 replicates, and PGC2 reached an NDE SD of 27) and are omitted from the panel for legibility. This degradation surface feeds a robustness-based recommendation, reported as the best eligible estimator under each collection scenario (instrument only, negative controls only, or both), so the recommendation reflects performance under realistic imperfection rather than a single best-case point. On an example dataset under strong mediator-outcome confounding, Phase 1 showed all three instrument-using estimators converging toward zero bias as instrument strength increases, and Phase 2 yielded first-stage partial *F*-statistics well above the *F* ≥ 10 weak-instrument threshold (*F* = 183.2 for the exposure instrument, *F* = 122.4 for the mediator instrument).

**Figure 4.**
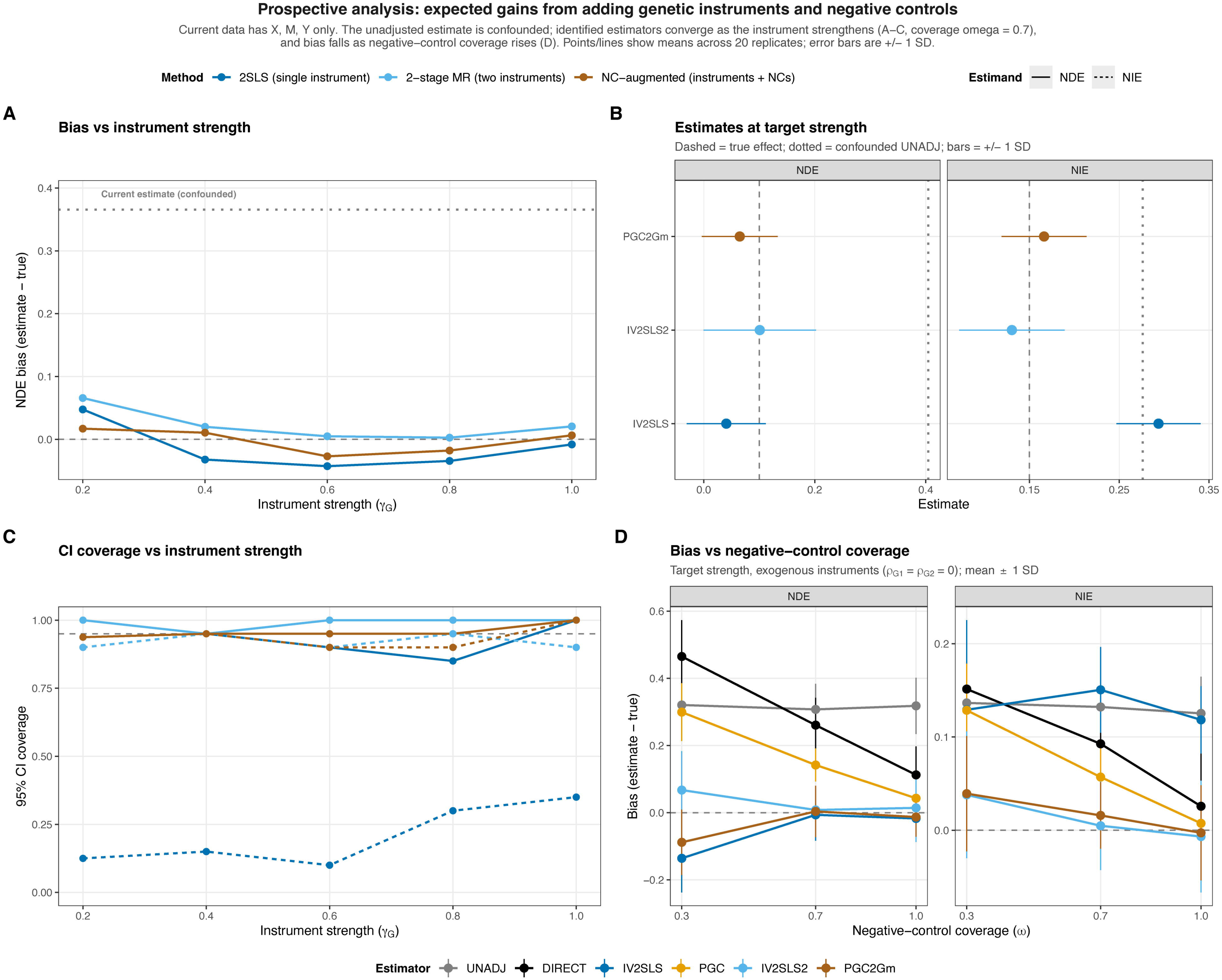
Prospective analysis: expected gains from adding genetic instruments and negative controls. For users with exposure, mediator, and outcome data but no instruments or negative controls, this analysis estimates how much estimates would improve upon collecting them (20 simulation replicates; points and lines show replicate means, and error bars in B and D show ±1 SD). **(A)** NDE bias versus exposure-instrument strength (*γ_G_*) for three instrument-using estimators, compared against the confounded UNADJ estimate (dotted reference). **(B)** Prospective NDE and NIE at the target strength (*γ_G_* = 0.6). Dashed lines: true effects (NDE = 0.10, NIE = 0.15); dotted: confounded UNADJ. **(C)** 95% confidence-interval coverage for NDE and NIE versus instrument strength; the dashed line marks nominal 95% coverage. Panels A-C are evaluated at negative-control coverage *ω* = 0.7. **(D)** NDE and NIE bias versus negative-control coverage *ω* (the fraction of each path’s confounder composite captured by the control panel) at the target strength with perfectly exogenous instruments (*ρ_G_*_1_ = *ρ_G_*_2_ = 0), for six estimators. COCA and PGC2 are omitted for legibility (unstable across the coverage grid). The recommended estimator is chosen by robustness over a Phase 3 sweep. *Abbreviations: NDE (natural direct effect); NIE (natural indirect effect)*.

The prospective tool is validated against reality via a three-arm case study design (below). Arm 2 runs the prospective analysis on data without instruments, and Arm 3 runs the full analysis with real instruments. If the prospective prediction matches the realized analysis, ICONIC functions as both a planning tool (for researchers deciding whether to collect instruments) and an analysis tool (for researchers who have them). Where the real instruments underperform the best-case simulation, the diagnostics flag the violation, and the degradation surface quantifies its impact.

### Benchmarking against existing tools

No existing tool combines genetic instruments and negative controls for omics mediation, so the benchmarking has two goals: 1) to confirm that ICONIC’s individual estimators match or improve upon their isolated counterparts in existing packages, and 2) to demonstrate that ICONIC’s unified framework provides guidance that no existing tool offers. We compared ICONIC’s estimators against their closest analogues in four established packages (TwoSampleMR, CMAverse, two-step MR, HDMT) and benchmarked ICONIC’s pleiotropy grid against MR-Egger and MR-PRESSO. All comparisons use the same structural simulation generator with known ground truth. The feature comparison matrix (**Table 1**) shows that ICONIC subsumes the capabilities of each individual tool while adding proximal inference, the degradation surface, data-calibrated texture, prospective planning, and diagnostics, none of which any existing package provides.

#### Total-effect estimation

ICONIC’s IV2SLS and TwoSampleMR’s IVW are both 2SLS estimators and should produce equivalent results on the same data. Across all confounding-by-sample-size parameter configurations, the absolute bias difference between the two implementations was ≤ 0.024 (median 0.008) **(Supplementary Figure S10)**, with both maintaining Type I error near the nominal 0.05 level and 95% CI coverage near the nominal level. ICONIC’s PGC, which uses negative controls rather than an instrument, was comparably unbiased up to *δ* = 0.8, degrading only slightly at *δ* = 1.0. ICONIC’s 2SLS implementation matches the established IVW estimator, while PGC provides a negative-control-based alternative when instrument validity is in doubt.

#### Mediation estimation

The mediation comparison draws the sharpest contrast between the methods. Under Scenario A (no mediator-outcome confounding, *δ_mo_* = 0), CMAverse’s regression-based and g-computation estimators agreed with ICONIC’s PGC2Gm **(Supplementary Figure S11)**. Under Scenario B (mediator-outcome confounding, *δ_mo_* = 0.8), the methods diverged sharply **(Supplementary Figure S12)**. CMAverse’s NDE and NIE bias grew with confounding, while ICONIC’s PGC2Gm, which uses the negative-control panel and a mediator instrument to purge the *M* → *Y* confounder composite, remained unbiased across all confounding levels. Two-step MR was also unbiased under mediator-outcome confounding but requires valid instruments for both the exposure and the mediator, limiting its applicability to mediators with identified cis-instruments. Under the null, PGC2Gm maintained calibrated Type I error for both NDE and NIE across the full confounding range, while IV2SLS2’s NDE Type I error was inflated by the *G_m_*-*U_MY_* correlation. PGC2Gm was the only estimator simultaneously unbiased, powerful, and calibrated under the null across the full confounding range.

#### Composite-null mediation testing

For testing the natural indirect effect against the composite null (*H*_0_:NDE = 0 or NIE = 0), we compared ICONIC’s joint test (JT-comp) and Sobel test against HDMT (**Supplementary Figure S13**). Under the null with PGC2Gm estimates, JT-comp achieved calibrated Type I error (0.052) with full power (1.000) under the alternative, while the Sobel test was conservative (Type I error 0.000). HDMT applied to the same PGC2Gm estimates was degenerate (Type I error 0.000), its permutation procedure collapsing to a near-zero rejection rate because the dense-signal setting leaves no null features to calibrate the permutation distribution. PGC2Gm was the only estimator for which the composite-null test was both calibrated and powerful.

#### Pleiotropy robustness

We compared ICONIC’s pleiotropy grid against MR-Egger and MR-PRESSO on the same pleiotropy simulation scenarios **(Supplementary Figures S14 and S15)**. All instrument-based estimators were biased by horizontal pleiotropy, with PGC less biased than IV2SLS at every pleiotropy level because the negative-control bridge partially absorbs the pleiotropic pathway. MR-Egger was also biased with inflated Type I error under the null, and MR-PRESSO was severely biased with Type I error of 1.0 at any *π* > 0. No estimator was immune to pleiotropy on the total effect. The mediation decomposition, however, revealed a structural advantage. For PGC2Gm, the NIE was completely immune to pleiotropy on the exposure instrument *G* (NIE Type I error 0.000 at all pleiotropy levels under the null), because the NIE depends on the mediator instrument *G_m_* rather than on *G*. The NDE, which depends on *G*, was biased. IV2SLS2 showed the same NIE immunity but had inflated NDE Type I even at *π* = 0 due to the *G_m_*-*U_MY_* correlation.

#### Survival outcomes

ICONIC extends all estimators except COCA to time-to-event outcomes via two-stage predictor substitution (2SPS) **(Supplementary Methods S1).** We benchmarked the survival estimators on the same structural DGP, converting the linear predictor to time-to-event via an exponential proportional-hazards model with ∼ 60% event rate **(Supplementary Figure S16)**. On the Cox log-HR scale, the results mirrored the continuous benchmark, with IV2SLS near-unbiased, PGC2Gm the best NDE estimator, and PGC the best NIE estimator. On the RMST scale, all estimators showed higher variance from pseudo-observation noise. The Cox log-HR scale is the recommended primary effect scale for survival outcomes. RMST provides a collapsible complement with an exact NDE/NIE decomposition when marginal effects are of interest, at the cost of higher variance.

#### Binary outcomes

ICONIC extends all estimators except COCA to binary outcomes via the same 2SPS construction **(Supplementary Methods S13)**. We benchmarked the binary estimators on the same structural DGP, drawing the 0/1 outcome from the linear predictor via a logistic model calibrated to 50% prevalence **(Supplementary Figure S17)**. The results mirrored the continuous and survival benchmarks. On the log-odds-ratio scale, IV2SLS remained near-unbiased as unmeasured confounding increased (bias 0.02 at the strongest confounding level versus 0.44 for the unadjusted estimator), and its Wald confidence intervals retained near-nominal coverage (0.94) while the unadjusted intervals collapsed (0.04); PGC was intermediate (bias 0.22, coverage 0.66). For mediation, the instrumented estimators IV2SLS2 and PGC2Gm kept natural-effect bias small (log-OR NDE bias −0.03 and −0.02, NIE bias +0.01 for both), whereas the unadjusted estimator was biased in both components (NDE −0.17, NIE +0.15). The risk-difference scale reproduced the same ordering at its smaller effect magnitudes, with unadjusted total-effect bias reaching 0.09 against near-zero bias for IV2SLS. COCA returned NA at all parameter configurations, as expected from its structural incompatibility with binary outcomes. The log-odds-ratio scale is the recommended primary effect scale for binary outcomes. The risk-difference scale provides a collapsible complement with an exact NDE/NIE decomposition when marginal effects are of interest.

### Case study 1 - gestational diabetes, placental gene expression, and birth weight

GUSTO (Growing Up in Singapore Towards healthy Outcomes) is a Singapore prebirth cohort study that recruited women in early pregnancy and followed their children from birth. The question for ICONIC is whether maternal gestational diabetes mellitus affects the child’s birth weight by altering placental gene expression, with maternal 2-hour glucose as the exposure, placental transcripts as mediators, and birth weight as the outcome. We applied a three-way comparison design to the GUSTO case study (*n* = 164 after QC, 5,616 mediator genes, 20 methylation-PC negative controls).

#### Arm 1 (Naive, no instruments or negative controls)

iconic_estimate with covariates only (fetal sex, gestational age, maternal ancestry PCs) yielded the confounded UNADJ estimate, with nominally significant NDE (*p* < 0.05) for 1,892 of 5,616 genes and nominally significant NIE for 336 genes.

#### Arm 2 (Prospective, simulated instruments and controls)

`iconic_prospect` simulated best-case instruments and negative controls. Phase 1 predicted NDE bias convergence from the confounded UNADJ level toward zero as instrument strength increases. Phase 2 projected NDE = 0.213 and NIE = 0.164 under IV2SLS2 at the target strength, against a confounded UNADJ projection of NDE = 0.413 and NIE = 0.262, with strong simulated first-stage *F*-statistics (*F_G_*_1_ = 83.9, *F_Gm_* median = 80.4) (**Figure 5A**). PGC2Gm was projected to be the least biased estimator at the target strength (NDE bias = −0.039, NIE bias = 0.017), followed by IV2SLS2 (NDE bias = 0.113, NIE bias = 0.014).

**Figure 5.**
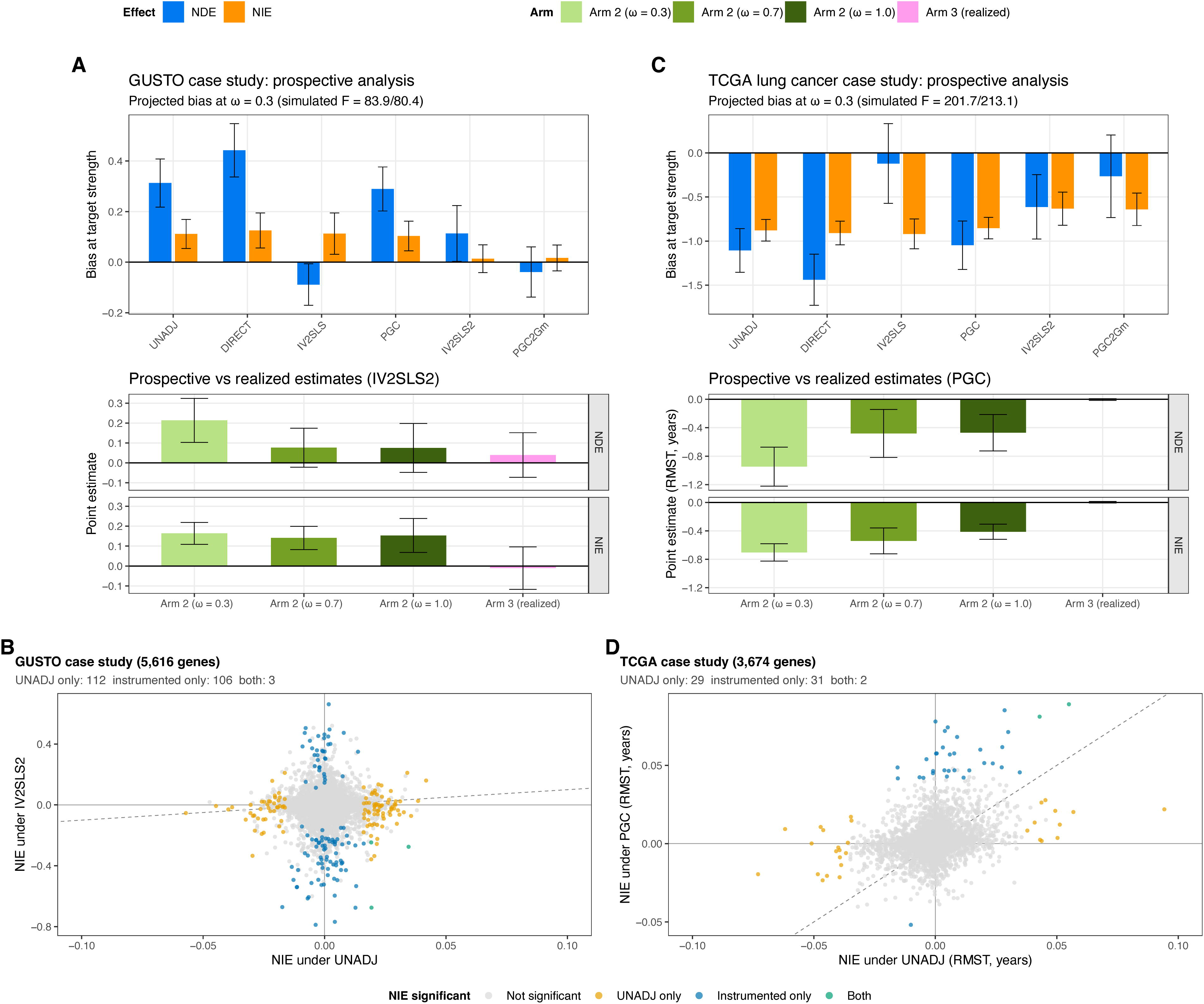
Case studies: prospective projections and genome-wide indirect-effect comparison. **(A)** GUSTO case study, prospective analysis: projected estimator performance at target instrument strength (*γ_G_* = 0.6), comparing best-case simulated instruments (Arm 2) to the realized analysis with real instruments (Arm 3). Top row: NDE and NIE bias at target strength for six estimators (COCA and PGC2 are excluded for instability in these benchmarks). The top row is evaluated at the Phase 2 reference negative-control coverage (*ω* = 0.3). Bottom row: NDE and NIE point estimates under IV2SLS2, with the prospective projection (Arm 2) graded by negative-control coverage. The *ω* = 0.3 bar is the Phase 2 prospective simulation at its reference coverage cell, the *ω* = 0.7 and 1.0 bars are from the Phase 1 instrument-strength surface at the same target strength, and the realized analysis (Arm 3) is shown alongside. Bars show means; error bars are ±1 SD across the 20 simulation replicates (top row and Arm 2) or across per-gene bootstrap point estimates (Arm 3). **(B)** GUSTO case study. Scatter of the natural indirect effect (NIE) under the instrumented IV2SLS2 estimator versus the naive UNADJ estimator for the 5,060 genes tested by both methods. Points are colored by significance class: not significant under either (grey), significant only under UNADJ (orange), only under IV2SLS2 (blue), or under both (green). The x axis is restricted to ±0.1 for visibility; all UNADJ estimates fall within this window. **(C)** TCGA lung cancer case study, prospective analysis (Arm 2 vs. Arm 3, as in A). Top row: NDE and NIE bias at target strength for the same six estimators. Bottom row: NDE and NIE point estimates under PGC (the recommended estimator for this study, consistent with panel D), with Arm 2 graded by negative-control coverage as in (A) versus the realized analysis (Arm 3, per-gene bootstrap means, as in panel D). Error bars are ±1 SD as in (A). Because the mediator-outcome confounding strength could not be inferred for the survival outcome, the prospective simulation used its default setting (*δ_mo_* = 0.8, *ϕ* = 0.8) with user-supplied GAN texture. **(D)** TCGA lung cancer case study. Scatter of the NIE under the recommended PGC estimator versus UNADJ for the 3,674 genes tested by both methods, on the restricted mean survival time (RMST) scale in years. Significance is at bootstrap *p* < 0.05. *Abbreviations: NDE (natural direct effect); NIE (natural indirect effect); RMST (restricted mean survival time*.

#### Arm 3 (Full, real instruments and controls with diagnostics)

`iconic_diagnose` on the full data reported weak exposure instrument strength (*F_G_*_1_ = 5.7, below the *F* ≥ 10 threshold) but adequate mediator instrument strength (*F_Gm_* median = 18.4, 5,385 of 5,616 genes passing *F* ≥ 5). All NC validity screens passed. `iconic_estimate` yielded near-null effects under both leading estimators (IV2SLS2 median NDE = 0.029, median NIE = −0.001; mean NDE = 0.039 and mean NIE = −0.010 across the 5,232 genes), with no gene surviving FDR at 20% for NIE. `iconic_sensitivity` mapped the degradation surface calibrated to the GUSTO data: IV2SLS2 showed moderate degradation (max |NDE bias| = 0.28 within *ρ* ≤ 0.3; 0.36 overall, with tipping at *ρ_G_*_1_ = 0.5), with PGC2Gm similar (0.23 and 0.26) **(Supplementary Figure S18)**. The inferred mediator-outcome confounding strength (*δ_mo_* = 0.066) was well below the *δ_mo_* = 0.8 assumed in the Arm 2 prospective simulation. `iconic_recommend` selected IV2SLS2 (0.758), followed by IV2SLS (0.496) and PGC (0.302). PGC2 and PGC2Gm were ineligible with a single outcome-side panel.

#### Gene-level analysis

**Figure 5B** summarizes the genome-wide results for the 5,060 genes tested by both methods. Of these, 4,839 were not significant under either method, 112 were significant only under the naive UNADJ estimator, 106 only under the instrumented IV2SLS2 estimator, and 3 under both. The genes significant only under UNADJ had small indirect effects (median |NIE| = 0.022), whereas the genes significant only under IV2SLS2 had substantially larger effects (median |NIE| = 0.35), so deconfounding both removed small spurious hits and revealed strong mediation that confounding had masked. **Supplementary Figure S19** shows the top 10 significant mediators per method by NIE (bootstrap confidence interval excluding zero), plotted as the union across UNADJ and IV2SLS2 (25 genes). The two methods identify completely disjoint gene sets. The top 10 UNADJ hits (*GFOD1*, *CARS1*, *NAT14*, *PEX11A*, *PI4K2B*, *ATL3*, *TRGV5*, *AAAS*, *LGMN*, and one unnamed transcript) showed small indirect effects (NIE ≈ −0.06 to +0.04) that vanished under IV2SLS2, consistent with residual mediator-outcome confounding. Conversely, IV2SLS2 hits (including *CDH5*, *LGALS16*, *LAMA5*, *THSD7A*, *GNG11*, *NID2*, *CEBPG*, *LDLR*, *PLIN3*, *STIM1*, *RXRG*, *CTNNBIP1*, *TRIB1*, *PGM1*, *BSG*) showed large indirect effects (NIE ≈ −0.79 to +0.45) that the naive estimator missed entirely.

The IV2SLS2 gene set is biologically coherent with the GDM-placenta-birth-weight pathway, with the strongest hits enriched for vascular and basement-membrane biology. *CDH5*, *LAMA5*, and *NID2* are canonical endothelial and basement-membrane components, and fetoplacental vascular dysfunction is an established consequence of gestational diabetes.^30^ A second cluster implicates placental lipid handling: *LDLR* mediates trophoblast cholesterol uptake^31^,, *TRIB1* regulates *LDLR* expression and is upregulated in fetal endothelial cells in proportion to hyperglycemic exposure,^32,33^ and *PLIN3* coats lipid droplets. Placental expression of *GNG11*, *NID2*, and *CEBPG* has been directly associated with fetal growth,^34^ and maternal diabetes reshapes the placental transcriptome.^35^ The UNADJ hits (*GFOD1*, *CARS1*, *ATL3*, *PI4K2B*) have no established connection to GDM, placental function, or fetal growth, consistent with confounding artifacts.

### Case study 2 - tobacco smoking, lung tumor expression, and lung cancer survival

The Cancer Genome Atlas (TCGA) is a National Cancer Institute consortium that molecularly characterized over ten thousand primary tumors across cancer types. The question for ICONIC is whether tobacco smoking affects survival after a lung cancer diagnosis by altering tumor gene expression, with pack-years as the exposure, tumor transcripts as mediators, and overall survival as the outcome. We applied the same three-way comparison to the TCGA lung cancer case study.

#### Arm 1 (Naive, no instruments or negative controls)

`iconic_estimate` with covariates only (age at diagnosis, sex, histological subtype, tumor stage, ancestry PCs) yielded the confounded UNADJ estimate on the Cox log-hazard-ratio scale, with median NDE = 0.035 (median SE = 1.04) and median NIE = 1.0 × 10^−4^ (median SE = 0.038). Unlike the GUSTO case, where UNADJ was nominally significant for a third of genes, here the naive estimator detected nothing (0 of 3,674 genes reached *p* < 0.05 for either NDE or NIE), because the per-gene log-HR standard errors (≈ 1.0) swamp the small point estimates.

#### Arm 2 (Prospective, simulated instruments and controls)

`iconic_prospect` simulated best-case instruments and negative controls. Phase 1 predicted NDE bias convergence from the confounded UNADJ level toward zero. Phase 2 projected, on the RMST scale, NDE = −0.51 and NIE = −0.48 under IV2SLS2 at the target strength, against a confounded UNADJ projection of NDE = −1.01 and NIE = −0.73, with strong simulated first-stage *F*-statistics (*F_G_*_1_ = 202, *F_Gm_* = 213) (**Figure 5C**). PGC2Gm was projected to be the least biased estimator at the target strength (NDE bias = −0.264, NIE bias = −0.640 years RMST). The corresponding projection under PGC at the reference negative-control coverage (*ω* = 0.3) was NDE = −0.947 and NIE = −0.703 years RMST.

#### Arm 3 (Full, real instruments and controls with diagnostics)

`iconic_diagnose` on the full TCGA data reported an at-threshold exposure instrument (*F_G_*_1_ = 9.6) and adequate mediator instrument strength (*F_Gm_* median = 18.1, with 3,469 of 3,674 transcripts passing *F* ≥ 5). The NC validity screens flagged one of 20 controls under A2 (W-PC 14, partial *r* = 0.17, *p* = 2.5 × 10^−4^, “associated with *G*”), while A1 and A2’ passed all 20 and the two-component completeness check was satisfied (19 valid controls vs *k* = 1), leaving six of eight estimators eligible. As in the GUSTO case study, the two-bridge estimators PGC2 and PGC2Gm were ineligible because only a single outcome-side negative-control panel was available. Because the exposure instrument sits at the weak-instrument threshold, we treat the negative-control estimator (PGC) as the primary analysis and the instrument-based IV2SLS2 estimator as a sensitivity analysis. Under PGC, `iconic_estimate` on the Cox log-HR scale yielded near-null effects (median NDE = 0.013, median NIE = 6 × 10^−5^), with no gene surviving analytic FDR at 20% for NIE. As a sensitivity analysis, the instrument-based IV2SLS2 estimator yielded median NDE = −0.99 and median NIE = −5 × 10^−4^, with 31 genes surviving FDR at 5% and 50 at 20% for the NIE (none for NDE). Given the at-threshold exposure instrument, we treat the PGC analysis as primary. `iconic_sensitivity` mapped the degradation surface using default survival texture. Because the RMST-scale truth is large and negative, all estimators showed substantial absolute NDE bias across the grid (max |NDE bias| within *ρ* ≤ 0.3: 1.07 for IV2SLS2, 0.94 for PGC, 1.15 for UNADJ; overall maxima 1.24 and 1.17 for IV2SLS2 and PGC), and every estimator crossed the 0.1 tipping threshold already at the origin. PGC’s mean |NDE bias| improved with negative-control coverage (0.78, 0.61, and 0.47 at *ω* = 0.3, 0.7, and 1.0), and the surface carries Monte Carlo error of ≈ 0.12 per configuration (10 iterations), so these magnitudes should be read relative to the RMST effect scale rather than as evidence of estimator failure **(Supplementary Figure S20)**. `iconic_recommend` selected PGC (final score 0.409), followed by IV2SLS2 (0.391) and IV2SLS (0.333).

#### Gene-level analysis

**Figure 5D** summarizes the genome-wide results for the 3,674 genes tested by both methods. Of these, 3,612 were not significant under either method, 29 were significant only under the naive UNADJ estimator, 31 only under the instrumented PGC estimator, and 2 under both (significance at bootstrap *p* < 0.05). The genes significant only under UNADJ and those significant only under PGC had comparable effect sizes (median |NIE| = 0.044 vs 0.052 years RMST). **Supplementary Figure S21** shows the top 10 significant mediators per method by NIE (bootstrap confidence interval excluding zero), plotted as the union across UNADJ and the recommended estimator (PGC, labeled “Instrumented”; 20 genes). Arm 3 per-gene bootstrap means under PGC were near null (mean NDE = −0.004, mean NIE = 0.002 years across the 3,674 genes). The top 10 UNADJ hits (*TIRAP*, *LY6K*, *TP53I3*, *STC1*, *NANS*, *SAA1*, *HSPB7*, *KLHL30*, *PIGB*, *CREB3L4*) showed small indirect effects whose intervals all spanned zero under PGC, consistent with confounded naive signal. Under PGC, positive indirect effects emerged whose bootstrap intervals excluded zero (top hits include *CTSL*, *LACTB*, *DECR1*, *SQOR*, *CCR1*, *ALDOC*, *DARS1*, *EPSTI1*, *RGS6*, *TPK1*; NIE ≈ 0.04 to 0.09 years), effects the naive estimator missed entirely. In this case, deconfounding revealed mediation that confounding and the high noise of the naive survival estimator had masked.

The PGC gene set is biologically coherent with the smoking-lung tumor-survival pathway. *CTSL* (cathepsin L) is a lysosomal protease whose high expression in lung adenocarcinoma is associated with advanced stage and poor overall survival.^36^ *LACTB* is a mitochondrial tumor suppressor that is downregulated in lung cancer, with high expression predicting improved survival,^37,38^ and *DECR1*, *SQOR*, and *ALDOC* encode mitochondrial and glycolytic enzymes linked to lung tumor progression and prognosis.^37,39^ *CCR1* is a chemokine receptor whose expression in tumor islets correlates with five-year survival in non-small cell lung cancer,^40^ and *EPSTI1* is an interferon-response gene, consistent with the immune infiltration that smoking shapes in the tumor microenvironment. That smoking’s effect on survival is mediated through tumor gene expression is itself supported by mediation analyses of smoking-related molecular alterations in lung cancer.^41,42^ By contrast, the UNADJ hits (e.g., *TIRAP*, *LY6K*, *SAA1*, *HSPB7*) are dominated by acute-phase and innate-immune markers with no specific connection to smoking-mediated survival, consistent with their interpretation as confounded associations.

## Discussion

Omics mediation estimates are vulnerable to unmeasured confounding at every stage of the mediation path, particularly mediator-outcome confounding, which persists even in experimental designs. MR, negative-control calibration, and proximal causal inference each address complementary aspects of this problem but rest on untestable assumptions whose relative importance varies across datasets. ICONIC addresses this by combining all three within a single tool and providing benchmarks to clarify when each identifying resource is most reliable for a given dataset. Supplying a valid IV for the mediator reduced the bias from unaddressed mediator-outcome confounding, with further improvement from increasingly relevant negative controls. Crucially, these relative advantages shifted with the confounding texture of the data, which is why ICONIC calibrates sensitivity analyses to the analyst’s own dataset rather than offering a universal estimator recommendation.

ICONIC implements diagnostic screens for violations of causal identification assumptions, highlighting potentially invalid IVs and negative controls and flagging effects estimands that do not have sufficient proxy variables before biased downstream estimates are produced. Applying these screens is an important first stage, as blind application of any particular estimator can yield estimates as biased as unadjusted regression. We show invalid instruments introduce pleiotropic bias that does not attenuate with sample size, and negative controls that violate independence or completeness propagate their violation into the bridge-function estimates. Sensitivity to instrument selection is now widely recognized in the MR literature.^43–45^ ICONIC provides a way forward under pleiotropy: The direct/indirect effect decomposition separates pleiotropy-contaminated direct paths from clean indirect paths. The incorporation of a mediator instrument allows the indirect effect to be immune to horizontal pleiotropy of the main exposure instrument. ICONIC’s diagnostic and sensitivity workflow makes such assessments and solutions routine.

This combination of instruments and negative controls also connects two bodies of work that have developed largely in isolation: MR studies have used negative-control outcomes to qualitatively detect residual bias,^17^ whereas ICONIC uses negative-control variable sets within bridge functions to quantitatively correct it, bringing recent proximal identification theory^23,24,46^ into general-purpose software for the first time.

Our benchmarking studies demonstrate the practical advantage of ICONIC for omics mediation analyses. Standard regression- and g-computation-based estimators show marked bias under unmeasured mediator-outcome confounding. Two-step MR remains unbiased but requires valid instruments for both exposure and mediator. ICONIC’s proximal estimators provide an alternative when mediator instruments are unavailable, requiring only sufficient negative controls. When a mediator instrument is also available, PGC2Gm’s direct/indirect decomposition maintains unbiased indirect-effect estimation even under pleiotropy violations that defeat MR-Egger and MR-PRESSO. Overall, ICONIC’s mediation estimators are valid when individual estimator assumptions hold and provide additional robustness in numerous circumstances when they are not. ICONIC also allows analysts to reproduce the behavior of existing tools and then test whether their assumptions hold in a given dataset.

ICONIC’s sensitivity analyses reflect a broader shift toward data-calibrated benchmarking. Method evaluations based on parametric simulations often fail to transfer to real data,^47–50^ and no single simulated dataset can identify a universally dominant estimator. ICONIC addresses this through a hybrid GAN-based texture model that generates confounding scenarios centered on the observed covariance matrix, paired with degradation surfaces that identify which estimator performs best under each scenario. To our knowledge, this is the first software implementation of quantitative, data-adaptive sensitivity analysis for causal mediation.

The two case studies show what this workflow contributes to practice, and how multi-omics data can be readily incorporated into this framework for a wide range of mechanistic discovery applications. They demonstrate two contrasting scenarios in which ICONIC detected the limiting assumptions and recommended the respective analysis which took advantage of the (more) valid accessory -omic data: In the perinatal analysis, the prospective tool identified the likelihood of unmeasured confounding that could benefit from collection/addition of negative control outcomes, obviating the subsequent, invariably biased mediation analyses. Instead, the recognition of valid exposure and mediator IVs enabled application of IV2SLS2 to recover unbiased mediated effects missed by the classic estimator, while simultaneously eliminating false-positive hits. Alternatively, in the lung cancer analysis, negative controls were found to be suitable even in the absence of strong instruments. Consequently, the recommended negative-control estimator (PGC) recovered indirect effects that were otherwise masked. In both studies, a default analysis based on a naive application of any single estimator without ICONICs validity checks would have returned different and misleading gene lists. Moreover, the recommended estimator differed in these cases, illustrating a central claim of this work, that estimator choice should follow dataset-specific diagnostics.

Despite the flexibility and extensibility presented here, we discuss some limitations that persist in our framework. Most fundamentally, MR-based effects are only point identified under both monotonicity and gene-exposure equivalence: the genetic variant must shift exposure in the same direction for everyone in the target population, and that shift must produce the same outcome effect regardless of who experiences it. These assumptions are often, but not always, reasonable. For example, epistasis may make genomic effects inconsistent in different populations. ICONIC does not eliminate the need to interrogate and defend these underlying causal assumptions. Specifically, when using a single instrument for mediation, natural effects under mediator-outcome confounding are bias-reduced approximations rather than point-identified quantities; ICONIC allows for the quantification of this residual bias through the degradation surface. This approximation rests on instrument independence assumptions that are difficult to verify empirically. The analyst must judge the likely magnitude of instrument-confounder correlation from biological context. Negative-control estimators carry their own requirements: causal completeness (coverage of all confounding relationships being proxied) and correct bridge specification at both stages. Violations may escape empirical screens, for example, if the true bridge function is nonlinear, the linear approximation introduces undetectable bias. The two-bridge estimators (PGC2, PGC2Gm) are particularly exposed, since two bridge specifications double the opportunities for misspecification. In both case studies, we used a single negative-control panel for both bridges, accepting potential misspecification as a tradeoff. Where two valid IVs are available and negative-control performance is uncertain, IV2SLS2, which requires no bridge for the mediator-outcome path, may be preferable.

Our sensitivity analyses functions have additional limitations to note. Each `infer_confounding()` estimate carries its own assumptions. The factor model, for example, requires at least five outcome features because parallel analysis infers the confounder count from the eigenvalue spectrum of the residualized outcome correlation matrix. A single-dimensional outcome has no spectrum to analyze, and with fewer than five features, the spectrum is too short for reliable comparison, so the method defaults to one confounder. There is also an inherent circularity: the benchmark ranks estimators using a confounding structure that it inferred itself. Thus, it cannot independently assess estimator validity against the true data-generating process, a limitation inherent to simulation studies generally. The texture model is constrained by GAN instability in small-sample regimes and a Gaussian copula that captures only linear dependence, though vine-copula extensions could relax this restriction.^47^ At the estimator-class level, all estimators are linear, so rankings may not transfer to nonlinear or interaction-rich settings, and the regression-based implementations lack the double robustness of semiparametric proximal estimators.^46^

Finally, outcome type imposes scale-specific constraints. For both binary and survival outcomes, `infer_confounding()` and data-driven texture calibration are unavailable, so sensitivity and prospective analyses fall back to default settings. The effect scale is also non-collapsible for both. The log-odds ratio (binary) and the Cox log-hazard ratio (survival) each make the product decomposition approximate. For survival, the approximation error grows with the magnitude of the conditional log-hazard ratios and does not vanish with sample size, so direct + indirect effects need not equal the total effect on the log-HR scale when effects are large. The RMST scale is exact but higher-variance and is the appropriate complement when strong effects are anticipated.^51^ For binary outcomes the log-odds-ratio decomposition is approximate in the same way, whereas the risk-difference scale is collapsible and yields an exact decomposition but can produce out-of-range fitted probabilities under the linear probability model.

Natural extensions include doubly-robust semiparametric estimators building on recent proximal path-specific inference work,^24,46^ bridge-function goodness-of-fit tests based on residual orthogonality, vine-copula extensions to the texture model, and an accelerated failure time outcome stage that would eliminate the non-collapsibility approximation inherent in the Cox model. The payoff of these practices remains to be demonstrated at scale: applying ICONIC’s diagnostic and recommendation workflow across a biobank with multiple cohorts would test whether data-calibrated estimator selection improves replication of causal mediation findings.

## Methods

### The causal model

All estimators are evaluated against a structural causal model (SCM, **Figure 2**). Let *X* denote the (scaled) exposure, *M* a mediator, *Y* an outcome feature, *G*_1_ a genetic instrument for the exposure, *G*_2_ a genetic instrument for the mediator, *W* a negative-control feature, *U* = (*U*_1_, …, *U_k_*) a *k*-dimensional vector of unmeasured confounders, *P* a latent population-structure variable, and *C* measured covariates. Each path is confounded by a distinct linear combination of the *k* confounders: the *X* → *M* path by the composite *U_XM_* = *U*^T^*λ_XM_* and the *M* → *Y* path by *U_MY_* = *U*^T^*λ_MY_*, where *λ_XM_* and *λ_MY_* are unit-norm loading vectors. When *λ_XM_* = *λ_MY_* a single confounder composite drives both paths; when they are distinct the two paths are confounded by overlapping or disjoint combinations of the *k* confounders:

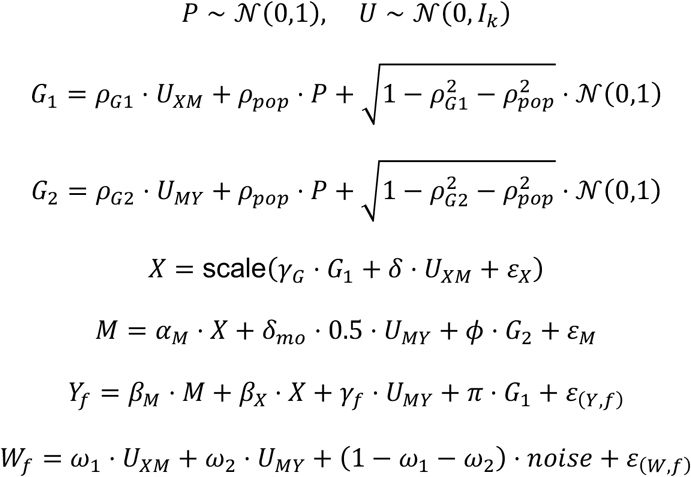

where *γ_G_* is a tunable instrument-strength parameter (default 0.6), *ε_X_*, *ε_M_*, *ε*_(*Y,f*)_, *ε*_(*W,f*)_ ∼ N(0, *σ*^2^) are independent errors, and scale(⋅) standardizes to unit variance. The noise terms *ε*_(F,*f*)_ and *ε*_(W,*f*)_ may be correlated across features, with the correlation structure learned from the user’s data (see Generative pipeline description, below). When no feature-level correlations are available, the noise is independent across features. The total effect of *X* on *Y* is *τ* = *β_X_* + *α_M_* ⋅ *β_M_*, decomposed into a natural direct effect NDE = *β_X_* and a natural indirect effect NIE = *α_M_* ⋅ *β_M_*. The structural equations are linear-additive with Gaussian errors, yielding a closed-form total effect that enables unambiguous bias assessment. The design mirrors the linear regression models routinely used in omics epidemiology; nonlinear dose-response and non-Gaussian errors are not evaluated here. The pluggable negative-control mechanism and generative texture model provide an extension path for nonlinear data-generating processes in future work.

### The three identifying resources and their assumptions

- *G*_1_, a genetic instrument (e.g., a polygenic score for GDM risk). It moves *X* but is independent of *U* and affects *Y* only through *X*. This is the Mendelian randomization premise of relevance, exclusion restriction, and exogeneity.^52^
- *G*_2_, a genetic instrument for the mediator (e.g., a cis-eQTL or elastic net of cis-eQTLs from germline genotypes). It moves *M* but is independent of *U* and has no direct path to *Y*. When available, it identifies the mediator-outcome relationship separately from the exposure, enabling point identification of natural effects under mediator-outcome confounding. More generally, let *G_m_* denote the genetic instrument for mediator *M_m_*, where *m* = 1, …, *K*, with one instrument used per mediator.
- *W*, negative-control outcomes. Features that share the confounder composites *U_XM_* and *U_MY_* with *Y* but are not on the causal path from *X*. ICONIC accepts path-specific panels: *W*_1_ proxies the exposure-mediator confounder composite *U_XM_* (coverage *ω*_1_) and *W*_2_ proxies the mediator-outcome confounder composite *U_MY_* (coverage *ω*_2_). A single shared panel is the special case *W*_1_ = *W*_2_. Each panel is adequate to the extent that it covers its path’s confounder composite. When only one path-specific panel is supplied (a lone *W*_1_ or *W*_2_), ICONIC derives the pooled panel *W* from it so that the single-panel estimators (DIRECT, COCA, PGC) and the negative-control validity/completeness screens operate on the available controls; the two-bridge estimators (PGC2, PGC2Gm) require both panels and remain gated unless the analyst sets `recycle_lone_panel = TRUE` to reuse the lone panel as both bridges (an opt-in that assumes one panel is complete for both composites). This is the negative-control and proximal-inference premise.^11,29^

### The estimators

ICONIC implements five estimators for the total effect, each leaning on a different identifying assumption, and three additional estimators for mediation when a mediator-specific instrument *G*_2_ or a negative-control panel *W* covering each path’s confounder composite is available (**Table 2**). All estimators except COCA extend to time-to-event (survival) and binary outcomes via two-stage predictor substitution, in which the first-stage regressions remain OLS and only the outcome stage switches: to a Cox model or RMST pseudo-observations for survival, and to a logistic regression (log-odds-ratio scale) or a linear probability model (risk-difference scale) for binary outcomes. The full survival and binary architectures are described in **Supplementary Methods S1** and **S13**.

### Total-effect estimators

The five total-effect estimators are: UNADJ (regresses *Y* on *X* alone, confounded reference floor); DIRECT (regresses *Y* on *X*, *G*, *W*, and covariates, using every observable but making no attempt to remove unmeasured confounding); COCA (the Control Outcome Calibration Approach,^12^ recovering τ̂ = −*β̂_X_/β̂_Y_* from the *W* ∼ *Y* + *X* regression, efficient when the negative control is a strong proxy but unstable when the denominator approaches zero); IV2SLS (uses *G* to instrument for *X* via two-stage least squares, purging the instrumented exposure of confounded variation because *G* ⊥ *U*); and PGC (proceeds in three stages: residualize *X* on *G*, regress the residual on *W* to construct a confounding proxy *Ŵ*, then fit *Y* ∼ *X* + *Ŵ*), in the spirit of the proximal-inference estimators of Miao et al.^29^ and Cui et al.^46^ Each estimator extends to mediation via the Baron-Kenny decomposition. The full regression equations for all five estimators (total-effect and mediation forms) are provided in **Supplementary Methods S2**.

### Mediation estimators

With a single genetic instrument, natural direct and indirect effects are not point-identified under mediator-outcome confounding.^22^ The instrument identifies the total effect *τ*, but decomposing it requires either no unmeasured mediator-outcome confounding (sequential ignorability) or additional instruments or negative controls that identify the mediator-outcome relationship separately.

When a mediator-specific instrument *G*_2_ is available, ICONIC implements a 2-stage MR mediation estimator IV2SLS2 that instruments both the exposure (with *G*_1_) and the mediator (with *G*_2_) via three sequential 2SLS stages, with weak-instrument checks (partial *F* ≥ 10) at each stage. IV2SLS2 optionally augments these stages with path-specific negative controls (*W*_1_ in the exposure first stage and *W*_2_ in the mediator and outcome stages) so that coverage of the confounder composites improves the estimate. Aa single pooled panel conditioned on in all three stages is a collider under multi-confounder designs and is therefore not used. When instrument independence is in doubt, ICONIC implements two-stage proximal mediation estimators that use the negative-control panel to purge confounding at each stage. The PGC2 estimator (no *G_m_* required) builds bridge proxies for the *X* → *M* and *M* → *Y* confounder composites from *W*_1_ and *W*_2_ respectively and requires completeness at both stages. A negative-control-augmented variant PGC2Gm uses *G_m_* in stage 2 to help isolate the effect of the *M* → *Y* confounder composite on *M*, with the bridge doing the confounding removal. Each bridge is adequate to the extent that its path-specific panel covers that path’s confounder composite (coverage *ω*_1_ and *ω*_2_), so a panel need only span its composite rather than consist of a single confounder. The full stage-by-stage regression equations for IV2SLS2, PGC2, and PGC2Gm are provided in **Supplementary Methods S2**.

### Parameters governing the identifying assumptions

- *Δ* (confounding strength) scales the effect of *U_XM_* on *X* and *U_MY_* on *Y*. When *δ* = 0 there is no unmeasured confounding.
- *Δ_mo_* (mediator-outcome confounding) controls whether *U_MY_* also affects *M*. When *δ_mo_* > 0, *U_MY_* affects both *M* and *Y*, creating mediator-outcome confounding.^21^
- *ϕ* (mediator-instrument strength) controls the effect of *G*_2_ on *M*; when *ϕ* = 0, no mediator instrument is generated. The motivating example is placental eQTL derived from fetal genotype, where cis-regulatory variants influence placental gene expression (*M*), providing exogenous variation in the mediator independent of maternal confounders.
- *π* (horizontal pleiotropy) controls a direct *G*_1_ → *Y* path that violates the exclusion restriction.^52^ When *π* = 0, *G*_1_ affects *Y* only through *X* and IV-based estimates are consistent.
- *ρ_G_*_1_ and *ρ_G_*_2_ (instrument exogeneity violations) control the correlation of *G*_1_ with *U_XM_* and *G*_2_ with *U_MY_*, respectively. Neither parameter can be estimated from data, as the confounders are unobserved, but their effects are assessed through sensitivity analyses.
- *ρ_pop_* (population structure) induces correlation between *G*_1_ and *G*_2_ through a shared latent factor *P*, modeling linkage or stratification.
- *ω*_1_ and *ω*_2_ (negative-control coverage) control how well the panel *W* captures the *X* → *M* and *M* → *Y* confounder composites, respectively. When coverage is high (*ω* → 1), the controls are strong proxies for the confounders; when coverage is low, the bridge-function estimators degrade.
- *k* (number of latent confounders) sets the dimension of the confounder vector *U*, and *λ_XM_* and *λ_MY_* (path-specific loading vectors) set how each path’s confounder composite combines the *k* confounders. The default is a shared composite (*λ_XM_* = *λ_MY_*, so a single confounder combination drives both paths and one panel *W* suffices). Setting *λ_XM_* and *λ_MY_* to distinct unit vectors (e.g., *e*_1_ and *e*_2_ at *k* = 2) makes the two paths draw on distinct confounders; intermediate overlapping loadings model the case in which the two paths share some but not all confounders.

The default parameter ranges span the empirically plausible regime for genetic-epidemiological instruments and negative controls: instrument strength *γ_G_* = 0.6 (default, first-stage partial *F* ≥ 10) swept down to *γ_G_* = 0.2 (weak, *F* < 5); mediator-instrument strength *ϕ* = 0.8 reflecting strong cis-eQTL first stages; and negative-control coverage *ω* = 0.7 (default) swept from 0.2 (weak) to 1.0 (perfect proxy). The `infer_confounding()` function automates calibration by estimating *δ* from the OLS-IV2SLS gap and *ω* from the NC-outcome *R*^2^, providing data-driven defaults that anchor the sensitivity grid to the user’s cohort. The full estimation procedure is provided in **Supplementary Methods S3**. To keep calibration computationally tractable on high-dimensional mediator panels, `infer_confounding()` estimates these parameters on a random subset of mediators (default 50, `max_infer_tasks`) rather than the full panel, and `iconic_sensitivity(confounding = “inferred”)` uses this subset directly. A precomputed `iconic_confounding` object can be passed to `iconic_sensitivity()` to avoid re-running the inference.

### Inference

Inference for the natural effects uses Wald tests throughout. The NDE p-value is the standard t-test for the exposure coefficient in the outcome regression, and the NIE p-value is a two-sided Wald test using a delta-method standard error (the Sobel test). ICONIC also supports a nonparametric bootstrap standard error (`se_method = “bootstrap”`, default `n_boot = 500` resamples) for estimators such as COCA whose delta-method variance is inflated by near-singular ratio terms. Confidence intervals are Wald intervals, *θ̂* ± 1.96 × SE(*θ̂*). The delta-method SEs do not propagate first-stage estimation uncertainty. The outcome-stage regression treats the fitted mediator *M̂* as if it were observed, so the reported SEs are conditional on the first-stage fit and may underestimate the true sampling variance when the first-stage instrument is weak. Full details of SE methods, CI construction, and empirical coverage assessment are provided in Supplementary Methods S4. For survival outcomes, inference uses the Cox partial-likelihood standard error on the log-HR scale and the OLS standard error on RMST pseudo-observations on the RMST scale (Supplementary Methods S1). For binary outcomes, inference uses the model-based standard error from the logistic outcome stage on the log-odds-ratio scale and the OLS standard error from the linear probability model on the risk-difference scale (Supplementary Methods S13).

### Composite null hypothesis test for the NIE

The NIE = *α_M_* ⋅ *β_M_* is a product of two coefficients, so the null *H*_0_: *α_M_* ⋅ *β_M_* = 0 is composite, holding under any of three cases (*α_M_* = 0 ∧ *β_M_* = 0, *α_M_* ≠ 0 ∧ *β_M_* = 0, or *α_M_* = 0 ∧ *β_M_* ≠ 0). The Sobel (delta-method) test is valid only under the partial-null cases; under the point null (sparse signals), the product of two independent normals follows a normal product distribution, making the Sobel test conservative.^53^ ICONIC provides a composite test (`se_method = “composite”`) implementing the closed-form JT-comp test of Huang (2019),^53^ which is most beneficial when signals are sparse. The complete derivation, variance estimation, clamping procedure, and limitation under the partial null are provided in **Supplementary Methods S5**.

### Estimator selection and recommendation

ICONIC ranks eligible estimators by per-estimand robustness, providing a transparent recommendation grounded in the data’s available identifying resources. Eligibility is determined empirically from the data, with instrument strength assessed via first-stage partial *F*-statistics^54^ (*F* ≥ 10), negative-control validity via the screens described below, and path completeness via the proximal completeness condition. The single-panel estimators (DIRECT, COCA, PGC) require a pooled panel *W*. When the analyst supplies only path-specific panels, ICONIC derives *W* automatically (as the average (*W*_1_ + *W*_2_)/2 when both are present, or as the lone panel itself when only one of *W*_1_/*W*_2_ is supplied) so these estimators remain eligible under a single outcome-side panel. The two-bridge estimators (PGC2, PGC2Gm) also require both *W*_1_ and *W*_2_; under a lone panel they are eligible only when `recycle_lone_panel = TRUE` reuses that panel as both bridges. No estimator is identified unless its assumptions are fulfilled, and the purpose of the diagnostic and sensitivity workflow is to test whether they are. The ranking reflects robustness to assumption violations rather than a guarantee of identification.

When a sensitivity analysis is available, eligible estimators are ranked separately for the NDE and the NIE by robustness across the assumption-violation surface, because the estimator that is most robust for the direct effect need not be most robust for the indirect effect. ICONIC supports three ranking criteria via the `criterion` argument to `iconic_recommend()`: `“combined”` (the default) balances worst-case bias and confidence-interval coverage across the assumption-violation surface; `“minimax_bias”` selects the estimator with the smallest worst-case absolute bias; and `“ci_coverage”` selects the estimator whose confidence intervals maintain nominal coverage furthest into the violation grid. Under the default combined criterion, each estimator receives a per-estimand robustness score derived from its worst-case bias and coverage degradation across the surface. Concretely, for estimator *e* and estimand *q* ∈ {NDE, NIE}, let

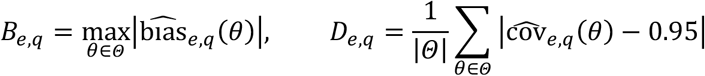

be the worst-case absolute bias and the mean absolute deviation of confidence-interval coverage from the nominal 0.95 across the violation grid *Θ*. Each quantity is min-max normalized across the eligible estimators, *B̃_e,q_* = (*B_e_*_,*q*_ − min*_e_B_e_*_,*q*_)/(max*_e_B_e_*_,*q*_ − min*_e_B_e_*_,*q*_) and likewise for *D̃_e,q_*, and the per-estimand robustness score is

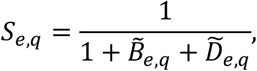

which lies in (0,1] and decreases as either component worsens. When coverage is unavailable on the surface, the score reduces to 1/(1 + *B̃_e,q_*). The alternative criteria use *S_e_*_,*q*_ = 1/(1 + *B_e_*_,*q*_) (`minimax_bias`) and *S_e_*_,*q*_ = 1/(1 + *D_e_*_,*q*_) (`ci_coverage`). The composite score is the minimum of the NDE and NIE scores, so an estimator that is robust for only one estimand is not rewarded. The composite is then multiplied by a confidence multiplier that discounts estimators whose identifying assumptions are only partially verifiable. The completeness penalty is 1.0 when the completeness condition is satisfied, 0.7 when borderline, 0.5 when capture is weak, and 0 when under-identified, and it applies only to the bridge-dependent estimators (DIRECT, COCA, PGC, PGC2, PGC2Gm), since the instrument-only estimators (IV2SLS, IV2SLS2) carry no completeness dependence and keep their full score. The final score is thus min(*S_e_*_,NDE_, *S_e_*_,NIE_) × *m_e_*, where *m_e_* is the completeness multiplier. The naive reference estimators (UNADJ, DIRECT) are demoted so that a confounded estimator cannot outrank an identifying one on the strength of low variance alone. These constants are fixed judgment parameters of the recommendation rule, disclosed here rather than learned from data. The recommendation is also per-scenario. `iconic_recommend()` returns both the overall preferred estimator and a `$per_scenario` field reporting the best estimator at the origin (no violation) and at the violated parameter configurations, because the estimator that is best when assumptions hold may differ from the one that degrades most gracefully when they are violated. The top-ranked eligible estimator is returned with a rationale documenting the assumptions required, the per-estimand robustness assessment, and the per-scenario breakdown.

### Generative pipeline

ICONIC employs two distinct simulation modes. The benchmark simulations use the structural generator (`generate_toy_data`) with parametric Gaussian noise to validate the estimators under controlled confounding scenarios with known ground truth **(Supplementary Figure S2)**. The data-calibrated simulations use the full generative pipeline, which trains a texture model on the user’s own data so that the sensitivity and prospective analyses are calibrated to realistic covariate, outcome, and mediator distributions **(Supplementary Figure S22)**. Estimator validation requires a transparent, reproducible data-generating process whose parameters are known exactly, while the user-facing workflow benefits from realistic data texture even at the cost of a learned noise model.

The design follows the plasmode approach^48^ of learning realistic data texture from observed data while imposing an investigator-specified causal effect, and the Credence framework^49^ of training a deep generative model to the empirical distribution with user-specified ground truth. A naive generative model trained directly on the full joint of (*X*, *M*, *Y*) cannot benchmark causal estimators, since it has no notion of the ground-truth effect *τ* and the unobserved confounder *U* is absent from observed data.^55^ The frugal parameterization principle^56^ addresses this by placing the causal effect of interest at the center of the model and learning only the surrounding nuisance texture from data.

The generative pipeline has three components: a causal skeleton implementing the SCM with *τ*, NDE, NIE, confounding strength *δ*, and negative-control coverage *ω* as known, tunable parameters; a pluggable negative-control mechanism (`nc_model`) that generates controls as coverage-weighted mixtures of the captured confounders plus noise; and a hybrid texture model combining a sample-level GAN for the low-dimensional covariate/exposure/outcome block with a feature-level Gaussian copula (following scDesign3^47^) for the mediator panel. The sample-level GAN follows the conditional tabular GAN (CTGAN) design,^57,58^ a generative adversarial network specialized for tabular data that accommodates mixed continuous and discrete columns through per-column normalization and one-hot encoding. It is trained on a low-dimensional per-sample frame of exposure, outcome, and mediator summary levels together with the encoded covariates, learning how these quantities co-vary across samples so that synthetic cohorts inherit realistic covariate structure and marginal distributions. Although the GAN’s training frame contains exposure, outcome, and mediator summaries, only the covariate columns and an exogenous outcome baseline are drawn from it at generation time. The synthetic exposure, mediator, and outcome are themselves regenerated by the causal skeleton, which imposes the investigator-specified *τ*, NDE, and NIE. No causal relationship among *X*, *M*, and *Y* is therefore learned from data, so the ground-truth effect is guaranteed by construction and requires no post-hoc validation.^50,59^ The full architecture (layer specifications, copula construction, residual-correlation handling, and the `residualize_on = “XCW”` double-counting mitigation) is provided in **Supplementary Methods S2**.

The model-selection workflow functions (`iconic_sensitivity()` and `iconic_prospect()`) train the texture layer directly from the user’s `iconic_data` object when no pre-trained model is supplied, so that synthetic covariate distributions, outcome marginals, and the mediator panel’s full feature-level structure match the user’s cohort. A pre-trained model may instead be attached at `iconic_data()` construction or passed via the `trained_gan` argument to avoid retraining across workflow steps.

### Sensitivity analyses

ICONIC provides complementary sensitivity analyses that profile estimator performance under violations of the identifying assumptions. Each analysis varies one or more untestable assumptions across a plausible range and reports how each estimator’s bias changes. We use “sweep” for single-parameter traversals and “grid” for multi-parameter factorial benchmarks.^60,61^ The full scenario manifest is exported by `scenario_manifest()` **(Supplementary Table S1)**.

#### Confounding and coverage grid

The `gan_sensitivity()` grid benchmarks every estimator across confounding strength *δ* (default {0.2,0.5,0.8}), negative-control coverage *ω* (default {0.3,0.7,1.0}), and the number of latent confounders *k* (default 1), with 50 replicates per scenario on texture-calibrated synthetic data. The model-selection workflow ranks estimators by the robustness criteria of `iconic_recommend()` described above. A companion negative-control validity check (`nc_validity_check()`) sweeps coverage *ω* ∈ {0.2,0.5,0.8,1.0} and *k* ∈ {1,2,3}, flagging scenarios as under-identified when *k* exceeds the number of valid controls.^13,29^ ICONIC estimates *k* from the data via parallel analysis on residualized outcomes in `infer_confounding()`, with a ±1 confidence interval, available when at least five outcome features are present.

#### Pleiotropy grid

A dedicated grid benchmarks every estimator across horizontal-pleiotropy strengths *π* (direct *G*_1_ → *Y* paths violating the exclusion restriction) and confounding strengths *δ*. Two arms are run per grid cell, an alternative arm (true effect > 0, yielding empirical power) and a null arm (true effect = 0, yielding empirical Type I error), so bias and false-positive rates are read from the same grid.

#### Instrument-exogeneity degradation surface

For mediation, `iconic_sensitivity()` generates a degradation surface over a grid of instrument- independence violations (*ρ_G_*_1_ × *ρ_G_*_2_), by default crossed with negative-control coverage (*ω*_1_ = *ω*_2_ swept over {0.3,0.7,1.0} on the diagonal), calibrated to the user’s data via the generative texture model. Confounding parameters may be left at fixed defaults, inferred via `infer_confounding()`, or specified manually. User-supplied sweep vectors for `omega_1`/`omega_2` (and a user-supplied `mo_confounding`) take precedence over the inferred scalars. When the analyst supplies a coverage sweep, the inferred value is not substituted, so the omega facet of the degradation surface is preserved under `confounding = “inferred”`. Tipping points are annotated at each grid cell where an estimator’s absolute bias crosses a user-defined threshold, defined marginally as the first grid value of *ρ_G_*_2_ (holding *ρ_G_*_1_ = 0) at which absolute bias crosses the threshold, and symmetrically for *ρ_G_*_1_ (holding *ρ_G_*_2_ = 0; computed for the NDE). This is distinct from the worst-case absolute bias, taken as the maximum over all parameter configurations. The surface’s use to delimit the trusted regime, quantify maximum error, and choose the robust estimator is described in the Results.

#### Mediation grid

A separate grid benchmarks NDE and NIE bias and Type I error across all parameters governing the identifying assumptions, including IV2SLS2 when a mediator instrument is available, PGC2 when a negative-control panel covering each path’s confounder composite is available, and PGC2Gm when both are available.

#### Prospective analysis

`iconic_prospect()` answers whether collecting an instrument or negative controls would change the estimate, and how strong the instrument would need to be. Phase 1 sweeps instrument strength (*γ_G_*) to show how estimates converge to the true effect as the instrument strengthens. Phase 2 runs a full prospective simulation at a target instrument strength, generating synthetic instruments and negative controls calibrated to the user’s sample size and confounding level, and reports the expected NDE and NIE estimates along with first-stage *F*-statistics and a recommended estimator (see Results).

### Data-calibrated confounding parameters

`infer_confounding()` estimates the held-fixed confounding parameters from the user’s data, supplying a `confounding = “inferred”` mode for `iconic_sensitivity()` and `iconic_prospect()`. Confounding strength is inferred from the gap between the unadjusted OLS and IV2SLS estimates; mediator-outcome confounding from the gap between the IV2SLS and IV2SLS2 NIEs; negative-control coverage from the NC-outcome *R*^2^; and the number of latent confounders via parallel analysis on residualized outcomes. The instrument-confounder correlations remain unestimable and are always swept. The inference uses estimator validity to calibrate a benchmark whose purpose is to test estimator validity, so inferred values should be read as best-case calibrations. The `confounding = “default”` mode avoids this circularity and remains the recommended starting point. The full estimation procedure is in **Supplementary Methods S3**.

### Negative-control validity checks

The validity of a negative-control analysis rests on three assumptions: that each control is independent of the exposure (A1) and of the genetic instrument (A2) given covariates and the unmeasured confounder, and that the valid controls are numerous enough to span the latent confounders (A3, completeness). When a mediator-specific instrument *G_m_* is used, a fourth assumption is required, namely that each control is independent of the mediator instrument (A2’). Violations bias the bridge-function estimators, so ICONIC implements an empirical check for each.^13^

ICONIC supports two screening criteria for A1 and A2: Benjamini-Hochberg FDR control at 0.10, and a magnitude-based criterion that flags controls whose association exceeds a user-specified effect-size threshold (default 0.10). The default `criterion = “both”` requires both. (A stricter threshold of 0.75 is used only in the COCA recount, which re-screens the controls that COCA’s ratio estimator is most sensitive to.) The COCA estimator is exempt from the A2 screen because it calibrates through the *W* ∼ *Y* + *X* ratio and does not require *W* ⊥ *G*_1_. Completeness (A3) is operationalized as a two-component condition combining a dimensional check (the number of valid controls must strictly exceed *k* for a “satisfied” verdict, with equality rated “borderline”) with a covariance-capture test (incremental *R*^2^ of *W* for *Y* above *C*, assessed against strong/weak thresholds via permutation; **Supplementary Figure S6D**). The number of latent confounders *k* is inferred from the data by Horn parallel analysis on the residualized-outcome correlation matrix (available when at least five outcome features are present, falling back to *k* = 1 otherwise), and a support/range check quantifies how much of the confounder proxy *Ũ* the panel explains and flags controls that add no unique coverage **(Supplementary Figure S6E)**. These completeness diagnostics are necessary-not-sufficient screens. They flag under-identified panels but cannot prove that the panel spans the confounder space; the full screening procedure is provided in **Supplementary Methods S6**.

### Case study rationale

#### Case study 1 - gestational diabetes, placental gene expression, and birth weight

The placenta is the interface between maternal metabolism and fetal growth, and placental gene expression is a plausible mediator of the effect of maternal metabolic dysfunction on birth weight. Placental gene modules mediate the effects of prenatal exposures on birth outcomes,^62,63^ and multi-omics colocalization places placental molecular features on the causal pathway from genetic variation to birth weight.^64,65^ Yet these mediation analyses have relied on standard methods that assume no unmeasured mediator-outcome confounding, an assumption unverifiable in observational birth cohort studies and likely violated by co-exposures, maternal diet, and genetic background. Maternal gestational diabetes mellitus (GDM) and birth weight offer a natural case study. GDM is among the most common metabolic complications of pregnancy, meta-analyses consistently report mean birth weight increases of approximately 100 grams and elevated odds of macrosomia among offspring of GDM mothers,^66,67^ and multiple placental mechanisms (altered glucose transporter expression, insulin-like growth factor signaling, inflammatory pathway activation, placental lactogen isoform diversity) provide biological plausibility for placental transcriptional mediation.^68,69^

#### Case study 2 - tobacco smoking, lung tumor expression, and lung cancer survival

Tobacco smoking and lung cancer provide a second, mechanistically distinct case study. Smoking is the leading preventable cause of cancer mortality, accounting for approximately 80% of lung cancer deaths in the United States and an estimated 1.3 million smoking-attributable lung cancer deaths globally each year.^70,71^ Smoking leaves a characteristic mutational signature (SBS4) in lung tumor genomes,^72^ and induces pervasive transcriptional alterations in both tumor and adjacent normal lung tissue, with smoking-associated gene-expression changes in the airway epithelium forming a detectable “field of injury” that persists in former smokers and predicts lung cancer risk.^73,74^ As in the placental setting, standard mediation analyses of smoking- expression-survival relationships are heavily confounded by co-exposures (occupational carcinogens, radon, air pollution), shared genetic background, and the field cancerization effect. The present framework addresses this by using genetic instruments and negative controls to identify and estimate mediation effects under weaker assumptions.

### Case study design

We apply ICONIC to two case studies: whether maternal GDM affects birth weight through placental gene expression, and whether tobacco smoking affects lung cancer survival through tumor gene expression. In both, the mediator set is restricted to features with a strong mediator instrument (panel rule, at least 50% of mediators with partial *F* ≥ 5), and the analysis estimates NDE and NIE using all eight estimators, with Benjamini-Hochberg FDR control at 20% and bootstrap confidence intervals (1,000 resamples). Both case studies apply the same sensitivity framework with instrument-strength diagnostics, negative-control independence tests, completeness checks, and the tipping-point sweep.

#### Case study 1 - GDM, placental gene expression, and birth weight

Data come from the GUSTO prebirth study^75^ (n = 164; maternal age at delivery 32.8 +/- 4.8 years; maternal ethnicity 70% Chinese, 15% Indian, 15% Malay; gestational age at delivery 38.7 +/- 1.2 weeks; birth weight 3.1 +/- 0.4 kg). GDM was measured as 2-hour plasma glucose concentration (mmol/L) during a 75 g oral glucose tolerance test (OGTT). Placental RNA-seq was quantified with Salmon against a tissue-specific long-read transcriptome reference,^76^ collapsed to gene-level counts via tximport, filtered to genes detected (TPM > 0) in at least 20% of samples, and inverse-normal transformed per gene to produce the expression matrix *M*. Maternal germline genotype was obtained from the Illumina OmniExpress + exome array^77^ and imputed to the South and East Asian reference Database (SEAD) panel,^78^ with QC and ancestry PCs computed via plink2. A polygenic score for GDM risk (2-hr OGTT)^79^ serves as *G*_1_, constructed via LDpred2-auto Bayesian shrinkage using an East Asian LD reference from the 1000 Genomes Project **(Supplementary Methods S7)**. An elastic net of fetal cis-eQTLs within ±1 Mb of each gene’s transcription start site serves as *G_m_*, generating cross-validated predicted expression as a composite instrument **(Supplementary Methods S8)**. Maternal DNA methylation was measured on the Illumina Infinium MethylationEPIC BeadChip (EPIC 850k),^77^ and used to construct *W* as the top 50 principal components of the methylation residual matrix after regressing out cell composition and technical covariates **(Supplementary Methods S9).** These negative-control CpG sites are selected for predictive value for gene expression rather than for linkage to maternal GDM-associated variants. Covariates comprise sex, maternal ethnicity, gestational age, fetal ancestry PCs, HCP latent factors (from PicardTools QC metrics on STAR BAMs), and RUVr unwanted-variation factors (from upper-quartile-normalized gene counts via edgeR + RUVSeq, *k* = 3).

#### Case study 2 - Smoking, lung tumor expression, and lung cancer survival

Data come from TCGA,^80,81^ combining lung adenocarcinoma (LUAD, n ∼ 522) and lung squamous cell carcinoma (LUSC, n ∼ 504). Smoking exposure is measured as self-reported pack-years (from cigarettes per day and years smoked); smoking status (current, former, never) is recorded as a covariate. Because TCGA is composed entirely of cancer cases, case study 2 estimates the mediation of smoking’s effect on survival among diagnosed lung cancer patients rather than on lung cancer incidence. Tumor RNA-seq was aligned with STAR and quantified at the gene level with Salmon, serving as *M*. Germline genotype was obtained from Affymetrix SNP 6.0 arrays and imputed to the TOPMed r2 reference panel (GRCh38),^82^ providing the basis for all three genetic instruments. A published polygenic score for cigarettes per day from GSCAN^83^ (PGS003368; LDpred2-weighted, 1,055,811 variants, European ancestry) serves as *G*_1_, applied to the imputed dosages by direct scoring in plink2 **(Supplementary Methods S10)**. An elastic net of lung cis-eQTLs, with the per-gene cis-SNP set defined by GTEx v11 lung eQTLs,^4^ serves as the composite mediator instrument *G_m_*, using the same cross-validated procedure as case study 1 **(Supplementary Methods S11)**. Genetically predicted expression from non-lung tissue serves as *W*: GTEx v8 tibial-nerve FUSION prediction weights are applied to the TCGA germline dosages and reduced to the top 20 principal components **(Supplementary Methods S12)**. The GSCAN PRS variants, lung cis-eQTL variants, and non-lung tissue eQTL weight variants are drawn from largely non-overlapping genomic regions. Covariates comprise age at diagnosis, sex, smoking status, ancestry PCs, HCP latent factors (from PicardTools sequencing-QC metrics), and RUVr unwanted-variation factors (from upper-quartile-normalized gene counts via edgeR + RUVSeq, *k* = 3). The ordinal stage and subtype indicators are excluded from the mediator screen as potential mediators. After genotype, expression, and covariate QC, the pooled LUAD+LUSC analysis retained *n* = 494 patients (193 deaths, 301 censored) and 3,674 mediator genes passing the panel instrument-strength rule.

#### Three-way comparison design

Each case study is analyzed in three arms. Arm 1 (Naive) runs `iconic_estimate` with covariates only, yielding the UNADJ estimate. Arm 2 (Prospective) runs `iconic_prospect` without instruments or negative controls, simulating best-case instruments. Arm 3 (Full) runs `iconic_estimate` with the real *G*_1_, *G_m_*, and *W*, preceded by `iconic_diagnose`, followed by `iconic_sensitivity` and `iconic_recommend`. Comparing Arm 2 to Arm 3 validates the prospective tool; where real instruments underperform the simulation, the diagnostics flag the violation. The case studies differ in exposure type (metabolic vs. behavioral), mediator tissue (placenta vs. lung tumor), and outcome scale (continuous vs. time-to-event).

## Ethics Approval

All study protocols follow the principles of the Declaration of Helsinki. The GUSTO prebirth study was approved by the National Healthcare Group Domain Specific Review Board and the SingHealth Centralized Institutional Review Board. Informed written consent was obtained from all mothers at recruitment. All data presented in this manuscript and associated materials are published in accordance with the terms of these approvals and the informed consent obtained from study participants. The secondary analysis of GUSTO cohort data reported here was approved by The University of Texas MD Anderson Cancer Center Institutional Review Board.

For the cancer genomics case study, controlled-access data from The Cancer Genome Atlas (TCGA; dbGaP accession phs000178) and the Genotype-Tissue Expression (GTEx) project (dbGaP accession phs000424) were obtained through the NIH database of Genotypes and Phenotypes (dbGaP) under approved data-access request. TCGA and GTEx data are de-identified; informed consent was obtained by the contributing studies as part of the original data collection, and secondary analysis was performed under the terms of the dbGaP Data Use Certification. The remaining data resources used in this study are openly available, aggregate-level summary statistics or prediction model weights (GWAS Catalog GCST90566423; PGS Catalog PGS003368; GTEx v8 FUSION TWAS prediction weights).

## Author Contributions

**Sean T. Bresnahan:** Conceptualization, Methodology, Software, Formal analysis, Data curation, Writing (original draft), Visualization. **Charis Xiong:** Conceptualization, Methodology, Software, Formal analysis, Writing (review and editing). **Taylor Head:** Writing (review and editing), Software quality control. **Yung-Han Chang:** Writing (review and editing), Software quality control. **Arjun Bhattacharya:** Conceptualization, Methodology, Supervision, Writing (review & editing). **Jonathan Y. Huang:** Conceptualization, Methodology, Supervision, Writing (review & editing).

## Data Availability Statement

The ICONIC R package is freely available at https://github.com/sbresnahan/iconic/ (v0.99.3). GUSTO cohort data are available to qualified researchers upon request, subject to approval by the GUSTO data access committee; see https://gustodatavault.sg/about/request-for-data for procedures. Data access is governed by applicable local laws and policies. TCGA data were accessed through the NCI Genomic Data Commons (https://gdc.cancer.gov/) and dbGaP (phs000178); GTEx data through dbGaP (phs000424). GDM (2-hour oral glucose tolerance test) summary statistics are publicly available from the GWAS catalog (GCST90566423). GSCAN cigarettes-per-day summary statistics are publicly available from the GWAS & Sequencing Consortium of Alcohol and Nicotine use (PGS003368). GTEx v8 FUSION TWAS weights for tibial nerve were obtained from the FUSION website (https://gusevlab.org/projects/fusion). All analysis code for the simulation benchmarks, case studies, and manuscript figures is publicly available at https://github.com/sbresnahan/iconic-manuscript.

## Acknowledgements

Some results shown here are in part based upon data from the GUSTO study, for which we thank the GUSTO participants and study team. Other results shown here are in part based upon data generated by the TCGA Research Network (https://www.cancer.gov/tcga). The Genotype-Tissue Expression (GTEx) Project was supported by the Common Fund of the Office of the Director of the National Institutes of Health and by NCI, NHGRI, NHLBI, NIDA, NIMH, and NINDS. Biomni (Phylo), an AI biomedical research platform,^84^ was used to assist with package documentation writing, the translation of statistical procedures implemented in the package code into mathematical notation and LaTeX, and the renumbering of figures and tables during manuscript revision; all AI-assisted outputs were reviewed and verified by the authors.

