## Supplementary Materials for "ICONIC: An R Package for Integrating Instrumental Variable- and Negative-Control-Informed Causal Discovery and Diagnostics in Multiomic Studies"

**Table of Contents**

**Supplementary Figures**
Figure S1. Simulation benchmark of eight ICONIC estimators under unmeasured confounding
Figure S2. Benchmark simulation mode: the structural synthetic-data generator (`generate_toy_data`)
Figure S3. Estimator bias and Type I error under horizontal pleiotropy
Figure S4. IV2SLS bias as a function of instrument strength
Figure S5. Negative-control coverage of each path's confounder composite for the PGC2-family mediation estimators
Figure S6. Simulation sweeps of empirical negative-control validity diagnostics
Figure S7. Robustness to feature-level correlations in the outcome and negative-control panels
Figure S8. Worst-case bias on the degradation surface across feature-level correlation in the outcome and negative-control panels
Figure S9. U/W heterogeneity: completeness capture and estimator robustness under heterogeneous proxy strength
Figure S10. Total-effect benchmark: ICONIC vs. TwoSampleMR (IVW)
Figure S11. Mediation benchmark Scenario A (equivalence): ICONIC vs. CMAverse
Figure S12. Mediation benchmark Scenario B (differentiation): ICONIC vs. CMAverse vs. two-step MR
Figure S13. Composite-null mediation test benchmark
Figure S14. Pleiotropy benchmark: ICONIC vs. MR-Egger vs. MR-PRESSO (total effect)
Figure S15. Pleiotropy benchmark: NDE/NIE decomposition (ICONIC two-instrument design)
Figure S16. Survival outcome benchmark: ICONIC estimators under confounding
Figure S17. Binary outcome benchmark: ICONIC estimators under confounding
Figure S18. GUSTO case study: degradation surface for the recommended estimator
Figure S19. GUSTO case study: forest plot of NDE and NIE for the top significant mediators per method by NIE
Figure S20. TCGA lung cancer case study: degradation surface for the recommended estimator
Figure S21. TCGA lung cancer case study: bootstrap forest of NDE and NIE for the top 10 significant mediators per method by NIE
Figure S22. Data-calibrated simulation mode: the hybrid GAN-plus-copula generative texture pipeline

**Supplementary Tables**
Table S1. Scenario manifest for the simulation benchmarks

**Supplementary Methods**
S1. Time-to-event (survival) outcome support
S2. Generative pipeline architecture
S3. Data-calibrated confounding parameters
S4. Inference and CI/SE calibration
S5. Composite null hypothesis test for the NIE
S6. Negative-control validity checks
S7. Exposure instrument construction via LDpred2-auto
S8. Elastic net composite instrument construction
S9. Negative-control panel construction from methylation PCs
S10. Exposure instrument construction from a GSCAN cigarettes-per-day polygenic score
S11. Mediator instrument construction from GTEx v11 lung cis-eQTLs
S12. Negative-control panel construction from tibial-nerve predicted expression
S13. Binary outcome support

### Supplementary Figures

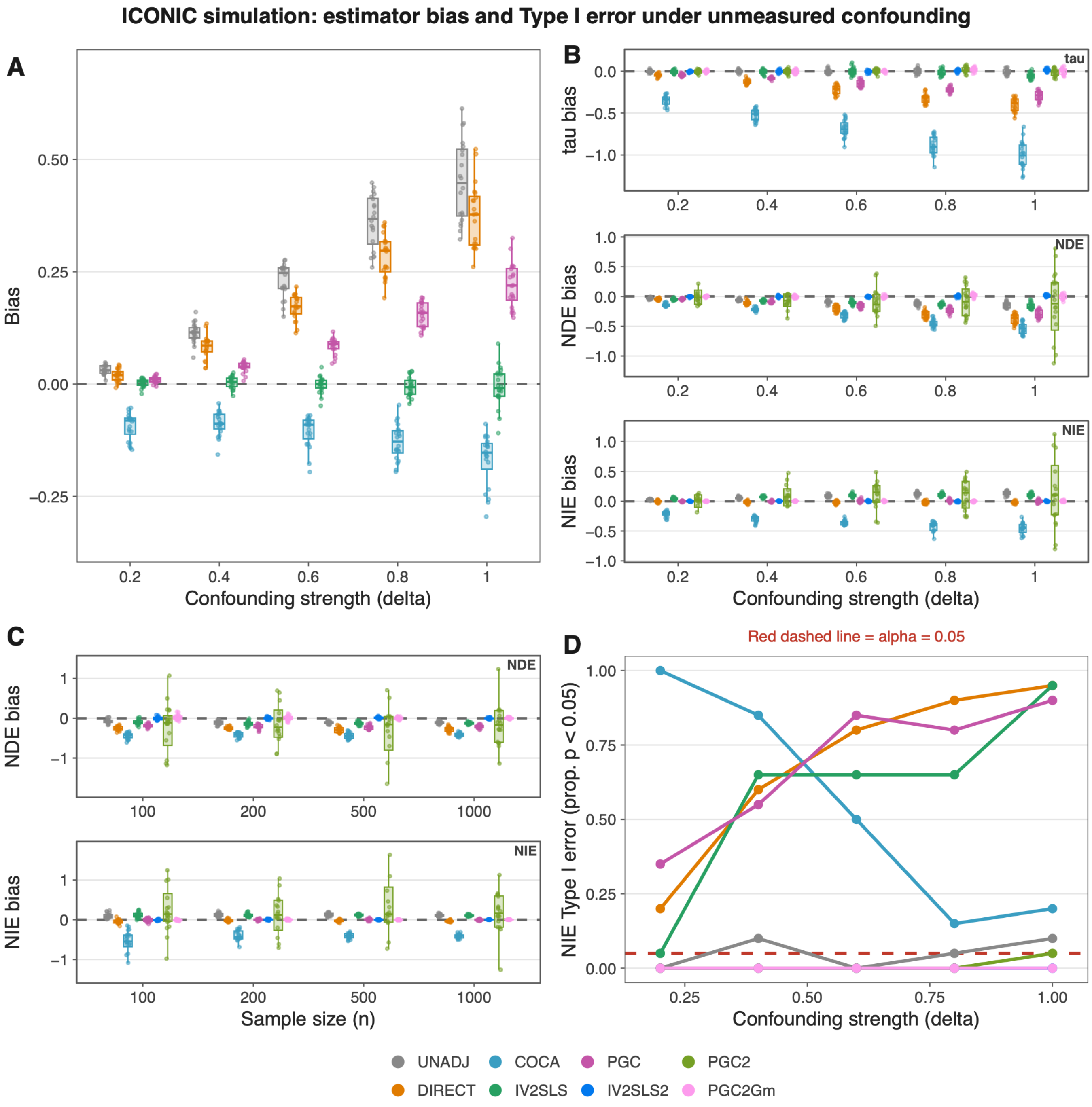


**Supplementary Figure S1. Simulation benchmark of eight ICONIC estimators under unmeasured confounding.** All panels use 50 simulation iterations and 10 outcome features. **Panel A** uses the total-effect DGP (single confounder $U$, no mediator, $\tau = 0.25$), where $\delta$ scales $U$'s effect on both $X$ and $Y$. **Panels B–D** use the mediation DGP ($k = 2$ confounders, $\varphi = 0.8$, $\rho_{G1} = 0.3$, $\omega_1 = \omega_2 = 0.7$, $\delta_{mo} = 0.8$). **(A)** Total-effect bias versus $\delta$. **(B)** Total-effect ($\tau$), NDE, and NIE bias versus $\delta$ for all eight mediation estimators. **(C)** NDE and NIE bias versus sample size $n$ at $\delta = 0.8$. **(D)** NIE Type I error rate under the null versus $\delta$. Red dashed line: nominal $\alpha = 0.05$. In all panels, $\omega = 0.7$ and $k = 1$. Abbreviations: NDE (natural direct effect); NIE (natural indirect effect); NC (negative control); MR (Mendelian randomization); GAN (generative adversarial network); DGP (data-generating process); RMST (restricted mean survival time); FDR (false discovery rate); 2SLS (two-stage least squares); IVW (inverse-variance weighted); OLS (ordinary least squares).

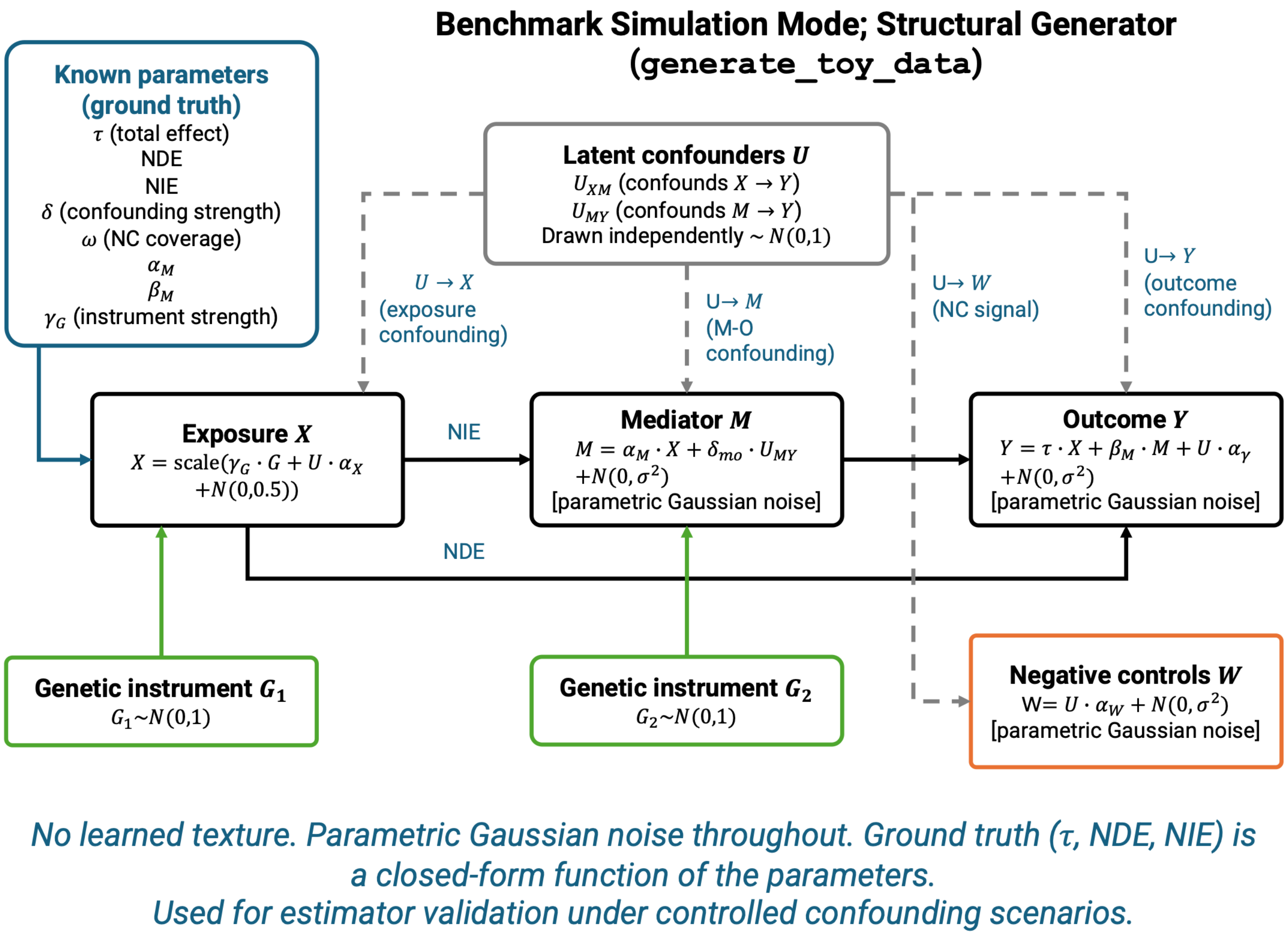


**Supplementary Figure S2. Benchmark simulation mode: the structural synthetic-data generator (`generate_toy_data`).** Schematic of the structural data-generating process used for estimator validation. A $k$-dimensional vector of latent confounders $U$ is drawn as $\mathcal{N}(0, I_k)$, and each path is confounded by a distinct linear combination of its components: the $X \to M$ backdoor by the composite $U_{XM} = U^\top \lambda_{XM}$ and the $M \to Y$ backdoor by $U_{MY} = U^\top \lambda_{MY}$. The exposure $X$, mediator $M$, and outcome $Y$ are generated from the structural causal model with parametric Gaussian noise at every stage: $X$ is a scaled function of its genetic instrument $G_1$ and $U_{XM}$; $M$ loads on $X$ and $U_{MY}$; and $Y$ loads on $X$ (direct path), $M$ (indirect path), and $U_{MY}$. Genetic instruments $G_1$ (for $X$) and $G_2$ (for $M$) are drawn as $\mathcal{N}(0,1)$, and the negative-control panel $W$ is generated as a coverage-weighted function of the confounder composites plus Gaussian noise. All ground-truth quantities (the total effect $\tau$ and its decomposition into NDE and NIE, confounding strength $\delta$, negative-control coverage $\omega$, instrument strengths, and the instrument-confounder correlations) are fixed, tunable parameters, so the ground truth is a closed-form function of the parameters rather than a learned quantity. This mode uses no learned texture and is used to validate the estimators under controlled confounding scenarios with exactly known ground truth. Abbreviations: NDE (natural direct effect); NIE (natural indirect effect); NC (negative control); MR (Mendelian randomization); GAN (generative adversarial network); DGP (data-generating process); RMST (restricted mean survival time); FDR (false discovery rate); 2SLS (two-stage least squares); IVW (inverse-variance weighted); OLS (ordinary least squares).

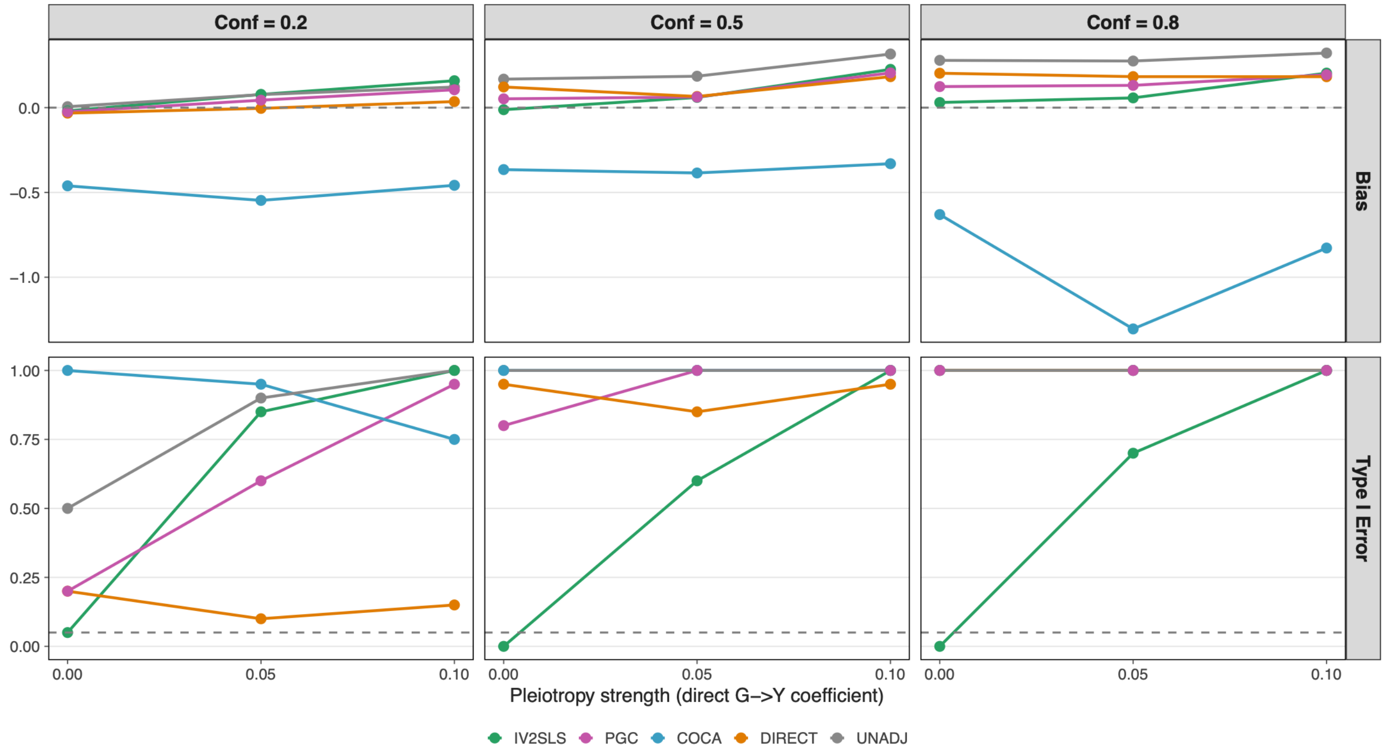


**Supplementary Figure S3. Estimator bias and Type I error under horizontal pleiotropy.** Bias (top row) and Type I error (bottom row) for the total-effect estimators (IV2SLS, PGC, COCA, DIRECT, UNADJ) as a function of pleiotropy strength shown across three confounding strengths. Abbreviations: NDE (natural direct effect); NIE (natural indirect effect); NC (negative control); MR (Mendelian randomization); GAN (generative adversarial network); DGP (data-generating process); RMST (restricted mean survival time); FDR (false discovery rate); 2SLS (two-stage least squares); IVW (inverse-variance weighted); OLS (ordinary least squares).

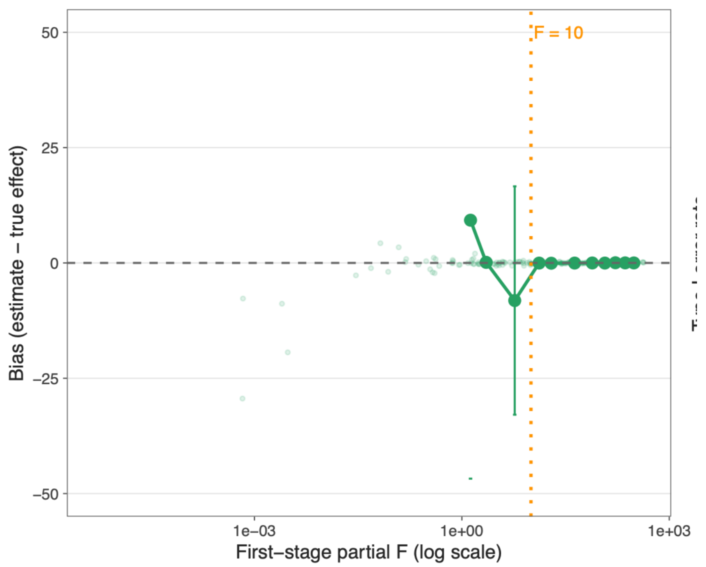


**Supplementary Figure S4. IV2SLS bias as a function of instrument strength.** Bias (estimate - true effect) for the IV2SLS total-effect estimator across a sweep of first-stage partial $F$-statistics for the exposure instrument, with the conventional weak-instrument threshold ($F = 10$) marked by the dashed line. Abbreviations: NDE (natural direct effect); NIE (natural indirect effect); NC (negative control); MR (Mendelian randomization); GAN (generative adversarial network); DGP (data-generating process); RMST (restricted mean survival time); FDR (false discovery rate); 2SLS (two-stage least squares); IVW (inverse-variance weighted); OLS (ordinary least squares).

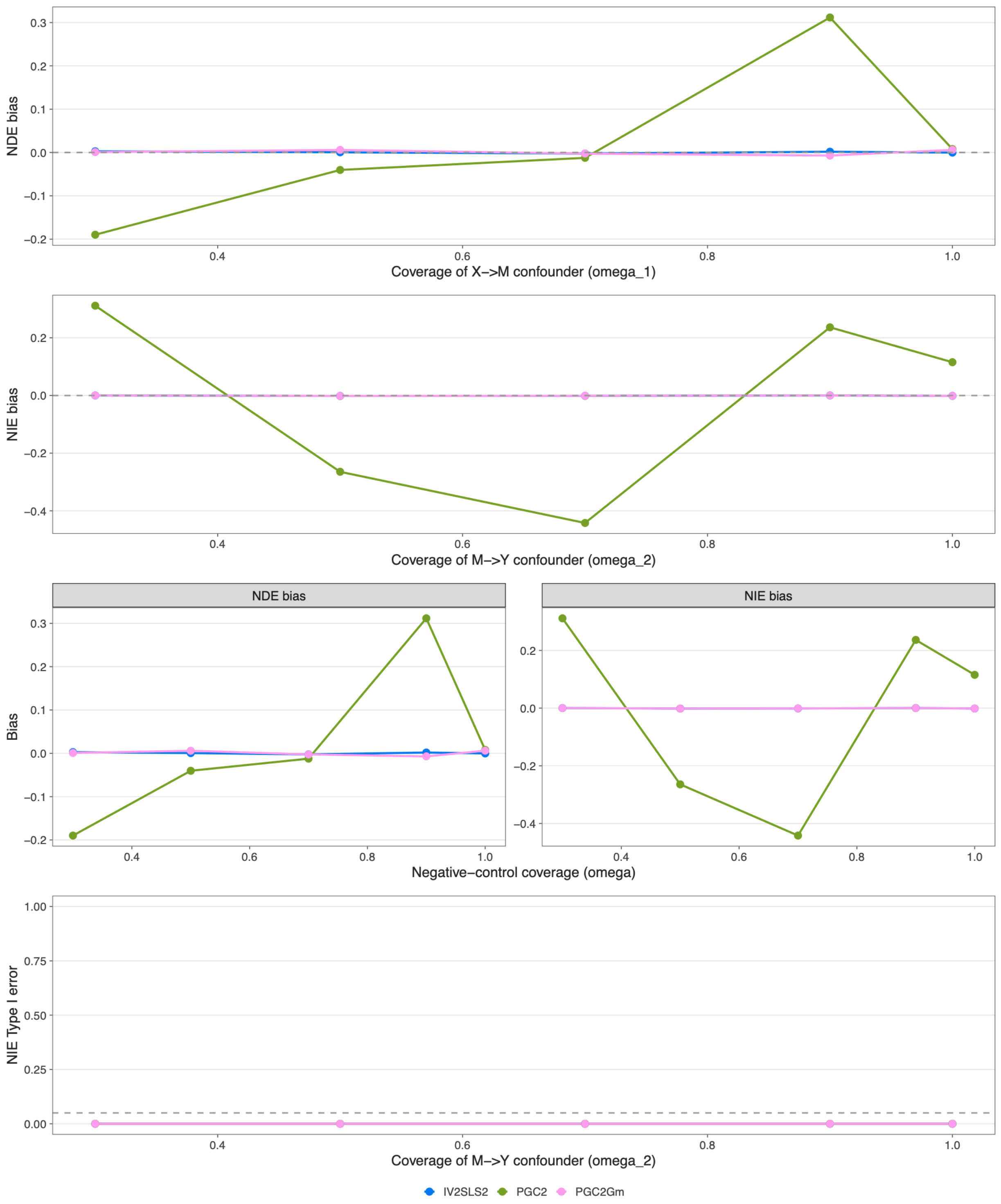


**Supplementary Figure S5. Negative-control coverage of each path's confounder composite for the PGC2-family mediation estimators.** Path-specific panels $W_1$ and $W_2$ proxy the two paths' confounder composites; adequacy is governed by how well each panel covers its path's composite ($\omega_1$ for the $X \to M$ composite, $\omega_2$ for the $M \to Y$ composite). **(A)** NDE bias as $\omega_1$ (coverage of the $X \to M$ composite) is swept from 0.3 to 1.0. **(B)** NIE bias as $\omega_2$ (coverage of the $M \to Y$ composite) is swept from 0.3 to 1.0. **(C)** NIE Type I error under the null as $\omega_2$ is swept. Abbreviations: NDE (natural direct effect); NIE (natural indirect effect); NC (negative control); MR (Mendelian randomization); GAN (generative adversarial network); DGP (data-generating process); RMST (restricted mean survival time); FDR (false discovery rate); 2SLS (two-stage least squares); IVW (inverse-variance weighted); OLS (ordinary least squares).

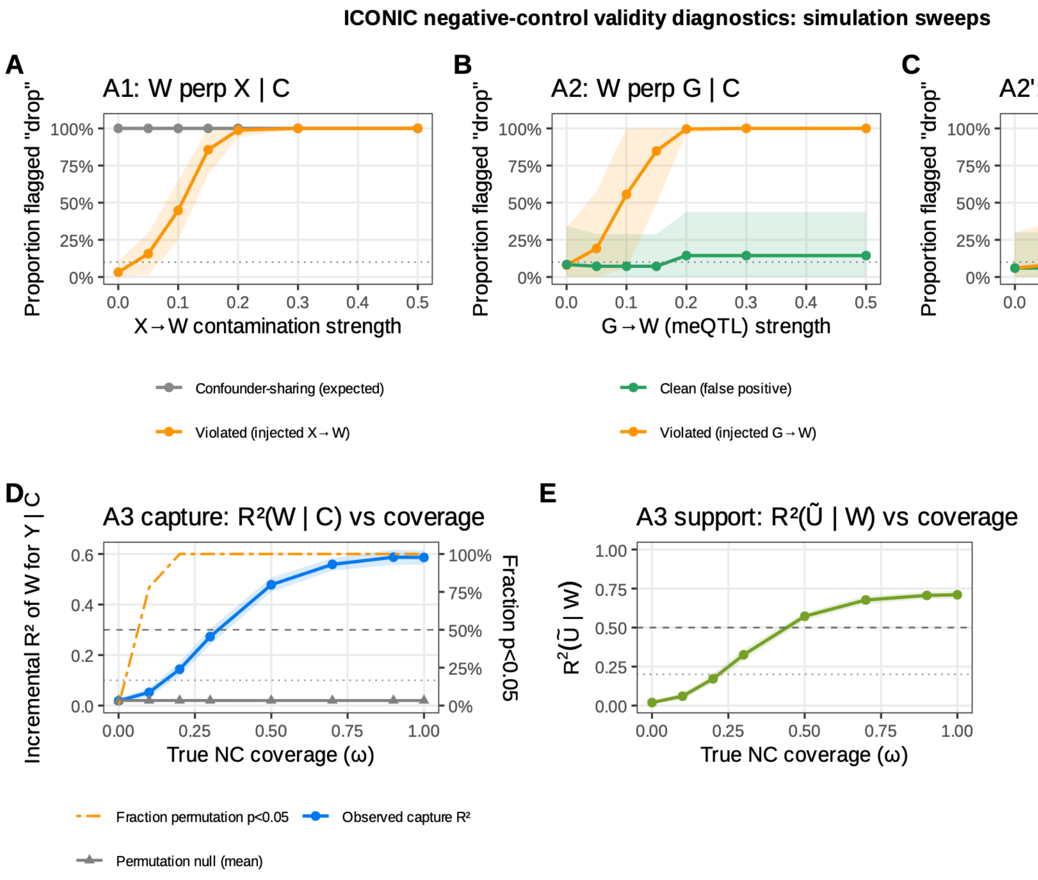


**Supplementary Figure S6. Simulation sweeps of empirical negative-control validity diagnostics. (A)** A1 ($W \perp X \mid C, U$): proportion of controls flagged versus injected $X \to W$ contamination strength. Five confounder-sharing controls (grey) share $U$ by construction; five noise controls (orange) carry the injected path. **(B)** A2 ($W \perp G \mid C, U$): proportion flagged versus injected $G \to W$ strength. COCA is exempt from the A2 screen. **(C)** A2' ($W \perp G_m \mid C, U$): proportion flagged versus injected $G_m \to W$ strength. **(D)** A3 covariance-capture component: incremental $R^2$ of $W$ for $Y$ above $C$ (mean $\pm$ SD) versus true negative-control coverage $\omega$. Dashed line: strong-capture threshold ($R^2 = 0.3$); dotted: weak ($R^2 = 0.1$). The grey series is the permutation-null mean (permuting the $W$–$Y$ association); the orange dash-dot series is the fraction of replicates with permutation $p < 0.05$ (right axis). **(E)** A3 support component: $R^2\left(\tilde{U} \mid W\right)$ (mean $\pm$ SD) versus $\omega$, where $\tilde{U} = \mathrm{resid}(X \sim G + C)$ is the instrument-purged exposure residual. Dashed line: broad-support threshold (0.5); dotted: partial (0.2). Panels A–C use $k = 1$ confounder; panels D–E use $k = 2$ confounders with distinct loadings. Data generated with $\varphi = 0.8$. All panels use $n = 500$ and 50 replications; each panel comprises 10 features; the capture test uses 200 permutations. The dotted line in panels A–C marks the nominal BH-FDR level (0.10). Abbreviations: NC (negative control); BH-FDR (Benjamini-Hochberg false discovery rate).

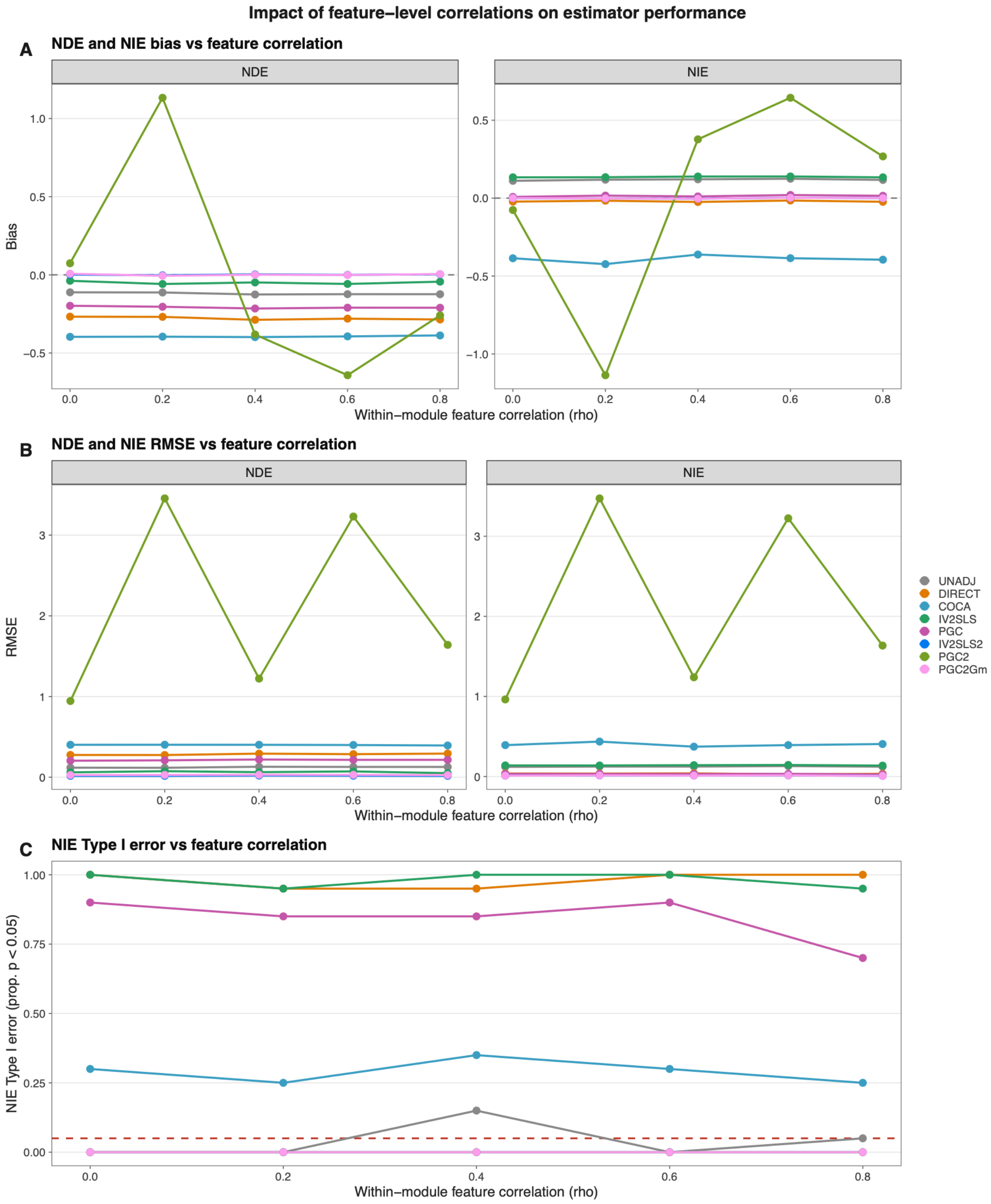


**Supplementary Figure S7. Robustness to feature-level correlations in the outcome and negative-control panels.** Estimator performance across a sweep of within-module feature correlation (`feat_cor`) from 0 (independent) to 0.8 (strong correlation), using a block-diagonal correlation matrix with $\lceil\sqrt{p}\rceil$ modules injected into the outcome and negative-control noise panels; the mediator panel's cross-feature dependence is modelled by the Gaussian copula and held fixed across the sweep. 50 replicates per grid point, $n = 500$, $p = 10$, $k = 2$ confounders with distinct path loadings, $\varphi = 0.8$, $\rho_{G1} = 0$, $\mathtt{mo\_confounding} = 0.8$. **(A)** NDE and NIE bias versus `feat_cor`, faceted by estimand, all 8 estimators. **(B)** NDE and NIE RMSE versus `feat_cor`, faceted by estimand. **(C)** NIE Type I error versus `feat_cor`, all 8 estimators, $\alpha = 0.05$ reference line. Abbreviations: NDE (natural direct effect); NIE (natural indirect effect); NC (negative control); MR (Mendelian randomization); GAN (generative adversarial network); DGP (data-generating process); RMST (restricted mean survival time); FDR (false discovery rate); 2SLS (two-stage least squares); IVW (inverse-variance weighted); OLS (ordinary least squares).

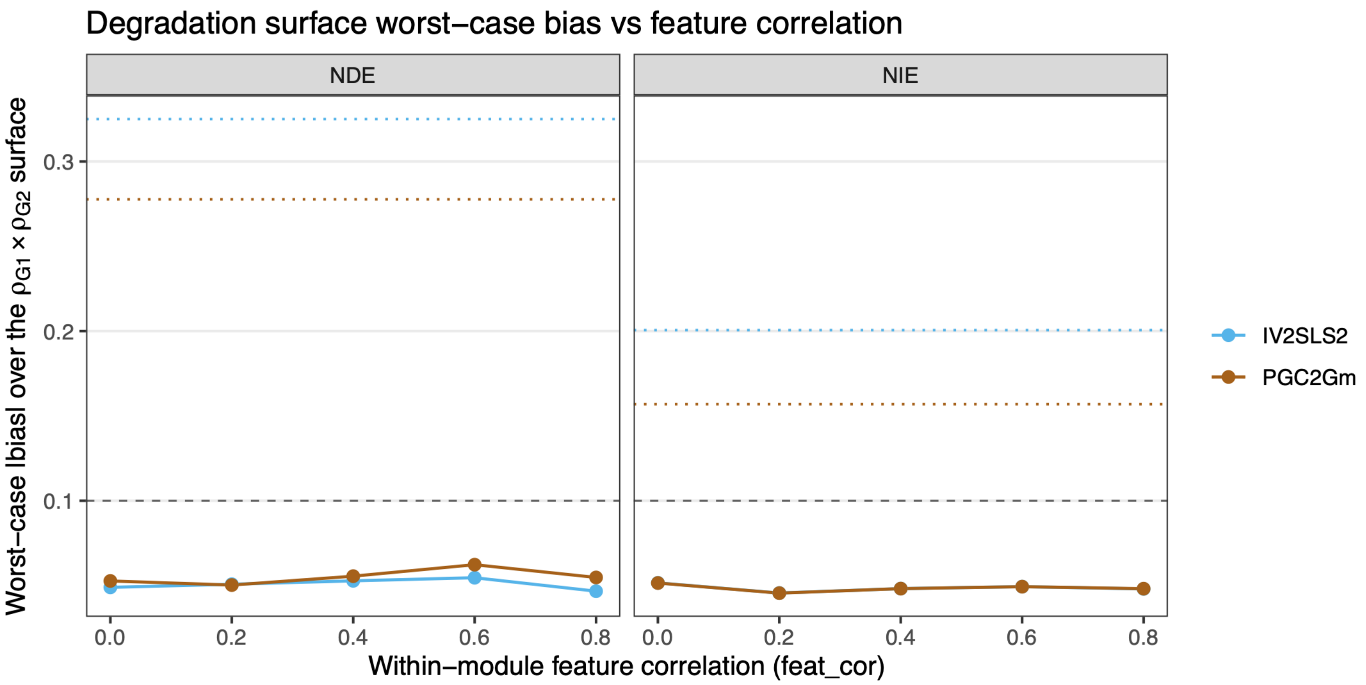


**Supplementary Figure S8. Worst-case bias on the degradation surface across feature-level correlation in the outcome and negative-control panels.** Across within-module feature correlation (`feat_cor` from 0 to 0.8 by 0.2, block-diagonal with $\lceil\sqrt{p}\rceil$ modules, injected into the outcome and negative-control noise panels), a $\rho_{G1} \times \rho_{G2}$ degradation surface was computed from 0 to 0.5 by 0.1 and collapsed to the worst-case absolute bias attained anywhere on that surface, shown for the two leading estimators (IV2SLS2, PGC2Gm) and faceted by estimand (NDE, NIE). Settings: $n = 500$, 50 replicates per cell, $p = 10$, $k = 2$ confounders with distinct path loadings, $\varphi = 0.8$, `mo_confounding` $= 0.8$, $\omega_1 = \omega_2 = 0.7$. Both instrument-exogeneity violations are swept at every `feat_cor` level, so worst-case bias reflects the joint effect of instrument violation and feature correlation. Dashed grey line marks the $|\text{bias}| = 0.10$ threshold. Dotted horizontal lines mark the worst-case bias attained by each estimator when the residual correlation structure is learned from the data by the generative texture model rather than imposed as a block-diagonal matrix. Abbreviations: NDE (natural direct effect); NIE (natural indirect effect); NC (negative control); MR (Mendelian randomization); GAN (generative adversarial network); DGP (data-generating process); RMST (restricted mean survival time); FDR (false discovery rate); 2SLS (two-stage least squares); IVW (inverse-variance weighted); OLS (ordinary least squares).

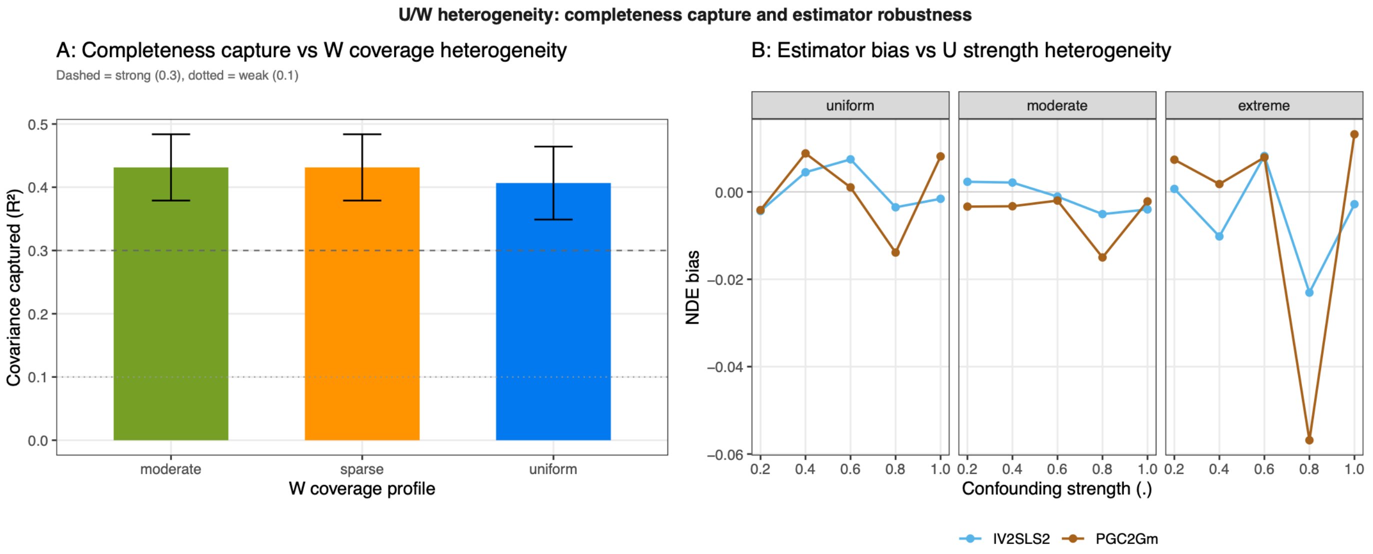


**Supplementary Figure S9. U/W heterogeneity: completeness capture and estimator robustness under heterogeneous proxy strength. (A)** Covariance-capture $R^2$ from `nc_completeness_capture()` across three $W$ coverage profiles: uniform (all at $\omega = 0.7$), moderate (0.9 to 0.2), and sparse (four strong, six near-zero). Dashed line: strong-capture threshold ($R^2 = 0.3$); dotted: weak ($R^2 = 0.1$). The coverage-level sweep of this diagnostic is shown in **Supplementary Figure S6D**. **(B)** NDE bias for IV2SLS2 and PGC2Gm across confounding strengths $\delta \in \{0.2, \ldots, 1.0\}$, faceted by $U$ strength profile: uniform, moderate, and extreme. Settings: $n = 500$, 50 replicates, $p = 10$, $k = 2$, $\varphi = 0.8$, $\rho_{G1} = 0.3$, $\texttt{mo\_confounding} = 0.8$, $\omega_1 = \omega_2 = 0.7$. Abbreviations: NDE (natural direct effect); NIE (natural indirect effect); NC (negative control); MR (Mendelian randomization); GAN (generative adversarial network); DGP (data-generating process); RMST (restricted mean survival time); FDR (false discovery rate); 2SLS (two-stage least squares); IVW (inverse-variance weighted); OLS (ordinary least squares).

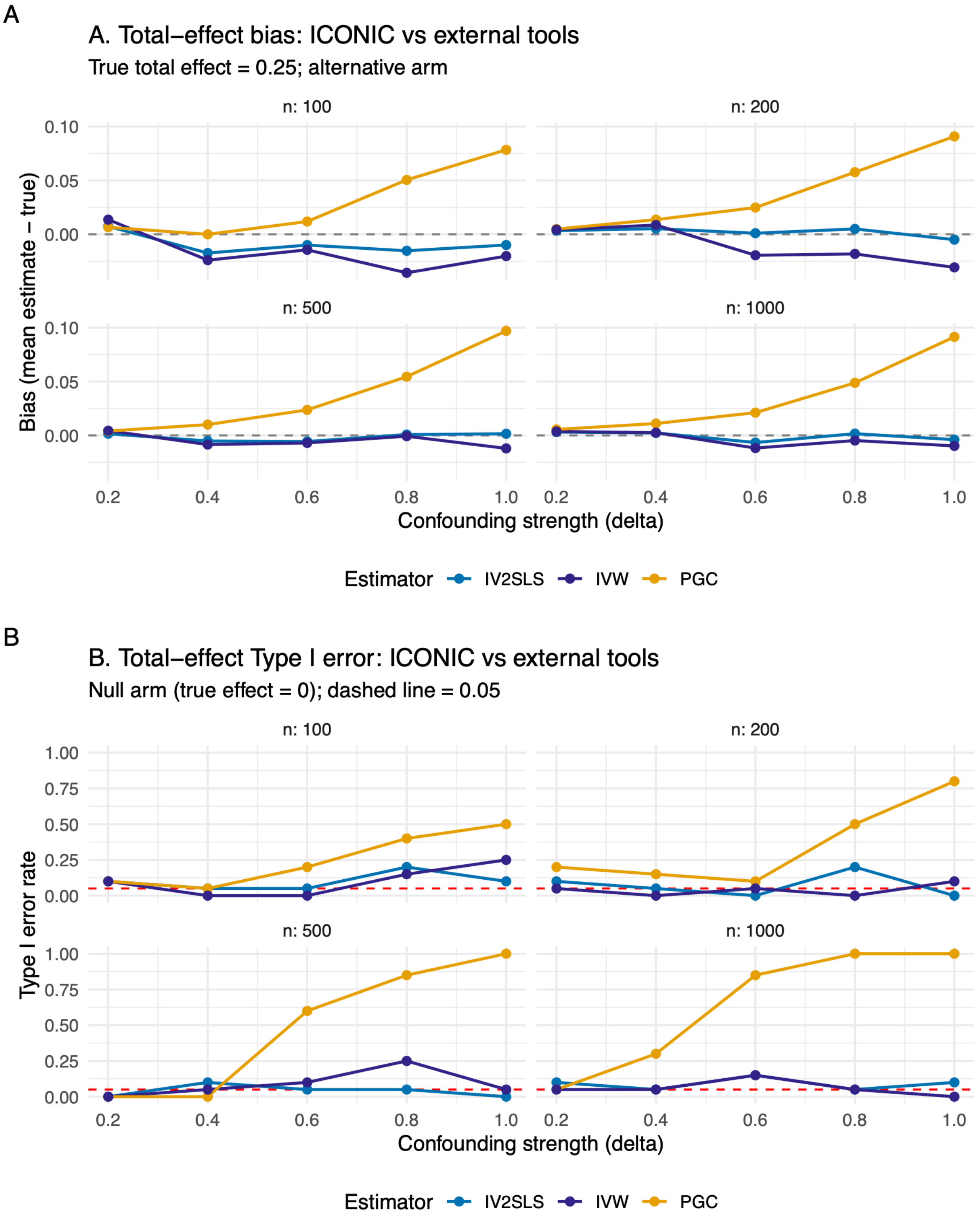


**Supplementary Figure S10. Total-effect benchmark: ICONIC vs. TwoSampleMR (IVW).** Bias (alternative arm, true $\tau = 0.25$) and Type I error (null arm, true $\tau = 0$) for five ICONIC estimators (UNADJ, DIRECT, COCA, IV2SLS, PGC) and the IVW analogue from TwoSampleMR, across confounding strength $\delta \in \{0.2, 0.4, 0.6, 0.8, 1.0\}$ and sample size $n \in \{100, 200, 500, 1000\}$ (faceted by $n$). **(A)** Bias versus $\delta$. **(B)** Type I error versus $\delta$; red dashed line marks the nominal $\alpha = 0.05$. IV2SLS and IVW are both 2SLS estimators. 50 replicates per cell, 10 features. Abbreviations: NDE (natural direct effect); NIE (natural indirect effect); NC (negative control); MR (Mendelian randomization); GAN (generative adversarial network); DGP (data-generating process); RMST (restricted mean survival time); FDR (false discovery rate); 2SLS (two-stage least squares); IVW (inverse-variance weighted); OLS (ordinary least squares).

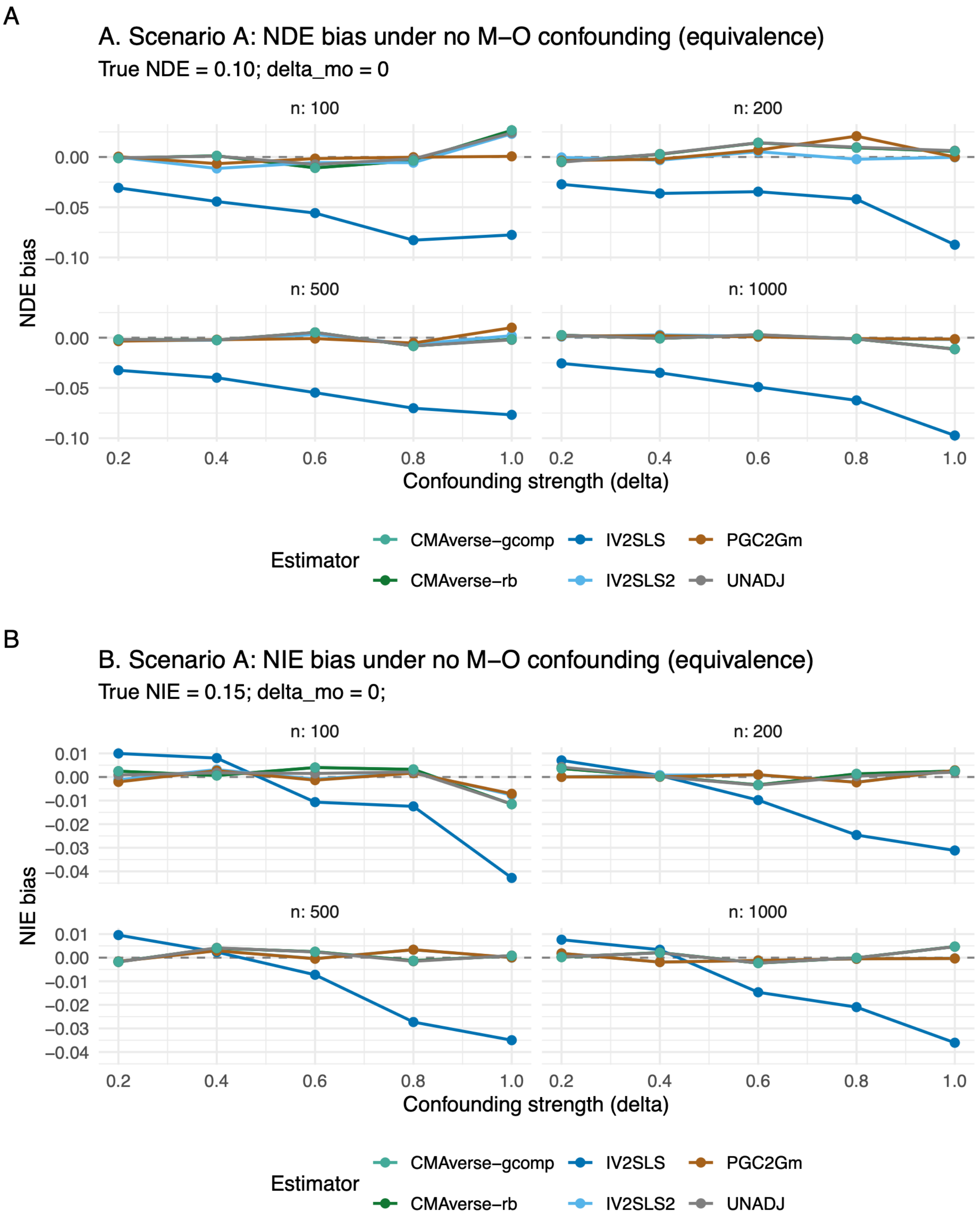


**Supplementary Figure S11. Mediation benchmark Scenario A (equivalence): ICONIC vs. CMAverse.** NDE and NIE bias (alternative arm) under no mediator-outcome confounding ($\delta_{mo} = 0$), across confounding strength $\delta$ and sample size $n$ (faceted by $n$). True NDE = 0.10, true NIE = 0.15. **(A)** NDE bias versus $\delta$. **(B)** NIE bias versus $\delta$. Methods shown: UNADJ, IV2SLS, IV2SLS2, PGC2Gm, CMAverse-rb, CMAverse-gcomp. 50 replicates per cell, 10 features. Abbreviations: NDE (natural direct effect); NIE (natural indirect effect); NC (negative control); MR (Mendelian randomization); GAN (generative adversarial network); DGP (data-generating process); RMST (restricted mean survival time); FDR (false discovery rate); 2SLS (two-stage least squares); IVW (inverse-variance weighted); OLS (ordinary least squares).

A

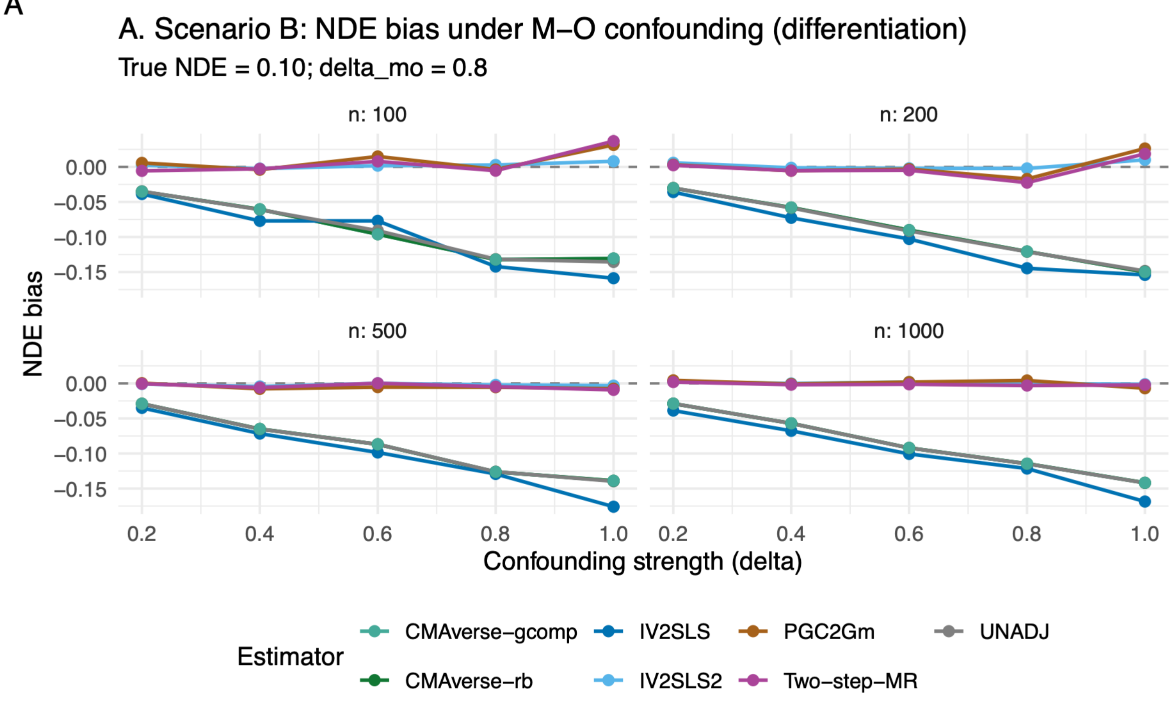


B

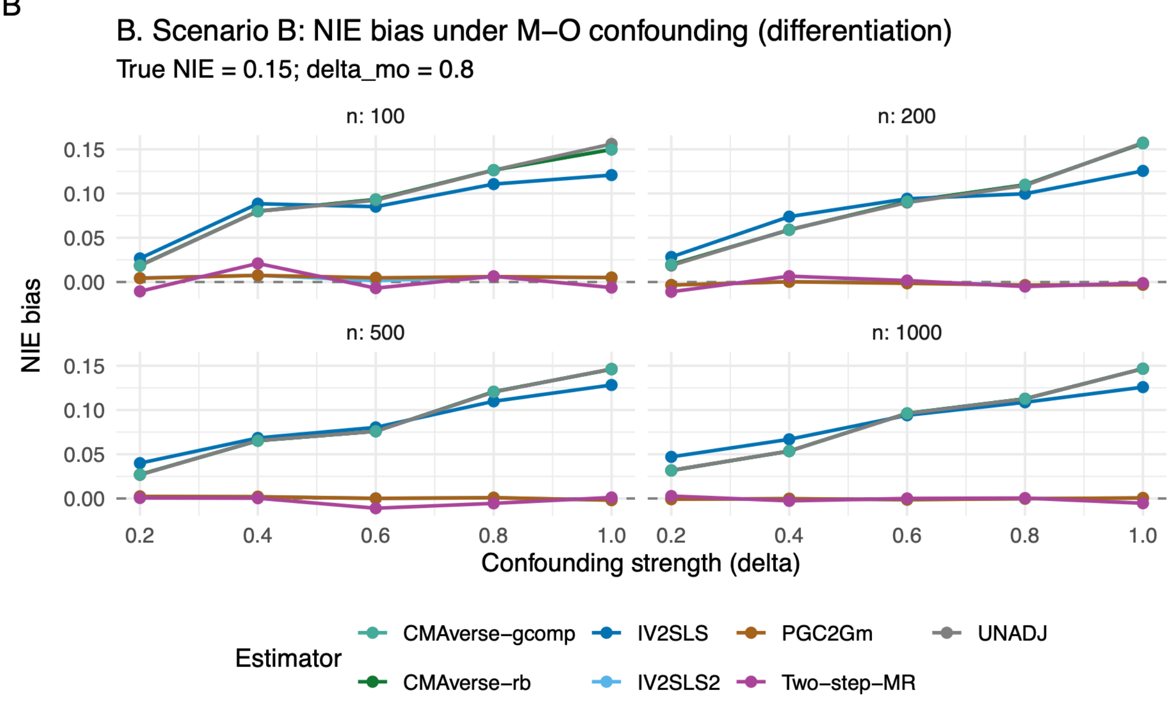


**Supplementary Figure S12. Mediation benchmark Scenario B (differentiation): ICONIC vs. CMAverse vs. two-step MR.** NDE and NIE bias (alternative arm) under mediator-outcome confounding ($\delta_{mo} = 0.8$), across confounding strength $\delta$ and sample size $n$ (faceted by $n$). True NDE = 0.10, true NIE = 0.15. **(A)** NDE bias versus $\delta$. **(B)** NIE bias versus $\delta$. Methods shown: UNADJ, IV2SLS, IV2SLS2, PGC2Gm, CMAverse-rb, CMAverse-gcomp, Two-step-MR. 50 replicates per cell, 10 features. Abbreviations: NDE (natural direct effect); NIE (natural indirect effect); NC (negative control); MR (Mendelian randomization); GAN (generative adversarial network); DGP (data-generating process); RMST (restricted mean survival time); FDR (false discovery rate); 2SLS (two-stage least squares); IVW (inverse-variance weighted); OLS (ordinary least squares).

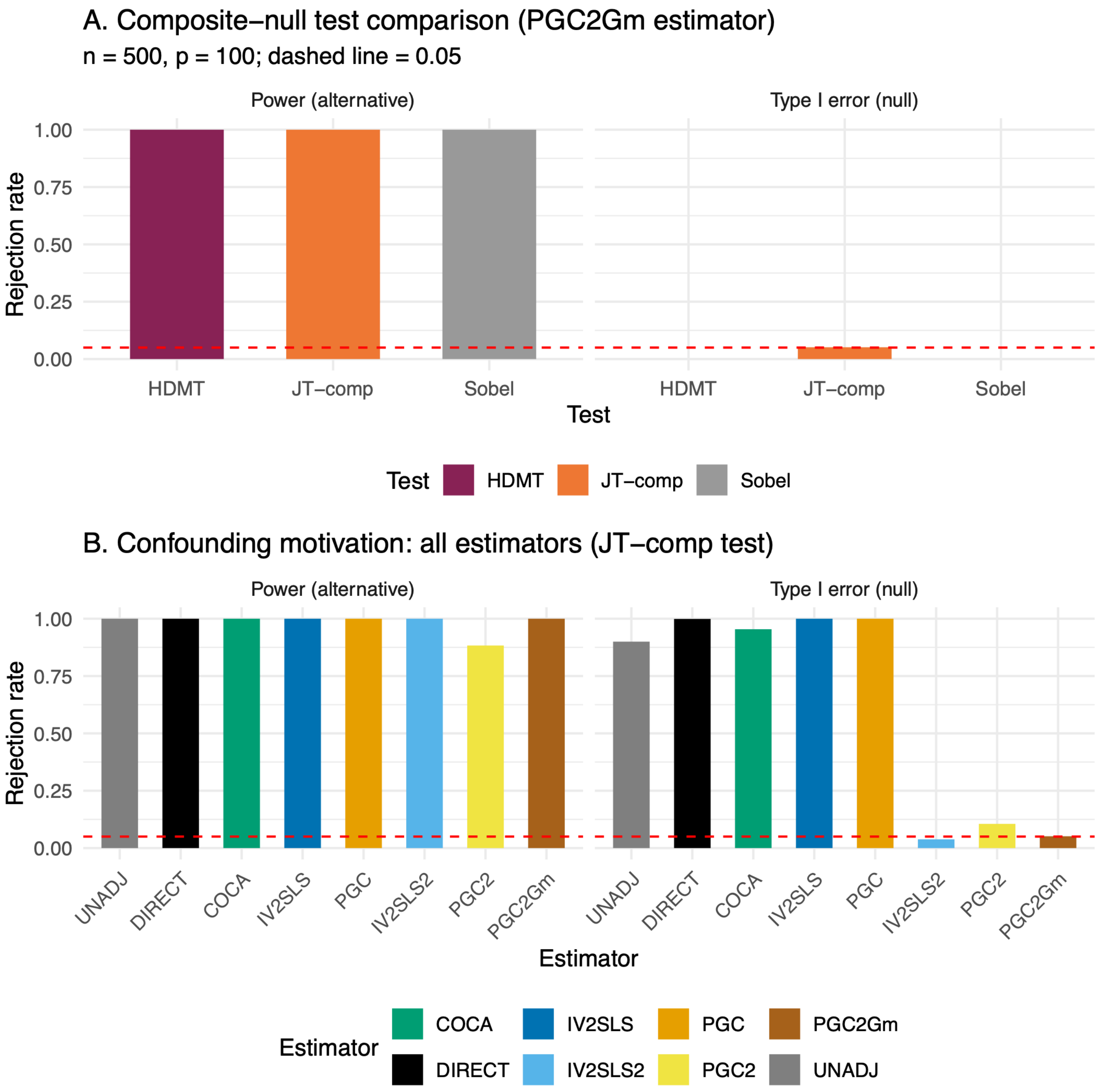


**Supplementary Figure S13. Composite-null mediation test benchmark.** Type I error (null arm, true NIE = 0) and power (alternative arm, true NIE = 0.15) for the joint test (JT-comp), Sobel test, and HDMT. **(A)** Test comparison on the deconfounded PGC2Gm estimator: JT-comp vs. Sobel vs. HDMT. **(B)** Confounding motivation: all eight estimators under the JT-comp test. Red dashed line marks $\alpha = 0.05$. $n = 500$, 100 features, 50 replicates. Abbreviations: NDE (natural direct effect); NIE (natural indirect effect); NC (negative control); MR (Mendelian randomization); GAN (generative adversarial network); DGP (data-generating process); RMST (restricted mean survival time); FDR (false discovery rate); 2SLS (two-stage least squares); IVW (inverse-variance weighted); OLS (ordinary least squares).

A

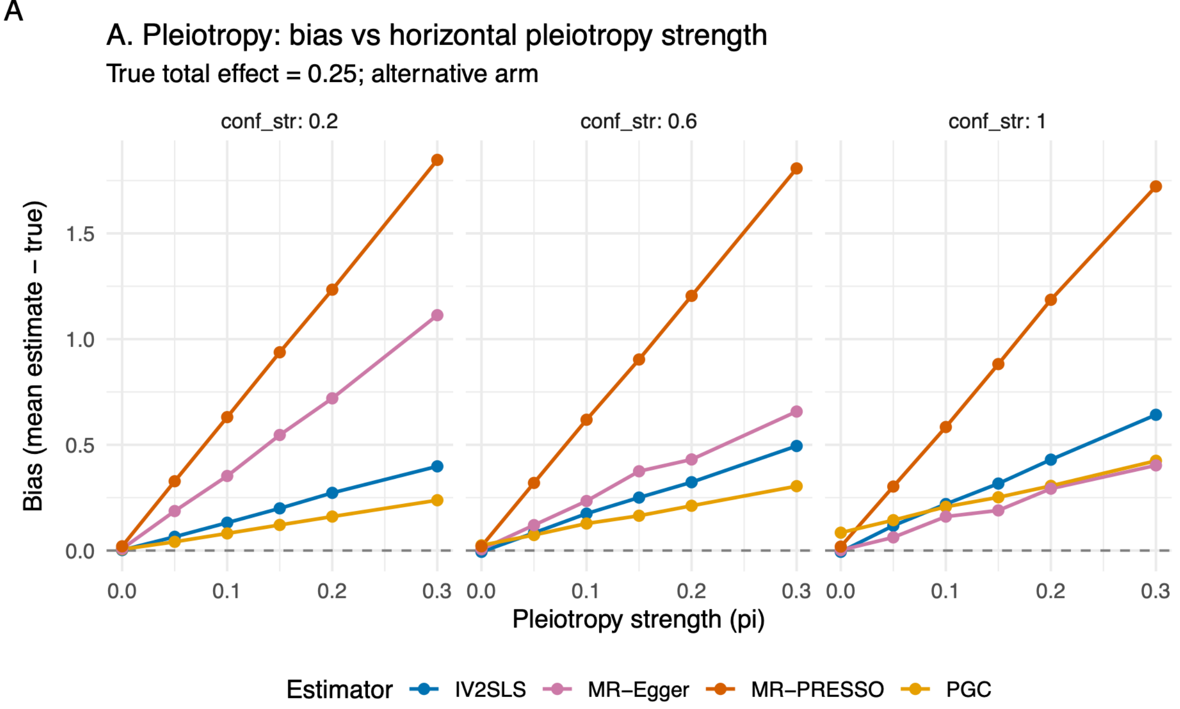


B

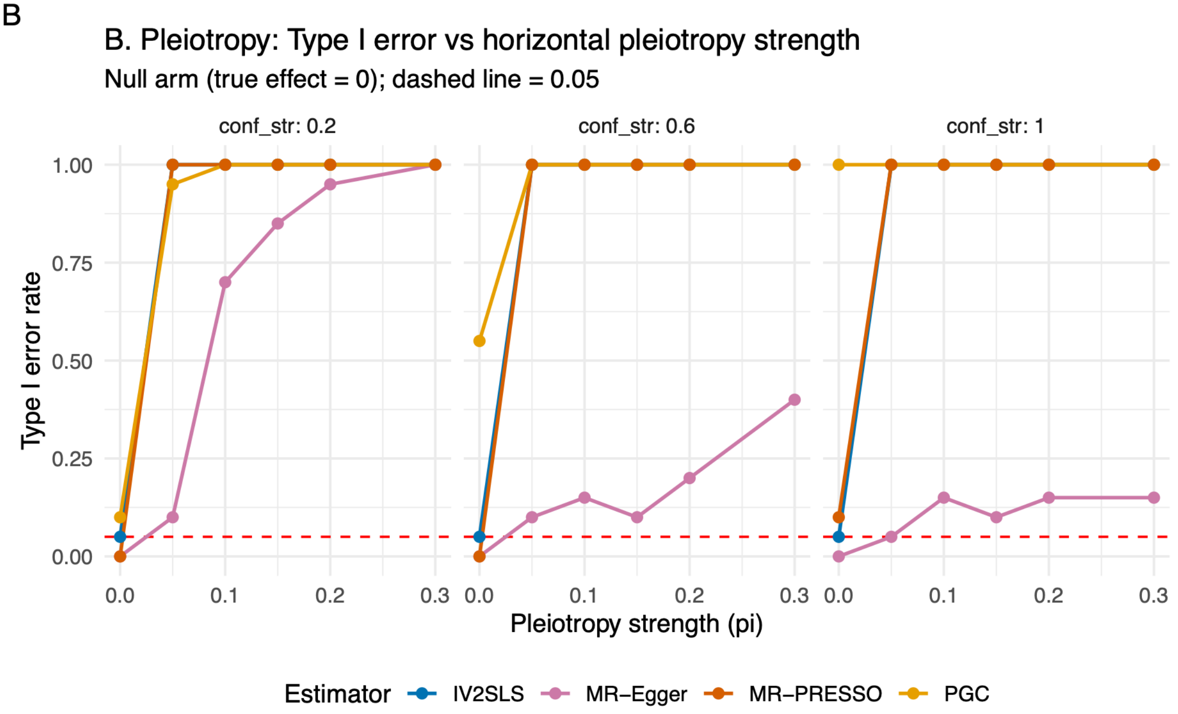


**Supplementary Figure S14. Pleiotropy benchmark: ICONIC vs. MR-Egger vs. MR-PRESSO (total effect).** Bias (alternative arm, true $\tau = 0.25$) and Type I error (null arm, true $\tau = 0$) for ICONIC's IV2SLS and PGC estimators and the MR-Egger and MR-PRESSO estimators from TwoSampleMR/MendelianRandomization, across pleiotropy strength $\pi \in \{0, 0.05, 0.1, 0.15, 0.2, 0.3\}$ and confounding strength $\delta \in \{0.2, 0.6, 1.0\}$ (faceted by $\delta$). **(A)** Bias versus $\pi$. **(B)** Type I error versus $\pi$; red dashed line marks $\alpha = 0.05$. $n = 500$, 10 features, 50 replicates. Abbreviations: NDE (natural direct effect); NIE (natural indirect effect); NC (negative control); MR (Mendelian randomization); GAN (generative adversarial network); DGP (data-generating process); RMST (restricted mean survival time); FDR (false discovery rate); 2SLS (two-stage least squares); IVW (inverse-variance weighted); OLS (ordinary least squares).

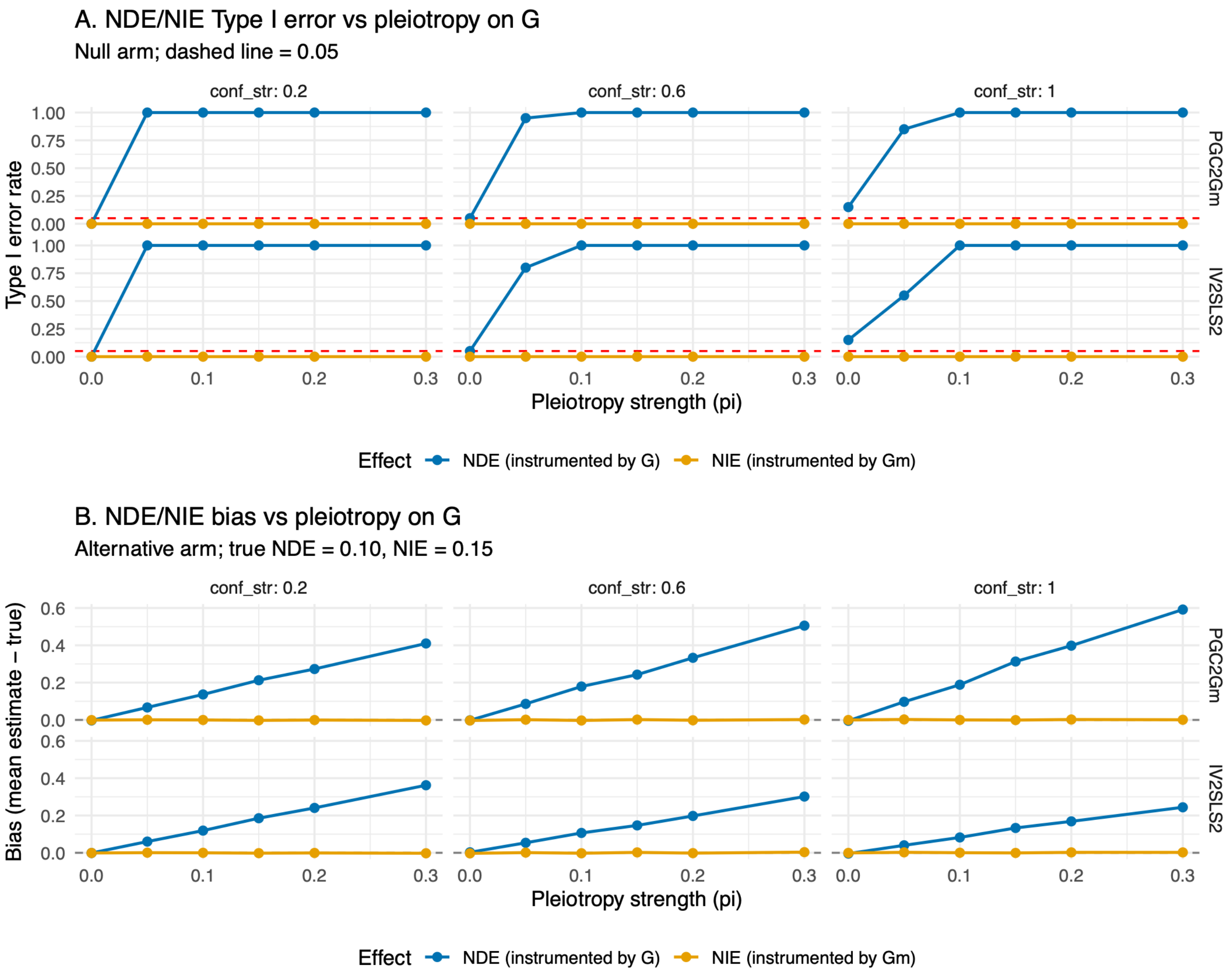


**Supplementary Figure S15. Pleiotropy benchmark: NDE/NIE decomposition (ICONIC two-instrument design).** Type I error (null arm) and bias (alternative arm) for the NDE and NIE estimates from ICONIC's two-instrument mediation estimators (PGC2Gm and IV2SLS2), across pleiotropy strength $\pi$ and confounding strength $\delta \in \{0.2, 0.6, 1.0\}$ (faceted by $\delta$, rows by method). **(A)** NDE and NIE Type I error versus $\pi$; red dashed line marks $\alpha = 0.05$. **(B)** NDE and NIE bias versus $\pi$. True NDE = 0.10, true NIE = 0.15. $n = 500$, 10 features, 50 replicates. Abbreviations: NDE (natural direct effect); NIE (natural indirect effect); NC (negative control); MR (Mendelian randomization); GAN (generative adversarial network); DGP (data-generating process); RMST (restricted mean survival time); FDR (false discovery rate); 2SLS (two-stage least squares); IVW (inverse-variance weighted); OLS (ordinary least squares).

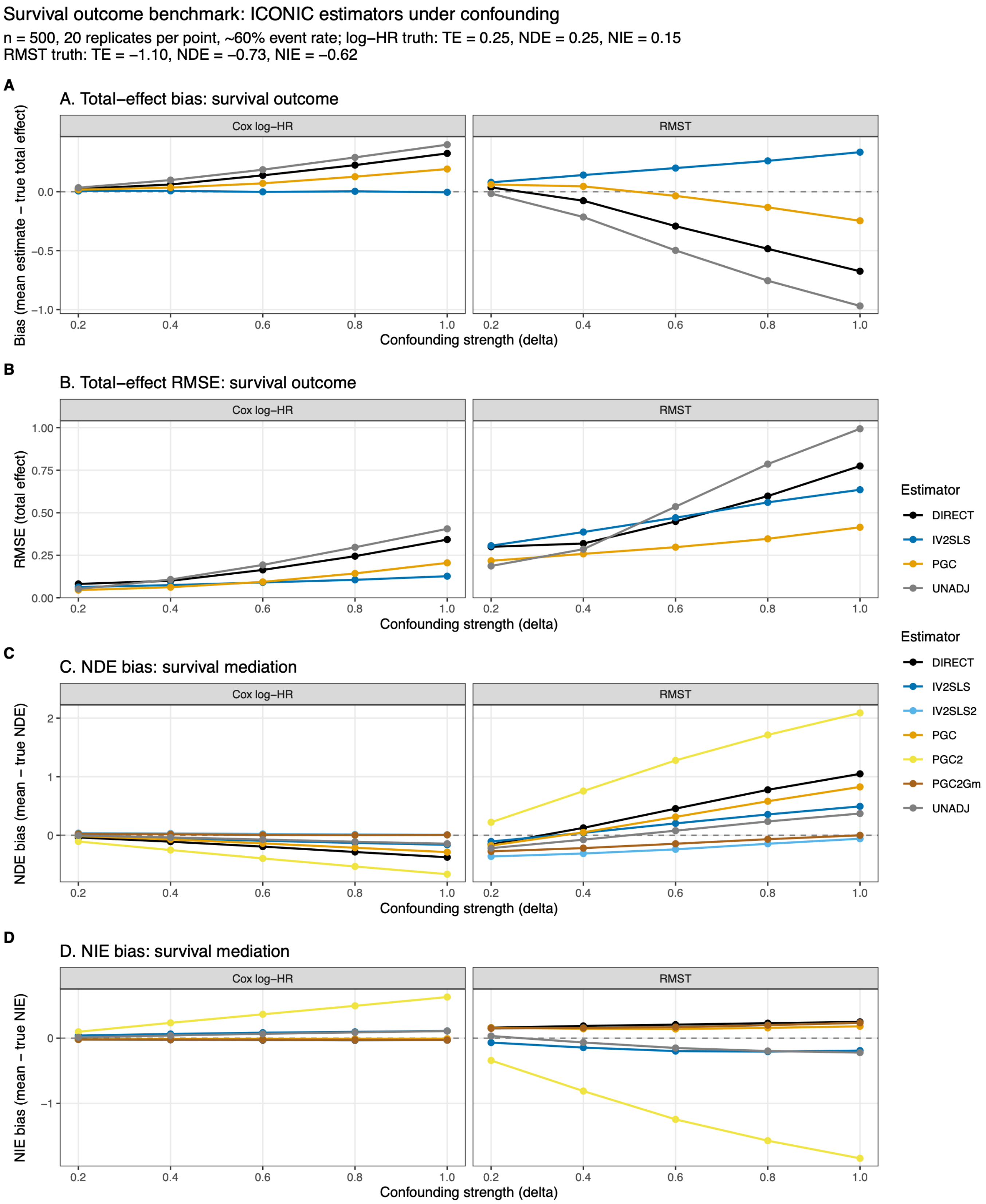


**Supplementary Figure S16. Survival outcome benchmark: ICONIC estimators under confounding.** All estimators except COCA (which is structurally incompatible with survival outcomes) are evaluated on a time-to-event outcome generated by converting the linear predictor of the structural causal model to time-to-event via an exponential proportional-hazards model with $\sim 60\%$ event rate. **(A)** Total-effect bias (mean estimate − true total effect) versus confounding strength $\delta$ for the four eligible total-effect estimators (UNADJ, DIRECT, IV2SLS, PGC), faceted by effect scale (Cox log-HR vs. RMST). **(B)** Total-effect RMSE versus $\delta$, same faceting. **(C)** NDE bias versus $\delta$ for all seven eligible mediation estimators (UNADJ, DIRECT, IV2SLS, IV2SLS2, PGC, PGC2, PGC2Gm), faceted by effect scale. **(D)** NIE bias versus $\delta$, same estimators and faceting. On the Cox log-HR scale (left facets), the true total effect is $\tau = 0.25$, true NDE $= 0.25$, and true NIE $= 0.15$. On the RMST scale (right facets), the truth is computed empirically at zero confounding with $n = 2{,}000$: true TE $= -1.0955$, true NDE $= -0.7284$, true NIE $= -0.6198$. Dashed grey lines mark zero bias. Settings: $n = 500$, 50 replicates per cell, $k = 2$ confounders with distinct path loadings, $\varphi = 0.8$, $\rho_{G1} = 0.3$, `mo_confounding` $= 0.8$, $\omega_1 = \omega_2 = 0.7$, matching the continuous mediation benchmark. Abbreviations: NDE (natural direct effect); NIE (natural indirect effect); NC (negative control); MR (Mendelian randomization); GAN (generative adversarial network); DGP (data-generating process); RMST (restricted mean survival time); FDR (false discovery rate); 2SLS (two-stage least squares); IVW (inverse-variance weighted); OLS (ordinary least squares).

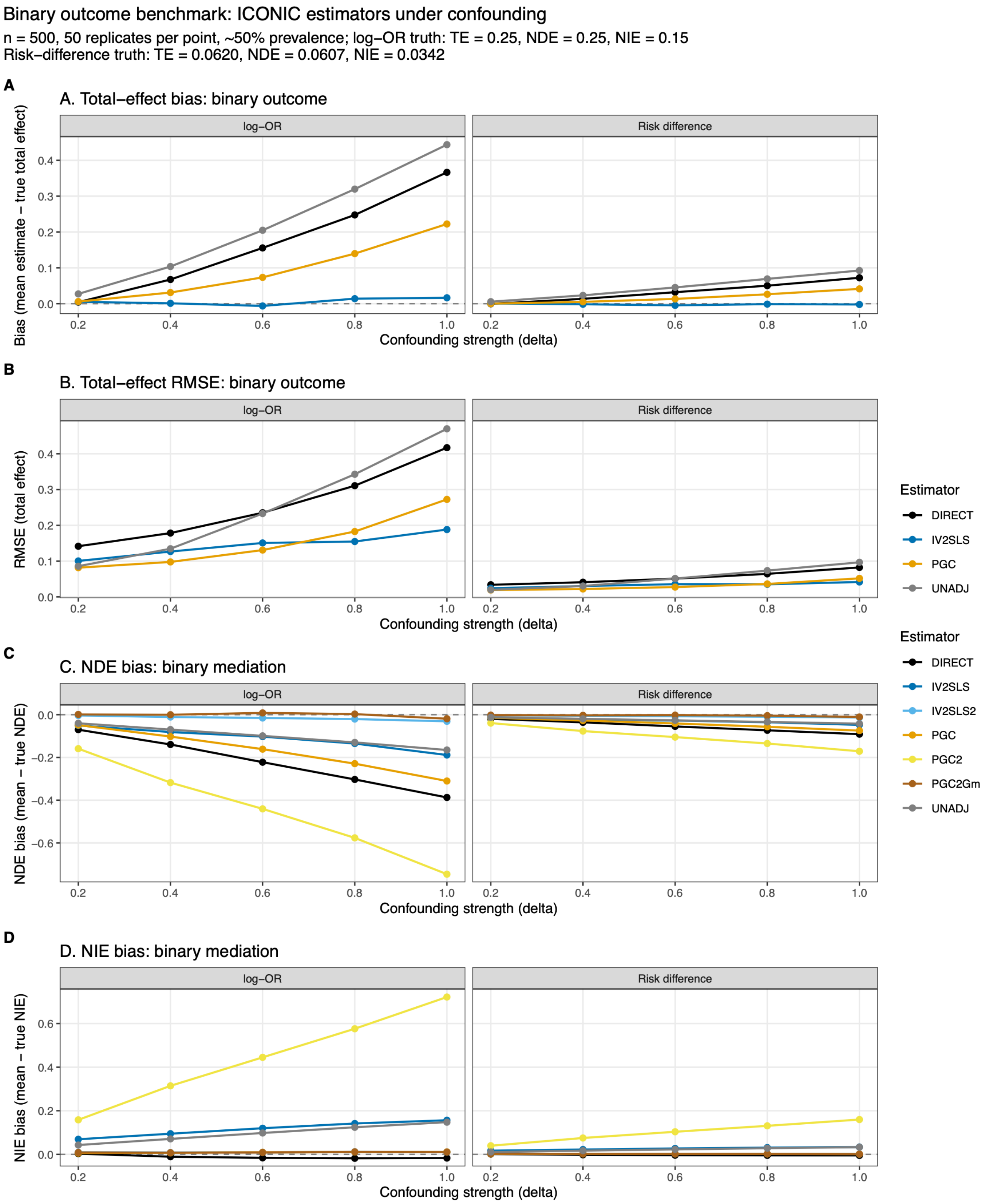


**Supplementary Figure S17. Binary outcome benchmark: ICONIC estimators under confounding.** All estimators except COCA (which is structurally incompatible with binary outcomes) are evaluated on a binary 0/1 outcome drawn from the linear predictor of the structural causal model via a logistic model calibrated to $\sim 50\%$ prevalence. **(A)** Total-effect bias (mean estimate $-$ true total effect) versus confounding strength $\delta$ for the four eligible total-effect estimators (UNADJ, DIRECT, IV2SLS, PGC), faceted by effect scale (logistic log-OR vs. linear-probability risk difference). **(B)** Total-effect RMSE versus $\delta$, same faceting. **(C)** NDE bias versus $\delta$ for all seven eligible mediation estimators (UNADJ, DIRECT, IV2SLS, IV2SLS2, PGC, PGC2, PGC2Gm), faceted by effect scale. **(D)** NIE bias versus $\delta$, same estimators and faceting. On the log-odds-ratio scale (left facets), the true total effect is $\tau = 0.25$, true NDE $= 0.25$, and true NIE $= 0.15$. On the risk-difference scale (right facets), the truth is computed empirically at zero confounding with $n = 2{,}000$: true TE $= 0.0620$, true NDE $= 0.0607$, true NIE $= 0.0342$. Dashed grey lines mark zero bias. Settings: $n = 500$, 50 replicates per cell, five confounding levels $\delta \in \{0.2, 0.4, 0.6, 0.8, 1.0\}$, $k = 2$ confounders with distinct path loadings, $\varphi = 0.8$, $\rho_{G1} = 0.3$, $\texttt{mo\_confounding} = 0.8$, $\omega_1 = \omega_2 = 0.7$, matching the continuous and survival mediation benchmarks. Abbreviations: log-OR (log-odds ratio); RD (risk difference); LPM (linear probability model); NDE (natural direct effect); NIE (natural indirect effect); NC (negative control); MR (Mendelian randomization); GAN (generative adversarial network); DGP (data-generating process); RMST (restricted mean survival time); FDR (false discovery rate); 2SLS (two-stage least squares); IVW (inverse-variance weighted); OLS (ordinary least squares).

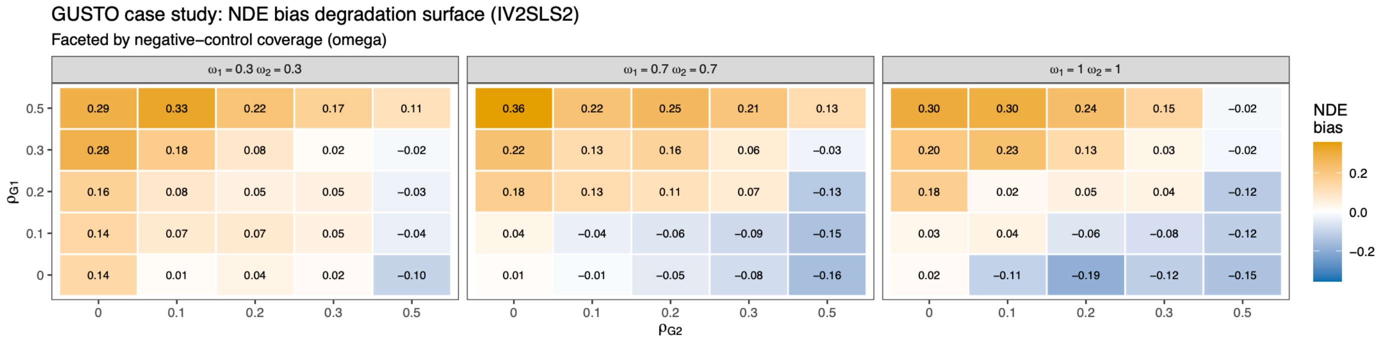


**Supplementary Figure S18. GUSTO case study: degradation surface for the recommended estimator.** NDE bias across the $\rho_{G1} \times \rho_{G2}$ grid of instrument-exogeneity violations for IV2SLS2 (the recommended estimator), calibrated to the GUSTO data texture by the generative texture model (GAN + copula). The surface is faceted by negative-control coverage ($\omega \in \{0.3,0.7,1.0\}$). The grid spans $\rho_{G1}, \rho_{G2} \in \{0,0.1,0.2,0.3,0.5\}$. Neither $\rho_{G1}$ nor $\rho_{G2}$ can be estimated from data (confounders are unobserved); the surface maps the plausible range calibrated to the user's cohort. Abbreviations: NDE (natural direct effect); NIE (natural indirect effect); NC (negative control); MR (Mendelian randomization); GAN (generative adversarial network); DGP (data-generating process); RMST (restricted mean survival time); FDR (false discovery rate); 2SLS (two-stage least squares); IVW (inverse-variance weighted); OLS (ordinary least squares).

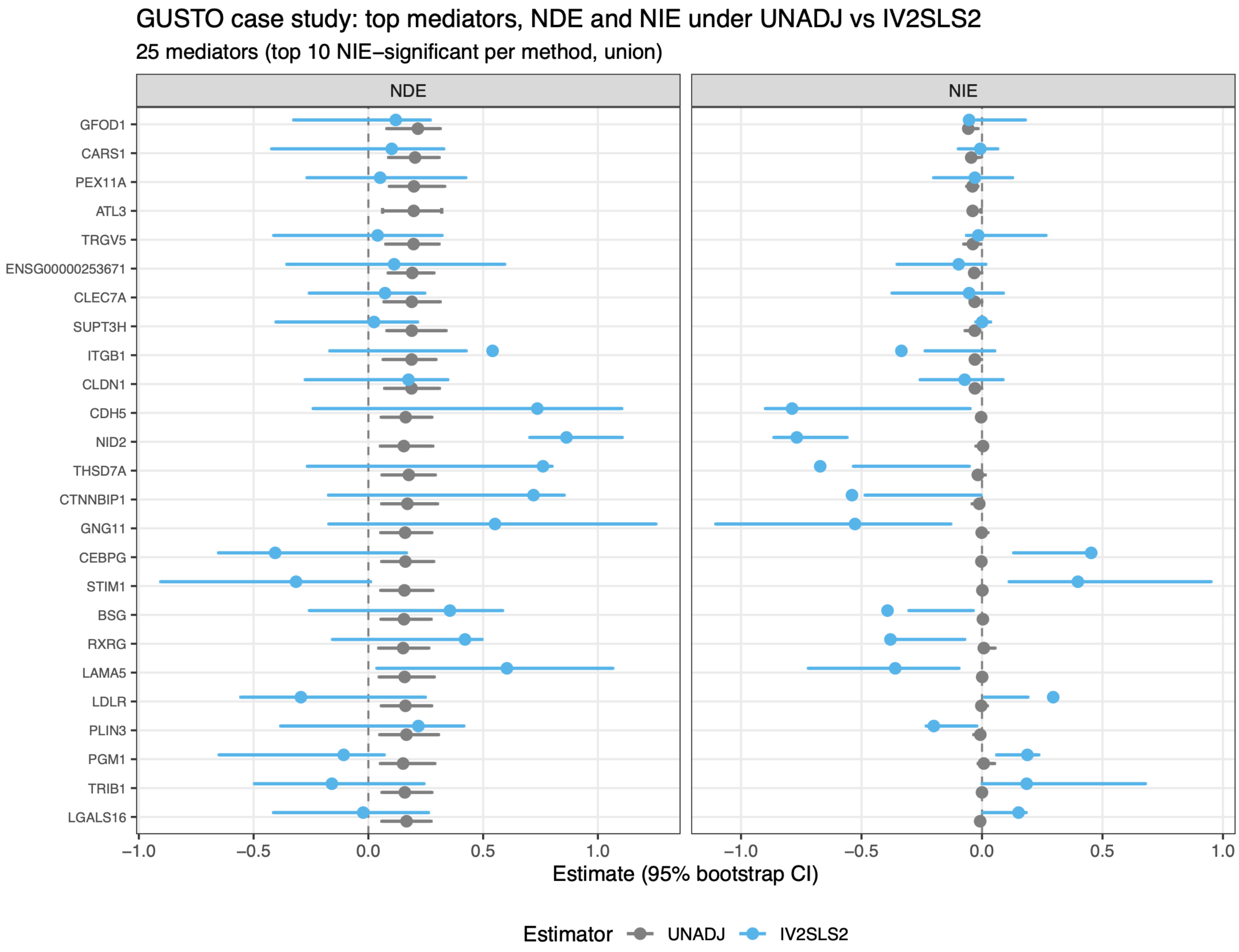


**Supplementary Figure S19. GUSTO case study: forest plot of NDE and NIE for the top significant mediators per method by NIE (union across UNADJ and IV2SLS2, 25 genes).** Point estimates with bootstrap 95% confidence intervals for the natural direct effect (NDE) and natural indirect effect (NIE) of maternal gestational diabetes on birth weight through placental genes. UNADJ estimates shown in grey; IV2SLS2 estimates shown in blue. Abbreviations: NDE (natural direct effect); NIE (natural indirect effect); NC (negative control); MR (Mendelian randomization); GAN (generative adversarial network); DGP (data-generating process); RMST (restricted mean survival time); FDR (false discovery rate); 2SLS (two-stage least squares); IVW (inverse-variance weighted); OLS (ordinary least squares).

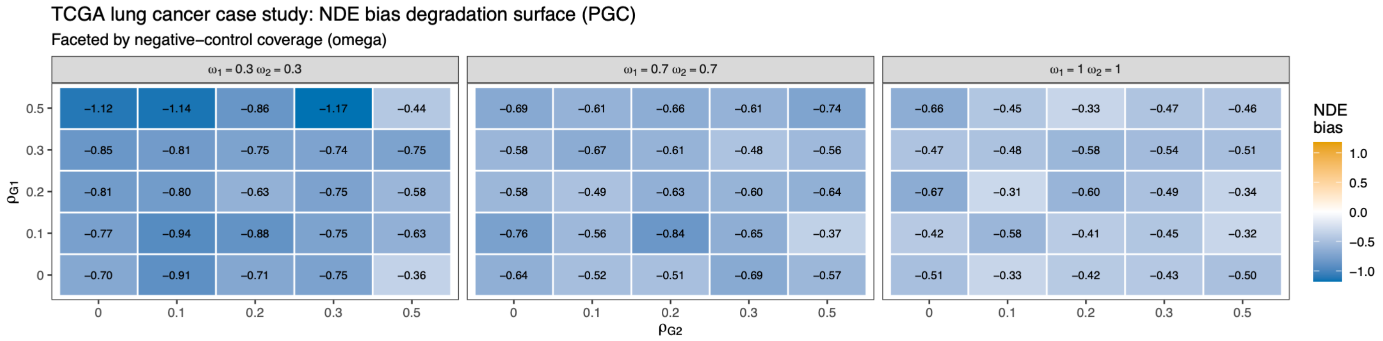


**Supplementary Figure S20. TCGA lung cancer case study: degradation surface for the recommended estimator.** NDE bias across the $\rho_{G1} \times \rho_{G2}$ grid of instrument-exogeneity violations for PGC (the recommended estimator). Because `infer_confounding()` does not support survival outcomes, the surface was calibrated to ICONIC's default survival texture rather than to the cohort's learned texture ($\varphi = 0.5$, $\texttt{mo\_confounding} = 0.8$). The surface is faceted by negative-control coverage ($\omega \in \{0.3, 0.7, 1.0\}$). The grid spans $\rho_{G1}, \rho_{G2} \in \{0, 0.1, 0.2, 0.3, 0.5\}$. Neither $\rho_{G1}$ nor $\rho_{G2}$ can be estimated from data (confounders are unobserved); the surface maps the plausible range. Abbreviations: NDE (natural direct effect); NIE (natural indirect effect); NC (negative control); MR (Mendelian randomization); GAN (generative adversarial network); DGP (data-generating process); RMST (restricted mean survival time); FDR (false discovery rate); 2SLS (two-stage least squares); IVW (inverse-variance weighted); OLS (ordinary least squares).

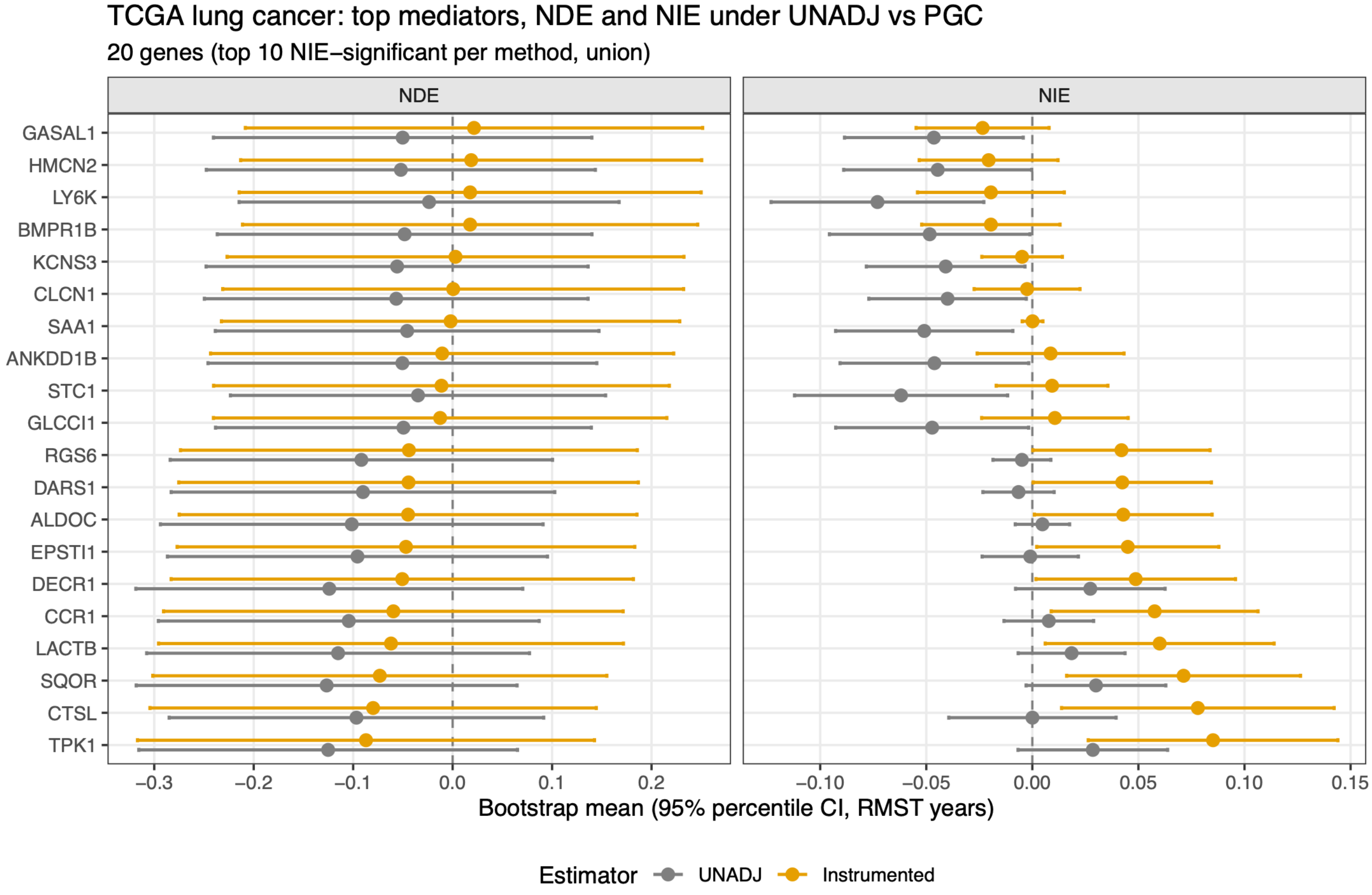


**Supplementary Figure S21. TCGA lung cancer case study: bootstrap forest of NDE and NIE for the top 10 significant mediators per method by NIE (union across UNADJ and the recommended estimator PGC, 20 genes).** Bootstrap mean estimates with percentile 95% confidence intervals for the natural direct effect (NDE) and natural indirect effect (NIE) of smoking on lung cancer survival through the top mediator genes, on the restricted mean survival time (RMST) scale in years (the bootstrap was run on the RMST scale; the Arm 3 results in the text are reported on the Cox log-hazard-ratio scale). UNADJ estimates shown in grey; PGC estimates shown in blue (“Instrumented”). Abbreviations: NDE (natural direct effect); NIE (natural indirect effect); NC (negative control); MR (Mendelian randomization); GAN (generative adversarial network); DGP (data-generating process); RMST (restricted mean survival time); FDR (false discovery rate); 2SLS (two-stage least squares); IVW (inverse-variance weighted); OLS (ordinary least squares).

### Data-Calibrated Simulation Mode; Generative Texture Pipeline

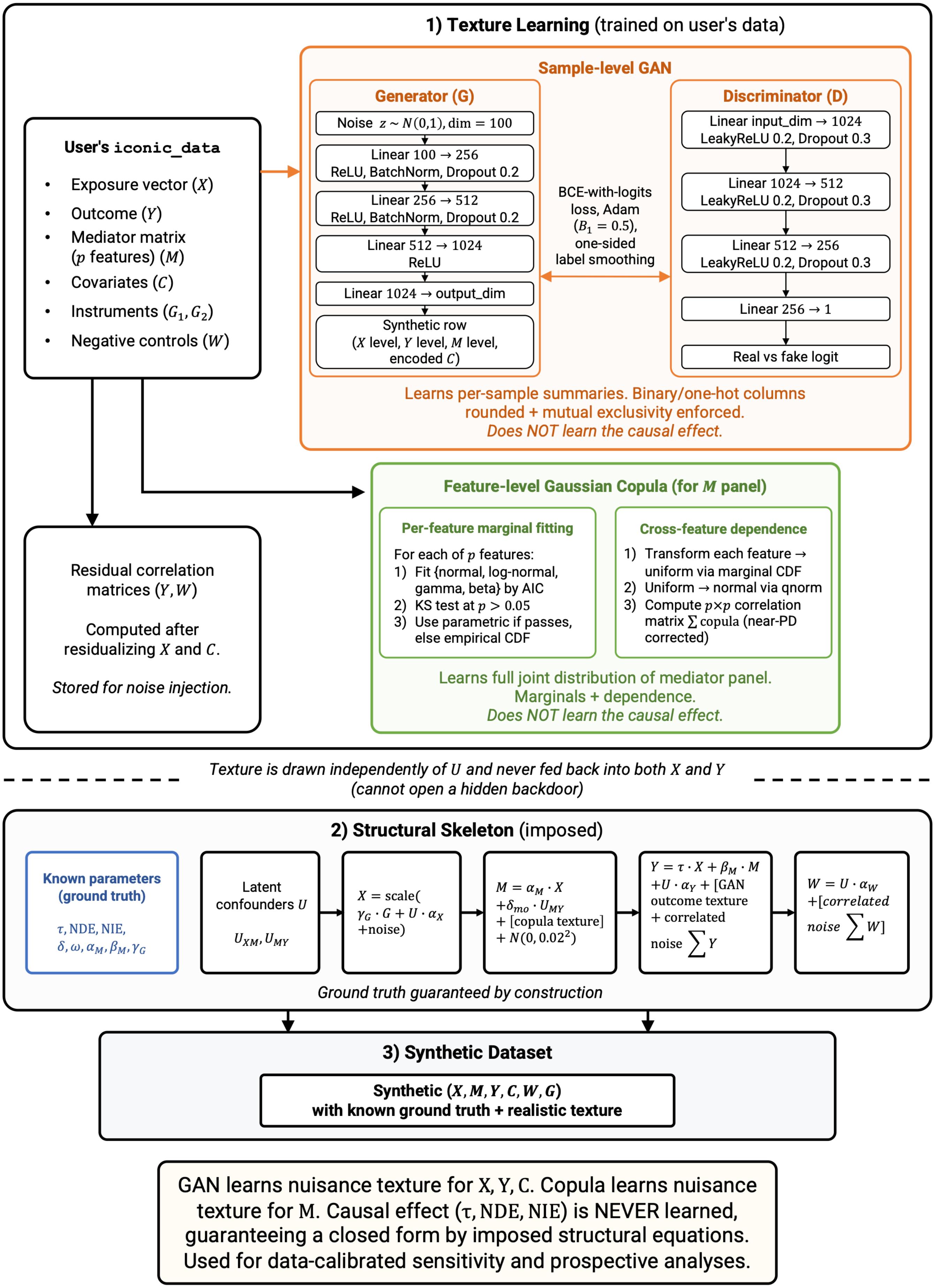

**Supplementary Figure S22. Data-calibrated simulation mode: the hybrid GAN-plus-copula generative texture pipeline.** Schematic of the data-calibrated generator used for sensitivity and prospective analyses **(Supplementary Methods S2)**. **(1) Texture learning (trained on the user's data).** From the user's `iconic_data` object, which contains exposure $X$, outcome $Y$, mediator matrix $M$ ($p$ features), covariates $C$, instruments $G_1, G_2$, and negative controls $W$, two components are trained independently, and neither learns the causal effect. A sample-level GAN (MLP generator and discriminator, non-saturating BCE-with-logits objective, Adam with $\beta_1 = 0.5$, one-sided label smoothing) learns the joint distribution of the per-sample exposure, outcome, and mediator levels and the encoded covariates, with binary/one-hot columns rounded and mutual exclusivity enforced. A feature-level Gaussian copula learns the full joint distribution of the mediator panel. Each feature's marginal is fit by the best-AIC parametric family (normal, log-normal, gamma, or beta) when that fit passes a Kolmogorov-Smirnov test at $p > 0.05$, falling back to the empirical CDF otherwise, and cross-feature dependence is captured by the correlation matrix of the normal-score-transformed marginals. Residual correlation matrices, computed after residualizing on $X$ and $C$, are stored for injection into the outcome and negative-control noise panels. **(2) Structural skeleton (imposed).** The learned texture is injected into the same structural causal model as the benchmark mode, but drawn independently of the latent confounders $U$ so it cannot open a hidden backdoor; the structural equations combine the imposed causal effect with GAN outcome texture, copula mediator texture, and correlated residual noise. **(3) Synthetic dataset.** The result is a synthetic $(X, M, Y, C, G, W)$ dataset with realistic data texture and known ground truth. Because the generative model never models the joint of $(X, M, Y)$ and learns only nuisance texture, the ground-truth $\tau$, NDE, and NIE are guaranteed by construction and require no post-hoc validation. Abbreviations: NDE (natural direct effect); NIE (natural indirect effect); NC (negative control); MR (Mendelian randomization); GAN (generative adversarial network); DGP (data-generating process); RMST (restricted mean survival time); FDR (false discovery rate); 2SLS (two-stage least squares); IVW (inverse-variance weighted); OLS (ordinary least squares).

### Supplementary Tables

**Supplementary Table S1. Scenario manifest for the simulation benchmarks.** Exported by `scenario_manifest()`. The manifest records the estimands (ground truth) and distinguishes modifiable parameters (swept across a grid) from fixed parameters (held constant), so the analyst can verify exactly what was varied and what was held fixed in each benchmark.

| Component | Parameter | Value(s) | Role |
|---|---|---|---|
| Estimands | True NDE ($\beta_X$) | 0.10 (mediation) / 0.25 (total-effect) | Ground truth |
| | True NIE ($\alpha_M \cdot \beta_M$) | 0.15 (per-mediator) | Ground truth |
| | True total effect ($\tau$) | 0.25 | Ground truth |
| Modifiable | Confounding strength ($\delta$) | $\{0.2,0.4,0.6,0.8,1.0\}$ | Swept |
| | Sample size ($n$) | $\{100,200,500,1000\}$ | Swept |
| | Instrument exogeneity ($\rho_{G1}, \rho_{G2}$) | $\{0,0.1,0.2,0.3,0.5\}$ | Swept (degradation surface) |
| | Feature correlation (`feat_cor`) | $\{0,0.2,0.4,0.6,0.8\}$ | Swept (Supplementary Figure S7) |
| | $U$ strength profile (`u_strength`) | uniform, moderate, extreme | Swept (Supplementary Figure S9) |
| | $W$ coverage profile (`w_coverage_profile`) | uniform, moderate, sparse | Swept (Supplementary Figure S9) |
| Fixed | M-O confounding ($\delta_{mo}$) | 0.8 | Held fixed |
| | Mediator-instrument strength ($\phi$) | 0.8 | Held fixed |
| | NC coverage ($\omega_1, \omega_2$) | 0.7 | Held fixed |
| | Latent confounders ($k$) | 2 | Held fixed |
| | Path loadings ($\lambda_{XM}, \lambda_{MY}$) | $e_1, e_2$ (distinct) | Held fixed |
| | Outcome features ($p$) | 10 | Held fixed |
| | Mediators | 1 | Held fixed |
| | Instrument strength ($\gamma_G$) | 0.6 ($F \geq 10$) | Held fixed |
| | SE method | `"delta"` (Sobel) | Default for benchmarks |
| | CI construction | Wald ($\pm 1.96 \times \mathrm{SE}$) | Coverage assessed at 95% |
| | Bootstrap resamples | 500 | For `se_method = "bootstrap"` comparisons |

| Component | Parameter | Value(s) | Role |
|---|---|---|---|
| Survival-specific | Outcome type | Continuous / Survival (time-to-event) | Survival benchmark (Supplementary Figure S16) |
| | Effect scale (survival) | Cox log-HR / RMST | Both scales benchmarked |
| | Event rate | 0.6 ($\sim 60\%$ events) | Held fixed |
| | Survival true TE (log-HR) | 0.25 | Ground truth (log-HR scale) |
| | Survival true NDE (log-HR) | 0.25 | Ground truth (log-HR scale) |
| | Survival true NIE (log-HR) | 0.15 | Ground truth (log-HR scale) |
| | Survival RMST truth | TE $= -1.0955$, NDE $= -0.7284$, NIE $= -0.6198$ | Empirical ($n = 2{,}000$, zero confounding) |
| Binary-specific | Outcome type | Continuous / Binary (0/1) | Binary benchmark (Supplementary Figure S17) |
| | Effect scale (binary) | Logistic log-OR / linear-probability risk difference | Both scales benchmarked |
| | Outcome prevalence | 0.5 ($\sim 50\%$ events) | Held fixed |
| | Binary true TE (log-OR) | 0.25 | Ground truth (log-OR scale) |
| | Binary true NDE (log-OR) | 0.25 | Ground truth (log-OR scale) |
| | Binary true NIE (log-OR) | 0.15 | Ground truth (log-OR scale) |
| | Binary risk-difference truth | TE $= 0.0620$, NDE $= 0.0607$, NIE $= 0.0342$ | Empirical ($n = 2{,}000$, zero confounding) |

### Supplementary Methods

#### Supplementary Methods S1. Time-to-event (survival) outcome support

ICONIC extends all estimators except COCA to time-to-event outcomes via two-stage predictor substitution (2SPS). The first-stage regressions (instrumenting $X$ on $G_1 + W + C$, instrumenting $M$ on $\hat{X} + G_m + W + C$, and constructing the bridge proxies $\widehat{W}$) involve only continuous variables ($X$, $M$, $W$) as responses, so they remain ordinary least squares regardless of the outcome type. Only the final outcome stage changes. The outcome model replaces OLS on a continuous $Y$ with a Cox proportional-hazards regression on the `Surv(time, event)` object (log-hazard-ratio scale) or OLS on restricted mean survival time (RMST) pseudo-observations (RMST scale). This 2SPS architecture preserves the identifying structure of each estimator (the instrument purges confounded variation in $X$, the bridge proxies absorb the residual confounding) while accommodating the censored, non-negative, skewed distribution of survival times.

*Two effect scales.*

ICONIC provides two effect scales for survival outcomes. The Cox log-hazard-ratio (log-HR) scale fits `coxph(Surv(time, event) ~ X_hat + M_hat + W_hat + C)` and reports the coefficients as log-HRs. The log-HR is non-collapsible. The conditional log-HR given covariates differs from the marginal log-HR even under no confounding, so the product decomposition NIE $= \hat{\alpha}_M \cdot \hat{\beta}_M$ is approximate on this scale because the product of two conditional log-HRs does not equal the marginal log-HR. The approximation error grows with the magnitude of the conditional log-HRs and does not vanish with sample size, so NDE + NIE need not equal the total effect on the log-HR scale when effects are large. An accelerated failure time (AFT) outcome stage would avoid this approximation entirely, because the AFT model is linear on the log-time scale and the product-of-coefficients decomposition is exact there; it can be substituted for the Cox outcome stage within the same 2SPS architecture and is a planned extension (see the main-text Future directions). The RMST scale first computes pseudo-observations via the leave-one-out jackknife of the Kaplan-Meier RMST estimate (Graw et al. 2009), then fits OLS on the pseudo-observations. The default truncation time $\tau$ is the 90th percentile of observed follow-up times. RMST is collapsible, so the NDE/NIE decomposition is exact on this scale; however, the pseudo-observations introduce additional variance relative to the Cox partial-likelihood estimator, producing wider confidence intervals and higher RMSE in the benchmarks **(Supplementary Figure S16)**. The log-HR scale is recommended as the primary effect scale; RMST provides a collapsible complement when marginal effects are of interest.

*COCA incompatibility.*

COCA regresses the negative control on the outcome ($W \sim Y + X$) and recovers $\hat{\tau} = -\hat{\beta}_X/\hat{\beta}_Y$. This requires $Y$ as a continuous response variable. With a censored time-to-event outcome, $Y$ is not available as a continuous variable (the outcome is represented as a `Surv(time, event)` object), so COCA is structurally incompatible with survival outcomes. ICONIC returns `NA` for COCA on survival data with a `reason` attribute explaining the incompatibility, and the estimator is excluded from the eligibility report.

*Stability guard.*

The PGC2 and PGC2Gm survival mediation estimators involve bridge-matrix inversion at two stages, which can produce numerically unstable coefficient estimates when the bridge matrix is near-singular. ICONIC implements a stability guard that rejects coefficient estimates with $|\hat{\beta}| > 10$ (a threshold chosen well above any plausible log-HR or RMST effect), returning `NA` so the unstable estimate does not propagate into the NDE/NIE decomposition. This guard prevents the extreme RMSE values (4–150) observed in earlier implementations from corrupting the benchmark results.

*Interface.*

Survival outcomes are specified at `iconic_data()` construction: `iconic_data(outcome_type = "survival", surv_time = time_vector, surv_event = event_indicator, ...)`. The effect scale is selected at estimation: `iconic_estimate(effect_scale = "loghr")` (default) or `iconic_estimate(effect_scale = "rmst", tau = truncation_time)`. When `tau` is not specified, it defaults to the 90th percentile of observed follow-up times. All downstream functions (`iconic_diagnose()`, `iconic_sensitivity()`, `iconic_prospect()`, `iconic_recommend()`) operate identically on survival and continuous data; the diagnostic screens (instrument strength, NC validity, completeness) are unchanged because they depend only on the first-stage regressions, which remain OLS.

*Benchmark design.*

The survival benchmark uses the same structural DGP as the continuous benchmark (`generate_toy_data`), with the linear predictor $\eta = \beta_X X + \alpha_M \beta_M M_{\text{path}}$ converted to time-to-event via an exponential proportional-hazards model: $T = -\log(U)/\lambda(\eta)$ where $U \sim \text{Uniform}(0,1)$ and $\lambda(\eta) = \lambda_0 \exp(\eta)$. The baseline hazard is fixed at $\lambda_0 = 0.1$, and the rate of the independent exponential censoring distribution is calibrated to achieve a target event fraction of $\sim 60\%$ (censor rate $= \bar{\lambda}(1 - 0.6)/0.6$, where $\bar{\lambda}$ is the mean hazard). The total-effect sweep uses $\alpha_M = 0$, $\beta_M = 0$ (no mediation path), so the true total effect on the log-HR scale is $\beta_X = 0.25$. The mediation sweep uses $\alpha_M = 0.50$, $\beta_M = 0.30$ (true NDE $= 0.25$, true NIE $= 0.15$ on the log-HR scale), with $k = 2$ confounders under distinct path loadings, `mo_confounding` $= 0.8$, $\rho_{G1} = 0.3$, $\omega_1 = \omega_2 = 0.7$, matching the continuous mediation benchmark. The RMST truth is not analytically tractable because it depends on the baseline hazard and truncation time; it is computed empirically by running the UNADJ estimator at zero confounding with $n = 2{,}000$, yielding TE $= -1.0955$, NDE $= -0.7284$, NIE $= -0.6198$. Fifty replicates per cell, $n = 500$, five confounding levels $\delta \in \{0.2, 0.4, 0.6, 0.8, 1.0\}$, and both effect scales are benchmarked **(Supplementary Figure S16)**.

**Supplementary Methods S2. Generative pipeline architecture**

*Estimator regression equations.*

This section gives the full stage-by-stage regression equations for all eight ICONIC estimators referenced in the main text. Throughout, $X$ is the exposure, $M$ the mediator (possibly a matrix of $p$ features), $Y$ the outcome, $G_1$ the exposure instrument, $G_m$ the mediator instrument, $W$ the negative-control panel proxying the path-specific confounder composites $U_{XM} = U^\top \lambda_{XM}$ and $U_{MY} = U^\top \lambda_{MY}$, and $C$ the covariates. Hatted quantities ($\hat{X}$, $\widehat{M}$, $\widehat{W}$) denote first-stage fitted values. The total effect is $\tau$, the natural direct effect $\mathrm{NDE} = \beta_X$, and the natural indirect effect $\mathrm{NIE} = \alpha_M \beta_M$ under the Baron-Kenny product-of-coefficients decomposition.

**Total-effect estimators** (no mediator; $\tau = \beta_X$ in a model with $M$ absent):

- **UNADJ** (confounded reference): $\hat{\tau} = \hat{\beta}_X$ from $Y \sim X + C$. No identifying assumption; biased by unmeasured confounding.

- **DIRECT**: $\hat{\tau} = \hat{\beta}_X$ from $Y \sim X + G_1 + W + C$. Uses all observables but does not remove unmeasured confounding; structurally biased.

- **COCA** (Control Outcome Calibration Approach): fit $W \sim Y + X + C$, recover $\hat{\tau} = -\hat{\beta}_X / \hat{\beta}_Y$. Efficient when $W$ is a strong proxy of $U$; unstable when $|\hat{\beta}_Y| \approx 0$.

- **IV2SLS** (two-stage least squares): stage 1, $\hat{X} = \hat{\gamma}_G G_1 + \hat{\gamma}_C C$ from $X \sim G_1 + C$; stage 2, $\hat{\tau} = \hat{\beta}_X$ from $Y \sim \hat{X} + C$. Requires $G_1 \perp U$ and a first-stage partial $F \geq 10$ (the squared $t$-statistic of the $G_1$ coefficient). When a panel $W$ is supplied it enters as an optional proximal augmentation, conditioned on in both stages ($X \sim G_1 + W + C$; $Y \sim \hat{X} + W + C$).

- **PGC** (proximal g-computation, three stages): stage 1, residualize $r_X = X - \hat{\gamma}_G G_1 - \hat{\gamma}_C C$; stage 2, fit the bridge by regressing the residual on the full panel, $r_X \sim W_1 + \cdots + W_q + C$, and take $\widehat{W}$ as its fitted values (a scalar confounder proxy); stage 3, $\hat{\tau} = \hat{\beta}_X$ from $Y \sim X + \widehat{W} + C$. Requires completeness $\dim(W_{\text{valid}}) \geq k$. (A legacy scalar variant, `fit_pgc_scalar()`, instead regresses a single control $w$ on $r_X + C$; it is algebraically equivalent to IV/2SLS and the completeness condition is not binding for it.)

Each total-effect estimator extends to mediation by replacing the outcome regression with the Baron-Kenny decomposition: fit $Y \sim \hat{X} + M + \widehat{W} + C$ (or the estimator-specific analogue), take $\mathrm{NDE} = \hat{\beta}_X$ and $\mathrm{NIE} = \hat{\alpha}_M \hat{\beta}_M$ where $\hat{\alpha}_M$ is the exposure coefficient from the mediator model $M \sim X + C$ (or its instrumented form).

**Mediation-specific estimators** (require $G_m$ or a negative-control panel $W$ covering each path's confounder composite):

- **IV2SLS2** (two-stage MR, three sequential 2SLS stages): stage 1a, $\hat{X} = \hat{\gamma}_G G_1 + \hat{\gamma}_{W1} W_1 + \hat{\gamma}_C C$ from $X \sim G_1 + W_1 + C$; stage 1b, $\widehat{M} = \hat{\pi}_X \hat{X} + \hat{\pi}_{Gm} G_m + \hat{\pi}_{W2} W_2 + \hat{\pi}_C C$ from $M \sim \hat{X} + G_m + W_2 + C$; stage 2, fit $Y \sim \hat{X} + \widehat{M} + W_2 + C$, giving $\mathrm{NDE} = \hat{\beta}_X$ and $\mathrm{NIE} = \hat{\alpha}_M \hat{\beta}_M$. The path-specific panels are optional: $W_1$ (proxying the $X \to M$ confounder) enters only stage 1a, and $W_2$ (proxying the $M \to Y$ confounder) enters stages 1b and 2; with both omitted the estimator reduces to plain two-instrument 2-stage MR. A single pooled panel conditioned on in all three stages is a collider under multi-confounder designs and is not used; when $W_1$ and $W_2$ proxy the same latent composite (single-confounder design) the estimator falls back to plain MR. Requires both $G_1$ and $G_m$ valid with partial $F \geq 10$.

- **PGC2** (two-stage proximal mediation, no $G_m$): stage 1, after a partial-$F$ gate on $G_1$ in $X \sim G_1 + W_1 + C$, residualize $r_X = \mathrm{resid}(X \sim G_1 + C)$, fit the bridge $r_X \sim W_1 + C$ and take $\widehat{W}_X$ as its fitted values, then form $\hat{X} = \mathrm{fitted}\big(X \sim G_1 + \widehat{W}_X + C\big)$; stage 2, residualize $r_M = \mathrm{resid}\big(M \sim \hat{X} + C\big)$, fit the bridge $r_M \sim W_2 + C$ and take $\widehat{W}_M$ as its fitted values, then form $\widehat{M} = \mathrm{fitted}\big(M \sim \hat{X} + \widehat{W}_M + C\big)$ with $\hat{\alpha}_M$ the coefficient on $\hat{X}$; stage 3, fit $Y \sim \hat{X} + \hat{M} + \widehat{W}_X + \widehat{W}_M + C$, giving $\text{NDE} = \hat{\beta}_X$ and $\text{NIE} = \hat{\alpha}_M \hat{\beta}_M$. Requires completeness at both stages (each panel must span its path's confounder composite).

- **PGC2Gm** (negative-control-augmented): identical to PGC2 except in stage 2, where the mediator instrument isolates the confounder-driven variation in $M$: residualize $r_M = \text{resid}(M \sim G_m + C)$, fit the bridge $r_M \sim W_2 + C$ for $\widehat{W}_M$, and form $\hat{M} = \text{fitted}\left(M \sim \hat{X} + G_m + \widehat{W}_M + C\right)$; stage 3 is unchanged. Requires adequate $W$ coverage of both composites and a mediator instrument; residual bias from $G_m$ correlated with the $M \to Y$ composite is smaller than IV2SLS2 but nonzero.

Both PGC2-family bridges draw on the path-specific panels $W_1$ and $W_2$; each bridge is adequate to the extent that its panel covers that path's confounder composite (coverage $\omega_1$, $\omega_2$). A panel need not consist of a single confounder; it need only span its composite, which a multi-feature panel can achieve even when the two paths are confounded by distinct combinations of the $k$ latent confounders. (A single shared panel is the special case $W_1 = W_2 = W$.)

ICONIC derives the pooled single panel $W$ used by the single-panel estimators (DIRECT, COCA, PGC) and by the negative-control validity/completeness screens from whatever panels are supplied: as the average $(W_1 + W_2)/2$ when both path-specific panels are present, or as the lone panel itself ($W = W_2$ or $W = W_1$) when exactly one is supplied. Consequently, under a single outcome-side panel ($W_2$ only) DIRECT, COCA, and PGC remain eligible alongside UNADJ, IV2SLS, and IV2SLS2. The two-bridge estimators PGC2 and PGC2Gm require both $W_1$ and $W_2$ and stay gated under a lone panel unless the analyst sets `recycle_lone_panel = TRUE` in `iconic_data()`, which reuses the lone panel as both bridges (the shared-panel special case $W_1 = W_2$). This opt-in assumes the single panel is complete for *both* path confounder composites; it is off by default because IV2SLS2 is the more defensible primary estimator when coverage of the exposure-side ($X \to M$) composite is in doubt.

*Generative pipeline architecture.*

ICONIC employs two distinct simulation modes that serve different purposes. The benchmark simulations use the structural generator (`generate_toy_data`) with parametric Gaussian noise and no learned texture; their role is to validate the estimators under controlled confounding scenarios with known ground truth. The data-calibrated simulations use the full generative pipeline, which trains a texture model on the user's own data so that the sensitivity and prospective analyses are calibrated to realistic covariate, outcome, and mediator distributions. This separation is deliberate. Estimator validation requires a transparent, reproducible data-generating process whose parameters are known exactly, while the user-facing workflow benefits from realistic data texture even at the cost of a learned (and therefore less transparent) noise model.

Unlike simple Monte Carlo simulations,[1] ICONIC uses a hybrid generative texture model to create realistic synthetic datasets that preserve authentic covariate structures from real cohort data[2,3] while enabling systematic manipulation of confounding scenarios. This context-specific simulation approach, calibrating the data-generating process to the target study rather than relying on generic benchmarks, is needed for methods whose performance depends on the empirical distribution of the analysis data.[4] The design follows the plasmode approach[5] of learning realistic data texture from observed data while imposing an investigator-specified causal effect, and the Credence framework[6] of training a deep generative model to the empirical distribution with user-specified ground truth. A classical plasmode redraws new outcomes from a fitted system of parametric equations with added noise, preserving the fitted covariate-exposure-mediator structure while the analyst injects the causal effect; ICONIC's hybrid GAN-plus-copula approach serves the same purpose but relaxes the parametric assumption on the texture layer, learning the marginal and dependence structure of the mediator panel nonparametrically so that synthetic data retain realistic distributional shapes that a misspecified parametric fit would distort. A naive generative model trained directly on the full joint distribution of $(X, M, Y)$ cannot benchmark causal estimators, since it has no notion of the ground-truth effect $\tau$, the unobserved confounder $U$ is absent from observed data, and a generator optimized for distributional or predictive fidelity can preserve those qualities while distorting the causal-effect contrast.[7] This motivates the frugal parameterization principle,[8] which places the causal effect of interest at the centre of the model and learns only the surrounding nuisance texture from data.

**(1) The causal skeleton** implements the SCM above with $\tau$, NDE, NIE, confounding strength $\delta$, and negative-control coverage $\omega$ as known, tunable parameters.

**(2) The negative-control mechanism** is pluggable. Any function $\mathrm{nc\_model}(U, C,\cdot) \to W$ can be supplied. Two such functions are built-in: `nc_proxy`, where each control is a coverage-weighted mixture of the captured confounders plus noise (reducing to the classic single-proxy model when $k = 1$), and `nc_cpg`, which simulates spatially correlated CpG methylation values along the genome partly driven by $U$, then forms each control as a sparse linear prediction from the methylation. Both built-in models accept an optional noise correlation matrix that, when supplied, draws the noise component from a multivariate normal with the learned cross-feature correlation structure, so the negative controls retain realistic correlations conditional on the confounder.

**(3) The texture model** is a hybrid of two independently trained components, each chosen to match the dimensionality and structure of its target block. The first is a sample-level GAN. An MLP generator and discriminator are trained with a non-saturating binary-cross-entropy objective (BCE with logits for numerical stability), Adam optimization ($\beta_1 = 0.5$, following DCGAN stabilization), one-sided label smoothing, and per-column normalization.[9] The architecture follows the conditional tabular GAN (CTGAN) design[10] and the CopulaGAN philosophy[11] of combining copula-based and GAN-based modeling within a single tabular synthesizer. This GAN learns the joint distribution of the per-sample exposure level, outcome level, mediator level, and encoded covariates (one row per sample), supplying realistic covariate structure and outcome marginals while preserving the discrete (binary and one-hot) structure of encoded categorical covariates through post-hoc rounding and mutual-exclusivity enforcement. A GAN is used for this low-dimensional sample-level block rather than the copula used for the mediator because the block contains mixed continuous and categorical variables whose nonlinear interactions and mutual-exclusivity constraints are more naturally preserved by a generator with a flexible output layer than by a Gaussian copula on normal scores. The second component is a feature-level Gaussian copula model that learns the full joint distribution of the mediator ($M$) panel, following the copula-based omics simulation framework of scDesign3.[12] For each mediator feature, the default (`marginal_method = "auto"`) fits the parametric families (normal, log-normal, gamma, or beta) and uses the best-AIC fit when it passes a Kolmogorov-Smirnov goodness-of-fit test at $p > 0.05$, falling back to the empirical CDF otherwise; the user may override this automatic selection (`marginal_method = "empirical"` or `"parametric"`). Cross-feature dependence is captured by the correlation matrix of the normal-score-transformed marginals.

The negative-control ($W$) panel is not learned from data; it remains generated by the pluggable $\mathrm{nc\_model}$, with feature-level residual correlation matrices (computed after residualizing on exposure and covariates) injected as correlated noise so the controls retain realistic cross-feature correlations conditional on the confounder. Because these residual correlations still carry part of the confounder's cross-feature signature, injecting them alongside the synthetic confounder loadings partially double-counts $U$; the optional `residualize_on = "XCW"` setting in `load_real_input_data()` mitigates this by also residualizing the outcome-panel correlations on the $W$ bridge proxy (the first principal component of the $W$ panel), and is recommended only when the completeness-capture test reports "strong", since it can over-partial when $W$ is a weak proxy. The outcome ($Y$) panel similarly uses residual correlation matrices for its idiosyncratic noise. These correlation matrices govern only the outcome and negative-control noise panels; the cross-feature dependence of the mediator ($M$) panel is handled separately by the feature-level Gaussian copula described above. The toy simulation also supports a `feat_cor` parameter that injects block-diagonal correlated noise into the outcome and negative-control panels, modelling correlation structure of the kind observed in co-expression modules. Features are divided into $\lceil\sqrt{p}\rceil$ equal-sized modules with within-module pairwise correlation equal to `feat_cor` and between-module correlation of 0.

For multiple confounders, $U \in \mathbb{R}^{n \times k}$ with per-pathway multipliers (`MMExp`, `MMOut`, `MMCon`, `MMCpG`) scaling how strongly the confounders load into the exposure, outcome, controls, and methylation, respectively. This structural-anchored hybrid enables systematic benchmarking of causal inference methods under known ground truth on data that resembles the analyst's cohort.[13] It shares with the frugal-parameterization approach[8] and recent normalizing-flow extensions[14] this principle, and with controllable causal-sandbox frameworks[15] the goal of factorial manipulation of confounding scenarios over realistic data texture, but resolves the fidelity-versus-controllability tradeoff that those frameworks confront within a single learned objective by a different route. Because ICONIC's generative model never models the joint of $(X, M, Y)$ and only learns the nuisance texture, the ground-truth effect is guaranteed by construction and requires no post-hoc validation that the generator realized the specified effect.

The model-selection workflow functions (`iconic_sensitivity()` and `iconic_prospect()`) train the texture layer directly from the user's `iconic_data` object when no pre-trained model is supplied. The object consists of an exposure vector, outcome (scalar or matrix), mediator, covariates, and any instruments or negative controls. It is converted to the generative pipeline's input format and used to fit the generator. The sample-level GAN is trained on the per-sample summaries (exposure, outcome, mediator levels, and covariates), while the feature-level copula is trained on the full mediator matrix, learning each feature's marginal distribution and their cross-feature dependence. Although the GAN training frame contains exposure, outcome, and mediator summary columns, these columns are not carried into the synthetic causal variables: at generation time only the covariate columns and an exogenous outcome baseline (the scaled outcome-level draw) are used, and the synthetic exposure, mediator, and outcome are regenerated by the causal skeleton with the investigator-specified effects. Synthetic covariate distributions, outcome marginals, and the mediator panel's full feature-level structure (marginals and correlations) therefore match the user's cohort rather than defaulting to generic synthetic covariates. A pre-trained model may instead be attached at `iconic_data()` construction or passed via the trained_gan argument to avoid retraining across workflow steps.

**Supplementary Methods S3. Data-calibrated confounding parameters**

`infer_confounding()` estimates the held-fixed confounding parameters from the user's data, supplying a `confounding = "inferred"` mode for `iconic_sensitivity()` and `iconic_prospect()`. Confounding strength is inferred from the gap between the unadjusted OLS and IV2SLS estimates, averaged across features and scaled by the standard deviation of the exposure; mediator-outcome confounding from the gap between the IV2SLS and IV2SLS2 natural indirect effects; negative-control coverage from the square root of the $R^2$ obtained by regressing each negative-control feature on the outcome residualized on the exposure and covariates; and the number of latent confounders via parallel analysis on the correlation matrix of residualized outcomes. Parameters that cannot be inferred fall back to defaults with a warning. To keep calibration tractable on high-dimensional mediator panels, `infer_confounding()` estimates these parameters on a random subset of mediators (default 50, `max_infer_tasks`) rather than the full panel; `iconic_sensitivity(confounding = "inferred")` uses this subset directly, and a precomputed `iconic_confounding` object may be passed as the `confounding` argument to avoid re-running the inference. When the analyst supplies explicit sweep vectors for `omega_1`/`omega_2` (or a `mo_confounding` value), those user-supplied values take precedence over the inferred scalars, so the coverage facet of the degradation surface is preserved under `confounding = "inferred"`. The instrument-confounder correlations remain unestimable, since $U$ is unobserved, and are always swept. The inference uses estimator validity (e.g., that IV2SLS is unbiased) to calibrate a benchmark whose purpose is to test estimator validity, so inferred values should be read as best-case calibrations under the stated assumptions. The `confounding = "default"` mode avoids this circularity and remains the recommended starting point. For transparency, ICONIC enumerates the fixed defaults in each function's documentation (`@section Defaults`): `iconic_sensitivity()` and `iconic_prospect()` use `mo_confounding = 0.8`, `phi = 0.8` (or inferred from the diagnosis), and a shared confounder composite ($\lambda_{XM} = \lambda_{MY}$); `iconic_sensitivity()` sweeps negative-control coverage by default (`omega_1 = omega_2 = c(0.3, 0.7, 1.0)`, on the diagonal) jointly with the exogeneity grid, while `iconic_prospect()` holds `omega_1 = omega_2 = 0.7` for its Phase 1/2 strength surface and runs a Phase 3 robustness sweep by default (`run_rho_sweep = TRUE`) crossing `rho_G1_grid = rho_G2_grid = c(0, 0.1, 0.2, 0.3, 0.5)` with coverage `omega_grid_rho = c(0.3, 0.7, 1.0)` at the target instrument strength; `iconic_estimate()` uses `se_method = "delta"` with `n_boot = 500` available; `iconic_recommend()` uses `criterion = "combined"` and, when no sensitivity surface is supplied, runs `iconic_sensitivity()` automatically (`auto_sensitivity = TRUE`) so the ranking is robustness-based by default; and `nc_validity_screen()` uses `criterion = "both"` with `magnitude_threshold = 0.10` (a stricter threshold of 0.75 is used only in the COCA recount). The iconic_diagnose() and iconic_prospect() functions also accept an allow_no_proxy argument (default TRUE). When FALSE, the user must explicitly acknowledge that proceeding without negative controls limits the analysis to instrument-based estimators only.

The feature-level texture models learned by the generative pipeline (see **Supplementary Methods S2**) are distinct from the scalar coverage parameter $\omega$ estimated by infer_confounding(). The coverage parameter controls how strongly the negative controls capture the confounder signal, while the copula and residual correlation matrices control the cross-feature structure of the mediator and negative-control noise, respectively. Both are learned from the data but serve different roles in the simulation: $\omega$ scales the confounder-driven component of each control, while the copula shapes the full joint distribution of the mediator panel and the residual correlation matrix shapes the noise that remains in the negative-control panel after removing the confounder component.

#### Supplementary Methods S4. Inference and CI/SE calibration

Inference for the natural effects uses Wald tests throughout. The NDE p-value is the standard t-test for the exposure coefficient in the outcome regression, and the NIE p-value is a two-sided Wald test using a delta-method standard error (the Sobel test). ICONIC also supports a nonparametric bootstrap standard error (se_method = "bootstrap", default n_boot = 500 resamples) for estimators such as COCA whose delta-method variance is inflated by near-singular ratio terms; the bootstrap SE is more honest for these estimators and is reported alongside the delta-method SE in the model-selection workflow. No bootstrap or permutation-based inference is applied in the simulation benchmarks except where explicitly noted.

Confidence intervals for the NDE and the NIE are Wald intervals, $\hat{\theta} \pm 1.96 \times \mathrm{SE}(\hat{\theta})$, using whichever SE method is selected. For the NDE, the SE is the regression standard error of the exposure coefficient; for the NIE, the SE is the delta-method (Sobel) standard error by default, the bootstrap standard deviation when se_method = "bootstrap", or the delta-method SE when se_method = "composite" (the composite test replaces only the NIE p-value, leaving the SE and CI unchanged). The simulation benchmarks report two SE-related metrics per estimator: the mean estimated SE across replicates, which assesses whether the analytic SE matches the empirical sampling variability, and the empirical 95% CI coverage rate, defined as the fraction of replicates in which the true effect falls within the Wald interval. Coverage is assessed under the delta-method SE throughout the benchmarks; bootstrap SEs are evaluated separately for estimators where the delta-method variance is unreliable (e.g., COCA's ratio estimator). A caveat is that the delta-method SEs do not propagate first-stage estimation uncertainty. The outcome-stage regression treats the fitted mediator $\widehat{M}$ as if it were observed, so the reported SEs are conditional on the first-stage fit and may underestimate the true sampling variance when the first-stage instrument is weak.

**Supplementary Methods S5. Composite null hypothesis test for the NIE**

The NIE $= \alpha_M \times \beta_M$ is a product of two coefficients, so the null $H_0: \alpha_M \beta_M = 0$ is composite, holding under any of three cases: $H_0^{(1)}: \alpha_M = 0 \wedge \beta_M = 0$, $H_0^{(2)}: \alpha_M \neq 0 \wedge \beta_M = 0$, or $H_0^{(3)}: \alpha_M = 0 \wedge \beta_M \neq 0$. The Sobel (delta-method) test approximates the product as normal, which is valid only under the partial-null cases $H_0^{(2)}$ and $H_0^{(3)}$; under the point null $H_0^{(1)}$ (sparse signals), the product of two independent normals follows a normal product distribution with density $f(z) = K_0(|z|)/\pi$ (where $K_0$ is the modified Bessel function of the second kind), making the Sobel test conservative.[16]

ICONIC provides a third inference option (`se_method = "composite"`) implementing the closed-form JT-comp test of Huang (2019).[16] The composite p-value is

$$\hat{p}_{\text{comp}} = F\left(\frac{ab}{\sqrt{\widehat{\text{Var}}(a)}}\right) + F\left(\frac{ab}{\sqrt{\widehat{\text{Var}}(b)}}\right) - F(ab),$$

where $a = \hat{\alpha}_M/\text{SE}(\hat{\alpha}_M)$ and $b = \hat{\beta}_M/\text{SE}(\hat{\beta}_M)$ are the standardized z-statistics, $\widehat{\text{Var}}(a)$ and $\widehat{\text{Var}}(b)$ are their variances across the collection of tests sharing the same estimator, and $F(z) = 2\int_{|z|}^{\infty} K_0\,(x)/\pi\, dx$ is the CDF of the standard normal product distribution. The variances are estimated per estimator across all tests sharing that estimator, i.e., across mediators (and outcome features, when present) within each method. In ICONIC's case study design, the outcome $Y$ is a scalar (e.g., birth weight) and $M$ is a panel of mediators (e.g., placental genes), so the variation in $a$ comes from the different stage-1 regressions $M_m \sim X$ for each mediator $m$. When fewer than 5 tests are available, the point-null value $\text{Var} = 1$ is used. When `se_method = "composite"`, only the NIE p-value changes; the NDE p-value, NIE standard error, and all point estimates are unchanged. The output also includes `var_a` and `var_b` columns for transparency.

The JT-comp approximation is derived via a Taylor-series expansion (Theorem 3.3 of Huang 2019) whose error term vanishes rapidly when signals are sparse and $\text{Var}(a), \text{Var}(b) < 1.5$ (approximately $n < 2000$). ICONIC clamps the estimated variances to $[1,1.5]$ to enforce this validity range. The lower bound of 1 corresponds to the point-null calibration and guards against the anti-conservative behavior that arises when the sample variance is deflated below 1 by positive correlation among tests sharing the same samples. Type I error is slightly inflated at small p-values (e.g., $\sim 15\times$ at the genome-wide significance level $5 \times 10^{-7}$), as documented by Huang (2019) and Du et al. (2023);[17] at conventional thresholds ($\alpha = 0.05$) the inflation is modest.

*Limitation under the partial null.*

The JT-comp test cannot distinguish the partial-null case $H_0^{(2)}$ ($\alpha_M \neq 0, \beta_M = 0$) from the alternative when $\alpha_M$ is large, because the product $ab = a \cdot b$ can be large even when $b \sim N(0,1)$. This inflates type I error under $H_0^{(2)}$ for estimators with strong stage-1 instruments. The Sobel test handles this case correctly via the delta-method SE, which scales the product by $|\hat{\alpha}_M| \cdot \text{SE}(\hat{\beta}_M)$ under $H_0^{(2)}$. When $\alpha_M$ is expected to be nonzero and signals are not sparse, `"delta"` may therefore be preferable; `"composite"` is most beneficial when signals are sparse (the point-null $H_0^{(1)}$ is the dominant case).

**Supplementary Methods S6. Negative-control validity checks**

The validity of a negative-control analysis rests on three assumptions: that each control is independent of the exposure (A1) and of the genetic instrument (A2) given covariates and the unmeasured confounder, and that the valid controls are numerous enough to span the latent confounders (A3). When a mediator-specific instrument $G_m$ is used, a fourth assumption is required, namely that each control is independent of the mediator instrument given covariates and the confounder (A2'). Violations bias the bridge-function estimators, so ICONIC implements an empirical check for each. We state the assumptions conditional on both $C$ and $U$ (e.g., $W \perp X \mid C, U$) because the identifying content of a negative control is that it shares $U$ with the outcome but is not independently driven by the exposure once $U$ is accounted for; conditioning on $C$ alone is the empirically testable projection of this condition, since $U$ is unobserved.[18]

**A1** (negative controls are independent of the exposure given covariates and the confounder; $W \perp X \mid C, U$). To test the empirically observable projection of this assumption, we regress each negative-control feature $W_f$ on exposure $X$ (adjusting for covariates $C$) and flag controls with significant $X$-associations. ICONIC supports two screening criteria: Benjamini-Hochberg (BH) FDR control at 0.10 (`criterion = "fdr"`), and a magnitude-based criterion (`criterion = "magnitude"`) that flags controls whose $W \sim X$ association exceeds a user-specified effect-size threshold (default 0.10). The magnitude criterion is better suited to the negative-control setting, where the intended confounder-sharing signal produces a $W$–$X$ association by construction (because $X$ is a proxy for $U$), so an FDR-only screen flags all valid controls as violations. The default `criterion = "both"` requires both FDR significance and a magnitude exceeding the threshold. In the model selection workflow, A1 is reported but not used for eligibility, because A1 cannot distinguish a control that is downstream of the exposure (a true violation) from one that shares a cause with $X$ (the intended negative-control behavior).

**A2** (negative controls are independent of the genetic instrument given covariates and the confounder; $W \perp G_1 \mid C, U$). To test this assumption, we compute the partial correlation of each $W_f$ with instrument $G_1$ (residualizing both on $C$), apply BH-FDR, and flag significant $G_1$-associations. The COCA estimator is exempt from the A2 screen: COCA calibrates through the $W \sim Y + X$ ratio and does not require $W \perp G_1$, so `iconic_diagnose()` re-counts the valid controls for COCA using an A1-only magnitude screen (`criterion = "magnitude"` with a stricter `magnitude_threshold = 0.75`, which flags only gross $X$-dependence rather than the intended confounder-sharing signal).

**A2'** (negative controls are independent of the mediator instrument given covariates and the confounder; $W \perp G_m \mid C, U$) using the same procedure as above.

**A3** (the valid negative controls capture the confounder covariance; completeness). ICONIC operationalizes completeness as a two-component condition, reflecting that spanning the confounder subspace is about the covariance the controls capture rather than merely their count. The dimensional component compares the number of valid negative controls $n_{\text{valid}}$ against the number of latent confounders $k$: the verdict is "satisfied" when $n_{\text{valid}} > k$ (strictly), "borderline" when $n_{\text{valid}} = k$, and "under-identified" when $n_{\text{valid}} < k$; this is necessary but not sufficient, because a panel can have many controls that each weakly capture $U$. The covariance-capture component tests whether adding $W$ to a model of the outcome above covariates $C$ alone captures a meaningful share of $U$'s contribution. We compute the incremental $R^2$ of $W$ for $Y$ above $C$ ($R^2(W \mid C) = R^2(Y \sim C + W) - R^2(Y \sim C)$, averaged across outcome features) and assess its magnitude against strong (default $R^2 > 0.3$, with permutation $p < 0.05$) and weak ($R^2 \geq 0.1$) thresholds, with a permutation null (permuting the $W$–$Y$ association while holding the $C$–$Y$ association fixed) providing a p-value for "W captures $U$-signal beyond chance." The composite verdict combines both components. The dimensional verdict stands when capture is strong or weak, but a dimensionally passing panel ("satisfied" or "borderline") whose capture is negligible is downgraded to "weak-capture"; "under-identified" is returned whenever $n_{\text{valid}} < k$. In the model selection workflow, A2 (instrument-independence) serves as the primary gate for counting valid controls; the standalone `nc_completeness_check()` uses both A1 and A2 when an instrument is available, falling back to A1 alone otherwise, and returns the dimensional verdict, the capture estimate, and the composite verdict.[18]

#### Supplementary Methods S7. Exposure instrument ($G_1$) construction via LDpred2-auto

In case study 1, the exposure instrument $G_1$ is a polygenic score for maternal gestational diabetes risk, constructed via LDpred2-auto Bayesian shrinkage regression[19] as implemented in the bigsnpr R package. The goal is to aggregate weak, genome-wide genetic effects on OGTT 2-hour glucose into a single composite instrument, replacing the simpler clumping-and-thresholding (P+T) approach with a method that jointly models all variants while accounting for linkage disequilibrium (LD).

GWAS summary statistics were obtained from Gu et al. (2025)[20] (GWAS Catalog accession GCST90566423), a genome-wide association study of OGTT 2-hour plasma glucose in 84,743 individuals of Chinese ancestry. Summary statistics were quality-controlled before matching: variants with missing beta, standard error, chromosome, position, or allele annotations were removed; variants with extreme beta values ($> 10 \times$ IQR from the median) were dropped as likely artifacts; and ambiguous A/T or C/G strand-inseparable SNPs were excluded. After QC, summary statistics were matched to the LD reference panel via bigsnpr's `snp_match()`, which handles allele flipping and strand reversal internally.

The LD reference panel was derived from the 1000 Genomes Project Phase 3 by subsetting to East Asian (EAS) individuals (CHB, CHS, JPT, CDX, KHV; $n \approx 504$) using plink2 with basic QC (MAF ≥ 0.01, genotype missingness ≤ 0.1). Variants were further restricted to the HapMap3 SNP list to improve LDpred2 stability, as recommended by the method's authors. A final QC step compared two independent estimates of genotype standard deviation, one derived from GWAS effect sizes ($\mathrm{sd}_{\mathrm{ss}} = \sqrt{n_{\mathrm{eff}} \cdot \mathrm{SE}^2 + \beta^2}$) and one from allele frequencies in the LD reference ($\mathrm{sd}_{\mathrm{geno}} = \sqrt{2f(1-f)}$), and removed variants where the ratio $\mathrm{sd}_{\mathrm{ss}}/\mathrm{sd}_{\mathrm{geno}}$ fell outside $[0.5,2]$, indicating allele/strand mismatches or frequency discordance between the GWAS and the LD reference.

A sparse LD correlation matrix was computed from the EAS reference genotypes using `snp_cor()`, and SNP-based heritability was estimated via LD score regression (`snp_ldsc2()`) to initialize the LDpred2-auto model. LDpred2-auto was then run with 30 chains spanning a range of causal variant proportions ($p$ from $10^{-4}$ to 0.2 on a log scale), 500 burn-in iterations and 500 sampling iterations per chain, with `allow_jump_sign = FALSE` and `shrink_corr = 0.95` for stability. Chains were filtered by convergence (keeping those whose posterior correlation estimate range exceeded 0.95 × the 95th percentile across all chains), and the final effect sizes were averaged across converged chains.

The averaged LDpred2 effect sizes were matched back to the GUSTO maternal genotype columns via `snp_match()`, with beta negated where GUSTO stored the opposite allele as the effect allele. Missing genotypes were imputed by column (variant) mean, and the polygenic score was computed as the matrix product of the genotype dosage matrix and the effect-size vector, then centered and scaled to unit variance to produce the final $G_1$ instrument.

### Supplementary Methods S8. Elastic net composite instrument construction

In case study 1, the mediator instrument $G_m$ is constructed as a cross-validated elastic net of cis-eQTLs, producing an $n$-length predicted-expression vector per gene rather than a single-SNP dosage. For each expressed gene, cis-SNPs within $\pm 1$ Mb of the gene's transcription start site (TSS, strand-aware) were extracted from the dosage matrix, and a cross-validated elastic net (cv.glmnet, $\alpha = 0.3$, 5-fold CV, type.measure = "mse", standardize = TRUE) was fit on the residualized cis-SNP dosages to predict the residualized gene expression. Both expression and dosages were residualized on the full covariate set (sex, maternal ethnicity, gestational age, fetal ancestry PCs, HCP latent factors, RUVr unwanted-variation factors) via Frisch-Waugh-Lovell projection, ensuring that the fitted SNP effects are covariate-adjusted and the resulting instrument is orthogonal to the covariates ICONIC conditions on. Out-of-fold (prevalidated) predictions at $\lambda_{\min}$ were extracted from cv.glmnet's fit.preval, ensuring that each sample's prediction comes from the fold in which it was held out and is not inflated by in-sample overfitting. Genes were retained as eligible mediators if the elastic net satisfied three criteria: (1) at least one non-zero SNP coefficient at $\lambda_{\min}$; (2) a signed out-of-fold prediction $R^2$ exceeding 0.005 (computed as $1 - \mathrm{MSE}_{\text{oof}}/TSS$, which can go negative to flag anti-predictive models); and (3) a one-sided correlation $p$-value $< 0.10$ between out-of-fold predictions and residualized expression. The signed $R^2$ is used rather than $\mathrm{cor}(\hat{y}, y)^2$ to avoid admitting intercept-only models whose negative between-fold correlation, when squared, spuriously clears the threshold. The composite instrument produces a predicted-expression matrix $G_m$ (genes $\times$ samples), per-gene SNP weights, and a QC table with per-gene cis-SNP count, out-of-fold $R^2$, correlation, $p$-value, and $\lambda_{\min}$.

The single best cis-eQTL per gene was identified via Matrix eQTL (cis window $\pm 1$ Mb, $p < 10^{-3}$ recording threshold) with Benjamini-Hochberg FDR correction at 10% across all tested genes. The elastic net composite instrument yields more eligible mediator genes and higher partial-$F$ statistics than the single-SNP approach because it aggregates regulatory signal across multiple cis-variants rather than relying on a single variant.

**Supplementary Methods S9. Negative-control panel ($W$) construction from methylation PCs**

In case study 1, the negative-control panel $W$ is constructed from maternal peripheral blood DNA methylation data. The rationale is that methylation patterns in maternal blood capture systemic confounding factors (maternal metabolic state, inflammation, cell composition) that influence both the exposure (GDM) and the outcome (birth weight) through pathways that bypass placental gene expression, satisfying the negative-control assumptions when used as $W$.

DNA methylation was profiled in 915 maternal buffy coat samples using the Illumina Infinium MethylationEPIC BeadChip (EPIC 850k array).[21] Raw IDAT files were processed in R with the minfi package. Probes with fewer than three beads in either channel or detection $p$-value $\geq 0.01$ were removed, along with Y-chromosome probes, cross-hybridizing probes, and probes containing SNPs at the CpG site or single-base extension. Within-sample normalization was performed with Noob preprocessing, and beta values were converted to M-values before applying ComBat to remove chip effects. Probes with a methylation range (max $-$ min, excluding outliers) below 5% were removed, yielding 422,788 quality-controlled CpG sites.

To construct $W$, the M-value matrix was first regressed on cell composition and technical covariates via ordinary least squares. Cell proportions for six types (CD8T, CD4T, NK, B-cell, monocyte, neutrophil) were estimated using a cell-type-specific reference panel and included alongside the first six cell-proportion PCs. Technical covariates comprised processing plate, hospital, DNA extraction batch, and maternal age. The residual matrix was obtained by subtracting the fitted values from the centered M-value matrix, computed in chunks of 5,000 probes to manage memory. Principal component analysis (via `irlba` truncated SVD) was then performed on the transposed residual matrix (samples $\times$ probes), and the top 20 principal component scores were retained as the $W$ panel. Each PC serves as a negative-control feature, capturing orthogonal axes of residual methylation variation after removing known cell and technical effects. The resulting $W$ matrix (20 features $\times$ samples) is passed to ICONIC's `nc_validity_screen()` to verify the A1 ($W \perp X \mid C$), A2 ($W \perp G_1 \mid C$), and A2' ($W \perp G_m \mid C$) independence assumptions before use in estimation.

**Supplementary Methods S10. Exposure instrument ($G_1$) construction from a GSCAN cigarettes-per-day polygenic score**

In case study 2, the exposure instrument $G_1$ is a published polygenic score for cigarettes per day, PGS003368, obtained from the PGS Catalog. This score derives from the GSCAN meta-analysis of smoking and drinking behaviours (Saunders et al. 2022, $n \approx 3.4$ million),[22] and its weights are already LDpred2-shrunk (1,055,811 variants, European ancestry, GRCh38 harmonized positions). Because the score is pre-weighted, no re-estimation from summary statistics is required; $G_1$ is obtained by direct scoring rather than by the LDpred2-auto pipeline used in case study 1 **(Supplementary Methods S7)**.

The harmonized scoring file (`PGS003368_hmPOS_GRCh38.txt.gz`) was reduced to a three-column plink2 score specification (variant ID, effect allele, weight; no header) and applied to the TCGA post-imputation, post-QC genotypes with `plink2 --pfile <tcga_imputed_qc> --score <score_file> 1 2 3 no-mean-imputation`. The `no-mean-imputation` flag forces plink2 to skip variants absent from the target genotypes rather than imputing them at the sample mean allele dosage, so the score reflects only observed dosages. The resulting per-sample allelic score was read from the `.sscore` output, centered, and scaled to unit variance to yield the final $G_1$ instrument.

The instrument targets the continuous exposure $X$ = pack-years, a lifetime cumulative smoking measure derived from the TCGA clinical record. Instrument strength was checked as a first-stage regression of pack-years on $G_1$; because an unadjusted first stage would be confounded by ancestry (both $G_1$ and pack-years vary with genetic background), the reported instrument strength is the *partial* $F$ statistic for $G_1$ from a model that also conditions on the genotype ancestry principal components, comparing the full model $X \sim G_1 +$ PCs against the reduced model $X \sim$ PCs. All exposure and genotype records were joined at the patient level (the first twelve characters of the TCGA barcode).

**Supplementary Methods S11. Mediator instrument ($G_m$) construction from GTEx v11 lung cis-eQTLs**

In case study 2, the mediator instrument $G_m$ uses the same cross-validated elastic net architecture as case study 1 **(Supplementary Methods S8)**, with two case-study-specific changes: the cis-SNP set for each gene is restricted to variants that are significant lung cis-eQTLs in GTEx, and the covariate set is the TCGA lung-cohort covariate set rather than the GUSTO placental one.

The candidate cis-SNP set was defined from the GTEx v11 lung significant variant-gene pairs (`Lung.v11.eQTLs.signif_pairs.parquet`). For each expressed gene, cis-SNPs within $\pm 1$ Mb of the transcription start site that also appeared as a significant lung eQTL for that gene in GTEx v11 were extracted from the TCGA dosage matrix; restricting to GTEx-significant lung eQTLs (rather than all cis-SNPs) focuses the instrument on variants with prior evidence of regulatory activity in the disease-relevant tissue. Both expression and dosages were residualized on the full TCGA covariate set (age at diagnosis, sex, histological subtype, tumor stage, smoking status, genotype ancestry principal components, HCP latent factors, and RUVr unwanted-variation factors) via Frisch-Waugh-Lovell projection, so the fitted SNP effects are covariate-adjusted and $G_m$ is orthogonal to the covariates ICONIC conditions on. A cross-validated elastic net (`cv.glmnet`, $\alpha = 0.3$, 5-fold CV, `standardize = TRUE`) was fit on the residualized cis-SNP dosages to predict residualized expression, and out-of-fold (prevalidated) predictions at $\lambda_{\min}$ were extracted from `cv.glmnet`'s `fit.preval` so that each sample's prediction comes from the fold in which it was held out.

Genes were retained as eligible mediators if the elastic net produced at least one non-zero SNP coefficient at $\lambda_{\min}$ and a signed out-of-fold prediction $R^2$ exceeding 0.002 (computed as $1 - \mathrm{MSE}_{\mathrm{oof}}/\mathrm{TSS}$, which can go negative to flag anti-predictive models). The signed $R^2$ is used rather than $\mathrm{cor}(\hat{y}, y)^2$ so that intercept-only or anti-predictive fits are excluded rather than admitted through a squared negative correlation. Of 3,674 genes tested, 3,469 passed this filter and were carried forward as mediators. The composite instrument yields a predicted-expression matrix $G_m$ (genes × samples) together with per-gene SNP weights and a QC table (cis-SNP count, out-of-fold $R^2$, correlation, and $\lambda_{\min}$). GTEx v11 data were obtained from the GTEx Portal.[23]

### Supplementary Methods S12. Negative-control panel ($W$) construction from tibial-nerve predicted expression

In case study 2, the negative-control panel $W$ is built from genetically predicted expression in a tissue that is off the smoking-to-lung-cancer-survival causal path. Whereas case study 1 used maternal blood methylation PCs **(Supplementary Methods S9)**, case study 2 uses GTEx v8 tibial-nerve predicted expression. Tibial nerve is a non-lung tissue whose genetically regulated expression is not plausibly on the causal path from smoking to lung-tumor progression, so its predicted-expression axes serve as negative-control features $W$ that capture ancestry- and genotype-linked variation without carrying the mediating signal.

Tissue-specific expression weights were taken from the GTEx v8 tibial-nerve FUSION models (`GTExv8.EUR.Nerve_Tibial.nofilter.pos` and the associated `.wgt.RDat` weight files). For each gene, the FUSION weight object stores several trained predictors (e.g. BLUP, LASSO, elastic net, single best eQTL); the predictor with the highest cross-validated $R^2$ (`cv.performance["rsq", ]`) was selected as the gene's model, matching FUSION's own best-model convention. Weights were matched to the TCGA imputed dosages by translating rsIDs to chromosome-position-ref-alt keys via a dbSNP GRCh38 lookup, and genetically predicted expression was computed as the dosage-by-weight product per gene. Principal component analysis (`prcomp`) was then run on the predicted-expression matrix, and the top 20 principal component scores were retained as the $W$ panel (20 features × samples). This matrix is passed to ICONIC's `nc_validity_screen()` to test the A1 ($W \perp X \mid C$), A2 ($W \perp G_1 \mid C$), and A2' ($W \perp G_m \mid C$) independence assumptions before estimation. GTEx v8 FUSION weights were obtained from the FUSION/TWAS resource.[23]

**Supplementary Methods S13. Binary outcome support**

ICONIC extends all estimators except COCA to binary (0/1) outcomes via two-stage predictor substitution (2SPS), exactly as for survival outcomes. The first-stage regressions (instrumenting $X$ on $G_1 + W + C$, instrumenting $M$ on $\hat{X} + G_m + W + C$, and constructing the bridge proxies $\widehat{W}$) involve only continuous responses ($X$, $M$, $W$), so they remain ordinary least squares regardless of the outcome type. Only the final outcome stage changes: it replaces OLS on a continuous $Y$ with a logistic regression of the 0/1 outcome (log-odds-ratio scale) or a linear probability model fit by OLS on the 0/1 outcome (risk-difference scale). This preserves the identifying structure of each estimator (the instrument purges confounded variation in $X$, and the bridge proxies absorb the residual confounding) while respecting the Bernoulli outcome.

*Two effect scales.*

ICONIC provides two effect scales for binary outcomes. The logistic log-odds-ratio (log-OR) scale fits `glm(y ~ X_hat + M_hat + W_hat + C, family = binomial)` and reports the coefficients as log-ORs. Like the Cox log-HR, the log-OR is non-collapsible: the conditional log-OR given covariates differs from the marginal log-OR even under no confounding, so the product decomposition NIE $= \hat{\alpha}_M \cdot \hat{\beta}_M$ is approximate on this scale. The risk-difference scale fits a linear probability model (OLS on the 0/1 outcome) and reports coefficients as differences in event probability. The risk difference is collapsible, so the NDE/NIE decomposition is exact on this scale; however, the linear probability model can produce fitted probabilities outside $[0,1]$, and its model-based standard errors are heteroskedasticity-naive. The log-OR scale is recommended as the primary effect scale; the risk-difference scale provides a collapsible complement when marginal effects are of interest. This log-OR/risk-difference duality mirrors the log-HR/RMST duality for survival outcomes **(Supplementary Methods S1)**.

*COCA incompatibility.*

COCA regresses the negative control on the outcome ($W \sim Y + X$) and recovers $\hat{\tau} = -\hat{\beta}_X/\hat{\beta}_Y$, a linear-ratio calibration that presumes the outcome enters the negative-control regression as a continuous, linearly related response. A binary 0/1 outcome does not satisfy this assumption, so the linear COCA ratio recovers neither the causal log-OR nor the risk difference. As with survival outcomes, COCA is therefore structurally excluded for binary outcomes: ICONIC returns `NA` for COCA on binary data with a `reason` attribute explaining the incompatibility, and the estimator is dropped from the eligibility report.

*Interface.*

Binary outcomes are specified at `iconic_data()` construction: `iconic_data(outcome_type = "binary", ...)` with a 0/1 outcome vector. The effect scale is selected at estimation: `iconic_estimate(effect_scale = "logor")` (default) or `iconic_estimate(effect_scale = "riskdiff")`. Requesting a survival scale (`"loghr"` or `"rmst"`) on a binary outcome is undefined; ICONIC emits a message and falls back to `"logor"`. All downstream reporting functions (`iconic_diagnose()`, `iconic_recommend()`) operate identically on binary and continuous data, because the diagnostic screens depend only on the first-stage regressions, which remain OLS.

*Unsupported features.*

As for survival outcomes, `infer_confounding()` and the data-calibrated (learned) generative texture are not available for binary outcomes: the gap-based confounding calibration relies on a continuous outcome scale. `infer_confounding()` therefore stops with an informative error instructing the analyst to supply confounding parameters explicitly (`confounding = "manual"`) or use the defaults; `iconic_sensitivity()` and `iconic_prospect()` called with `confounding = "inferred"` error accordingly, and the GAN-plus-copula texture falls back to the default benchmark texture with a message. Sensitivity and prospective analyses for binary outcomes therefore use the default (benchmark) texture rather than a cohort-learned texture.

*Benchmark design.*

The binary benchmark uses the same structural DGP as the continuous and survival benchmarks (`generate_toy_data`), with the linear predictor $\eta = \beta_X X + \alpha_M \beta_M M_{\text{path}}$ mapped to a Bernoulli outcome via the logistic link, $\Pr(Y = 1) = \text{plogis}(e_0 + \eta_c)$, where $\eta_c$ is the centered linear predictor and the intercept $e_0$ is solved by root-finding (`uniroot`) so that the marginal event probability equals the target prevalence (fixed at $0.5$). The total-effect sweep uses $\alpha_M = 0$, $\beta_M = 0$ (no mediation path), so the true total effect on the log-OR scale is $\beta_X = 0.25$. The mediation sweep uses $\alpha_M = 0.50$, $\beta_M = 0.30$ (true NDE $= 0.25$, true NIE $= 0.15$ on the log-OR scale), with $k = 2$ confounders under distinct path loadings, `mo_confounding` $= 0.8$, $\rho_{G1} = 0.3$, $\omega_1 = \omega_2 = 0.7$, matching the continuous and survival mediation benchmarks. The risk-difference truth is not analytically tractable because it depends on the calibrated intercept and the outcome nonlinearity; it is computed empirically by running the UNADJ estimator at zero confounding with $n = 2{,}000$, yielding TE $= 0.0620$, NDE $= 0.0607$, NIE $= 0.0342$. Fifty replicates per cell, $n = 500$, five confounding levels $\delta \in \{0.2, 0.4, 0.6, 0.8, 1.0\}$, and both effect scales are benchmarked **(Supplementary Figure S17)**.
